# Urinary collagen peptides predict mortality

**DOI:** 10.1101/2025.06.05.25329025

**Authors:** Maria Frantzi, Felix Keller, Agnieszka Latosinska, Joachim Beige, Alexandre Mebazaa, Anaïs Caillard, Dewei An, Paul Perco, Joost P. Schanstra, Lorenzo Catanese, Ralph Wendt, Harald Rupprecht, Jan A. Staessen, Antonia Vlahou, Harald Mischak, Justyna Siwy

**Author notes:** Corresponding author: Dr. Justyna Siwy, PhD mosaiques diagnostics GmbH, Rotenburger Straße 20, 30659 Hannover, Germany. equally contributing authors (Maria Frantzi, Felix Keller, Agnieszka Latosinska).

## Abstract

**Background:** Organ fibrosis caused by the presence of excessive extracellular matrix (ECM) is strongly related to mortality. Urinary peptide signatures were reported predictive of death in SARS-CoV-2 and chronic kidney disease. Such signatures were composed for 68% of collagen fragments. In this study, we examined whether an exclusively collagen-based urinary peptide model, potentially representing organ fibrosis, could predict mortality in patients with critical and non-critical conditions.

**Methods:** Urinary proteomic data from 1,012 patients infected with SARS-CoV-2 were considered to evaluate the association of collagen peptide levels with short term mortality. Additional datasets from 9,193 patients were used for validation, including 1,719 patients sampled at intensive care unit (ICU) admission and 7,474 patients with other diseases (outside the ICU) were extracted from the Human Urinary Proteome Database.

**Findings:** A collagen peptide-based model based on 210 peptides (COL210) predicting mortality was developed for patients after SARS-CoV-2 infection. This model was validated in patients in (ICU_HR_: 2·64; 1·71-4·10; p<0·001) and outside the ICU (Non-ICU_HR_: 2·16 95% CI: 1·47–3·17; p<0·001), showing strong associations to mortality regardless underlying conditions.

**Interpretation:** This study demonstrates a link between the presence of ECM fragments in urine, specifically collagens, and increased mortality risk. The availability of such a non-invasive collagen-based predictor of mortality may serve as basis for proteomics guided targeted intervention.

**Funding:** This study was funded by “DisCo-I” (HORIZON–MSCA; 101072828), “SIGNAL” (BMBF; 01KU2307, FWF; project number I 6471 and Grant-DOI 10.55776/I6471 and ANR-22-PERM-0002-06), “UriCov” (BMG,2523FSB114), by “Accurate-CVD” (BMWK), by UPTAKE (BMBF; 01EK2105A-C) and MULTIR (101136926)

**Research in Context:** *Evidence before this study:* Fibrosis, marked by excessive extracellular matrix (ECM) deposition, is a key factor in chronic diseases and organ failure and is correlated with increased mortality in conditions such as idiopathic pulmonary fibrosis (IPF), chronic kidney disease (CKD), liver disease, cardiovascular disease (CVD), and cancer. A literature search conducted on 10/12/2024 using the MeSH terms “collagen” AND “fibrosis” AND “mortality” OR “death” OR “failure”, revealed a shift from static histological assessments of ECM deposition to a more dynamic approach based on the assessment of biomarkers of collagen turnover within specific fibrotic conditions, which offer real-time and potentially actionable insights into disease progression and predict adverse outcomes, including mortality. Key biomarkers include products of collagen synthesis (e.g., pro-peptides of type III, V, and VI collagen), but also of collagen degradation. These biomarkers have individually shown correlations with fibrosis severity and mortality, with links to disease progression and survival in IPF, CKD, CVD, and cancer. In heart failure, urinary peptides from collagen α-1 (I) chain predict adverse events and mortality. Collagen α-1 (XXIV) chain was also among the circulating plasma proteins that were related with transplant-free survival of patients with IPF.

*Added value of this study:* This work further adds to the growing relevance of collagen turnover in the context of mortality across fibrotic diseases, in an effort to generalize evidence for collagen degradation markers, independent from the underlying pathological condition. It additionally explores integration of collagen degradation fragments into risk models, potentially improving predictive accuracy and advancing precision medicine.

*Implications of all the available evidence:* Our study demonstrates that urinary collagen degradation markers integrated into a machine learning model (COL210) can identify “vulnerable” individuals from different fibrotic backgrounds, offering potential for improved prognostication and targeted interventions.

## INTRODUCTION

Extracellular matrix (ECM) is the non-cellular key component of the microenvironment of cells with various crucial functions.^1^ As a highly dynamic structure, it continuously undergoes remodelling that under physiological conditions is a critical mechanism to regulate cell differentiation, establish and maintain stem cell niches, branch morphogenesis, but also support angiogenesis and wound repair, when necessary.^2, 3^ ECM, thus, plays a vital role in maintaining tissue structure and function, but when improperly regulated, can lead to fibrotic tissue formation.

Fibrosis is characterized by the excessive deposition and remodelling of ECM, particularly collagens.^4^ It leads to stiffening and scarring of the organ tissue, is a hallmark of chronic diseases and a critical driver of increased mortality across diverse pathological conditions, including idiopathic pulmonary fibrosis (IPF),^5–12^ chronic kidney disease (CKD),^13–15^ liver disease,^16^ heart failure (HF),^17–21^ and cancer.^21–24^ In the context of organ fibrosis, the excessive ECM deposition impairs organ function by disrupting normal cellular architecture and interfering with cellular processes such as nutrient and oxygen diffusion.^25^ Key drivers of fibrosis include activation of fibroblasts, myofibroblast differentiation, and an inflammatory response, often due to chronic injury or inflammation.^26, 27^ These cells secrete ECM components and promote a pro-fibrotic environment, which amplifies fibrosis.^27^

However, synthesis only represents one arm of ECM turnover, degradation appears equally relevant, although less studied. Consequently, research has shifted focus from static histological measures of ECM deposition to dynamic assessment, including analysis of specific biomarkers of collagen degradation, which may be linked to disease progression and predictive of adverse outcomes, such as mortality, in fibrotic diseases.^8, 13, 16, 17, 20^ Dynamic biomarkers of collagen turnover can broadly be categorized into indicators of collagen synthesis, such as the pro-peptides generated during collagen assembly^16^ and degradation markers, collagen fragments from the mature collagen fibre, generated by e.g. matrix metalloproteinases (MMP)^28^. Previous reports from our group have revealed that peptide profiles in urine detected using capillary electrophoresis coupled to mass spectrometry (CE-MS) are indicative of an individual’s risk for disease progression and/or death.^29^ This observation was initially linked to specific diseases including diabetic and generally CKD,^30,31^ HF,^17^ SARS-CoV-2 infection^32^ or cancer progression and metastasis.^33^ However, a classifier primarily generated to predict disease course after SARS-CoV-2 infection (COV50 classifier) was also predictive of mortality even in the absence of SARS-CoV-2 infection, demonstrating a pre-existing vulnerability.^34^ Similarly, urinary peptidomics profiles were predictive of mortality risk post-acute SARS-CoV-2 infection.^35^ These classifiers largely consist of naturally excreted urinary collagen fragments that result from collagen degradation rather than from collagen synthesis (i.e. pro-collagens).^36^ The observation that these degradation-derived collagen peptides are associated with mortality across various conditions -such as CKD, HF, and post–SARS-CoV-2 infection^34, 35, 37, 38^-has led to the hypothesis that a solely collagen peptide-based model may depict mortality risk that is not restricted to a specific underlying pathology or morbidity^39–41^ Based on this hypothesis, in this study we focus on urinary collagen peptides, representing collagen degradation fingerprint, aiming to evaluate possible commonalities of chronic pathological conditions as depicted by altered collagen fragment excretion in urine. If specific collagen peptide profiles are associated with mortality, this fact could be exploited to guide intervention with existing therapeutic options or by introducing novel biologically driven targets to manipulate ECM turnover.

## METHODS

### Study population and design

A total of four distinct cohorts were included in the study. The development cohort was used to identify collagen peptide biomarkers in urine associated with mortality and to develop a predictive classifier. Two additional, independent cohorts were employed to validate the classifier and assess its performance in predicting mortality across different datasets.

Furthermore, based on the hypothesis that collagen peptides associated with mortality may also reflect underlying fibrosis, a fourth cohort of CKD patients with available data on fibrosis severity (interstitial fibrosis and tubular atrophy, IFTA) was analyzed to examine the correlation between classifier scores and the degree of fibrosis. The study design is illustrated in **Figure 1**.

**Figure 1:**
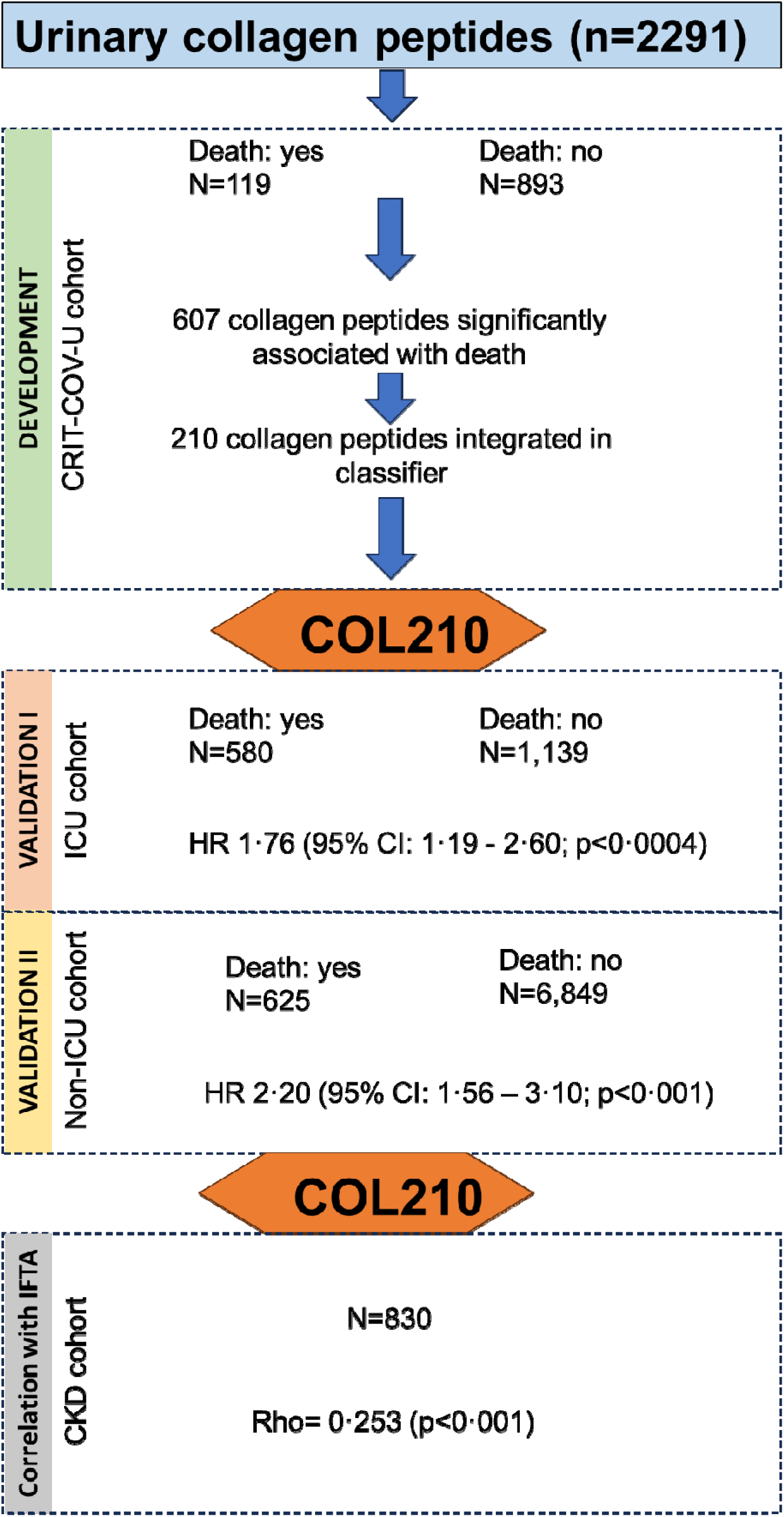
Study design.

### Patients with SARS-CoV-2 infection (CRIT-COV-U cohort)

Data of the entire CRIT-COV-U cohort (n=1,012) were used as development cohort. ^32^ Urinary collagen peptide abundance was analysed in 893 patients who survived the acute phase after a SARS-CoV-2 infection and 119 patients who died within 28 days after SARS-CoV-2 infection. This cohort was also utilized for machine learning modelling and optimization of the COL210 predictive model.

### ICU cohort

Data from 1719 patients from the medical, surgical, or mixed ICUs at 14 university hospitals from the FROG-ICU study were considered.^42^ These data were used as an independent validation cohort. Inclusion criteria were mechanical ventilation or administration of vasoactive agents for at least 24 h. Exclusion criteria were age under 18, severe head injury with a Glasgow Coma Scale below eight, brain death or persistent vegetative state, pregnancy or breastfeeding, transplantation in the past 12 months, moribund status, and lack of social security coverage. All CE-MS datasets were complemented with one-year follow-up and information on relevant co-variables (age, body mass index (BMI), sex, blood pressure, estimated glomerular filtration rate (eGFR), presence of diabetes, kidney, cardiovascular disease, hypertension). No pre-selection criteria were applied. Patient characteristics are summarized in **Supplementary Table 1.**

### Non-ICU

The second validation of the COL210 model was performed in a non-ICU population using 7,474 datasets extracted from the Human Urinary Proteome Database.^43, 44^ These datasets were selected from previous studies including more than 50 individuals and with available follow-up information by applying the same criteria as above regarding the availability of co-variables. These datasets included patients with HF, diabetes, kidney diseases, or cancer, with patient characteristics provided in **Supplementary Table 2**

### CKD cohort

To explore the association between urinary collagen peptides and kidney fibrosis, data from CKD patients with available histological assessments of fibrosis severity (interstitial fibrosis and tubular atrophy, IFTA) were used.^45^ **Supplementary Table 3** includes the characteristic of this cohort.

#### Urinary mass spectrometry datasets

The urinary proteome is well characterized and reference standards are available.^46,47^ Proteome analysis was performed on urine samples collected at study inclusion and bio-banked until assayed. Detailed information on urine sample preparation, proteome analysis by CE-MS, data processing, and sequencing of the urinary peptides allowing identification of parental proteins is available in previous publications.^47–49^ All datasets were from previously published studies and fully anonymized.

#### Survival endpoints

In the CRIT-COV-U cohort all participants were followed up until recovery, hospital discharge, or death. In this, acute phase after a SARS-CoV-2 infection the vital status was collected.

In the FROG-ICU study, information on vital status was collected three, six and 12 months after ICU discharge, as previously described.^50^ For the non-ICU patients, vital status and outcome was assessed as described in the specific original studies.^17,30, 40, 41, 48, 51–62^

#### Statistics

In respect to descriptive statistics for the ICU and non-ICU samples, as shown in **Supplementary Tables 1 and 2**, median and 1^st^ and 3^rd^ quartile (IQR) were assessed for continuous variables and absolute (N) and relative frequencies (%) for categorical variables. Hypotheses of no differences in scale or distribution of patient characteristics between the death and non-death groups were tested with Wilcoxon– Mann–Whitney tests for continuous and with χ2-homogeneity tests for categorical variables. For the definition of the mortality associated peptides, the collagen fragments abundance levels between the death and non-death group of the CRIT-COV-U cohort were compared using the Wilcoxon–Mann–Whitney tests followed by Benjamini–Hochberg-based false discovery rate (FDR) correction. Mortality per person-time stratified by age and COL210 groups was estimated as the ratio of the number of the deceased to the sum of all patients’ observation times within each group scaled to 100 person-years. Unadjusted longitudinal risk of mortality was estimated as cumulative incidence using the Kaplan-Meier estimator for non-ICU and ICU patients. In the latter group, estimates were carried out over the whole observation time and separately within ICU and after ICU survival. Adjusted Cox regressions stratified by age groups were performed for non-ICU and ICU patients separately. Models were adjusted for sex, presence of kidney disease, cardiovascular disease, hypertension, mean arterial pressure, and eGFR. To describe the distribution of COL210 between deceased and surviving subjects within each study, boxplots were used. The relationship between time until death and the COL210 score (as derived based on the machine learning classification) is displayed in a scatter plot indicating ICU and non-ICU groups by colour and a local average smoother is applied to assess the relationship of COL210 on failure time. For statistical testing, we allow for a type 1 error of 5%, all hypotheses are two-sided. The hazard ratios (HR) were compared using z-test. All analyses were carried out using R (version 4.2.2, R Core Team, Vienna, Austria).^63^

#### Generation of the COL210 classifier

Urinary abundance levels of collagen peptides were modelled through a machine learning approach based on support vector machine (SVM) algorithms using MosaCluster proprietary software (version 1.6.5). All peptides demonstrating a significant difference (adjusted for the FDR set at 0·05) between death and non-death group were included in the classifier. The SVM model was optimized in the CRIT-COV-U cohort. The SVM binary classification model (any death event vs survivors) was optimized in k-fold cross-validation. The optimal C and gamma parameters were: 10,200 and 0·000004, respectively with radial basis function kernel.

### Ethical approval

All datasets were from previously published studies and fully anonymized. The Ethics Committee of the German-Saxonian Board of Physicians (Dresden, Germany; number EKBR88/20·1) and the Institutional Review Boards of the recruiting sites provided ethical approval for the CRIT-COV-U. For the ICU cohort, the study was approved by our Institutional Review Board (IRB) (Comité de Protection des Personnes—Ile de France IV, IRB n°00003835 and Commission d’éthique biomédicale hospitalo-facultaire de l’hôpital de Louvain, IRB n°B403201213352). For the CKD cohort the local ethics committee of the Friedrich-Alexander Universität Erlangen-Nürnberg provided approval for the nephrological biobank of the Klinikum Bayreuth (ethic approval code 264_20 B) and the urinary proteomics analysis (ethic approval code 221_20 B). Approval from the Ethics Committee of the Saxonian Board of Physicians, Dresden, Germany, was obtained for the study center Klinikum St. Georg Leipzig (ethic approval code EK-BR-14/20-1). In addition, ethical review and approval are not required for this study due to all data being fully anonymized, based on the opinion of the ethics committee of the Hannover Medical School, Germany (no. 3116-2016).

## RESULTS

### Collagen profiles in urine are predictive of survival after SARS-CoV-2 infection

Using CE-MS peptidomics datasets from a prospective multicenter cohort of 1,012 subjects with a confirmed SARS-CoV-2 infection (the CRIT-COV-U cohort),^32^ we first investigated whether urinary collagen peptides detected in this cohort (n=2,291) are significantly different in abundance in urine from patients who survived the acute disease phase (n=893) in comparison to those who died within 28 days (n=119) after a SARS-CoV-2 infection. This analysis resulted in the identification of 607 urinary collagen peptides significantly associated with death (FDR-adjusted p<0·05, **Supplementary Table 4**).

At the level of single collagen peptides, of the peptides significantly associated with mortality, 208 (23·3%) are derived from COL1A1, 121 reduced, and 87 increased in abundance. Notably, the most significant COL1A1 peptides (top significant based on adjusted p-value) are reduced in abundance and correspond to relatively large peptides (with molecular weight > 3,000 Da). The second most frequent were fragments of COL3A1, with also high number of reduced peptides (51 of 96 mortality-associated COL3A1 peptides). These were followed by fragments of COL1A2 (n=23; 15 reduced in abundance) and COL2A1 (n=31; 22 reduced in abundance).

The significant collagen peptides were combined into an SVM classifier. The classifier was trained and optimized using a take-one-out procedure in the 1,012 CRIT-COV-U data. The classifier generated, termed COL210, included finally 210 different collagen fragments.

### COL210 externally validated in predicting survival in patients under critical condition

COL210 was first independently validated in patients in critical condition (ICU) (n=1,719 datasets, **Supplementary Table 1**). ^37^ Of the 1,719 subjects, 580 (33·7%) died during a median follow-up of 12 months. This included 287 deaths occurring in the ICU and 293 at a median of 47 days following hospitalisation. Survival curves estimated using COL210 quartiles clearly showed that an increased COL210 score was associated with increased mortality (**Figure 2)**. The highest COL210 score strata displayed a HR of 1·76 [95% confidence interval (CI): 1·19 – 2·6; p<0·001; **Supplementary Table 4**]. Compared with the previously reported COV50 classifier^34^, COL210 better predicted the mortality than COV50 (HR 1·2; 95% CI: 1·17-1·24; p<0·001). Outcomes for short term (deaths within ICU, HR 3·85; 95% CI: 1·87-7·93; p<0·001) but also long-term mortality for patients that survived and exited ICU (HR 1·34, 95% CI: 0·76-2·35; p=0·3) are depicted (**Figures 3A and 3B**), with significant (p = 0·024) better prediction within the ICU.

**Figure 2:**
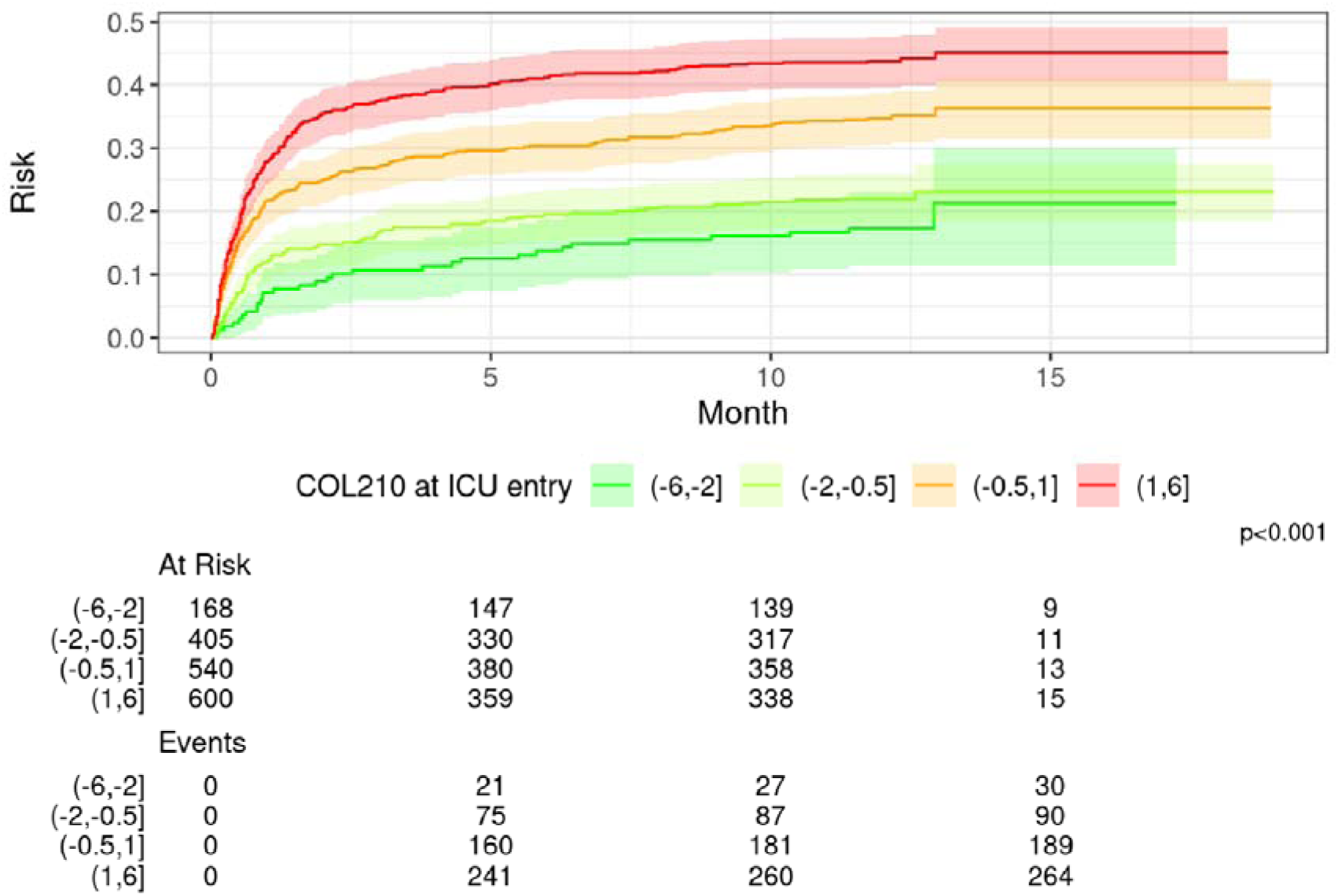
Survival curve depicting overall mortality in ICU group stratified into quartiles based on the COL210 score.

**Figure 3:**
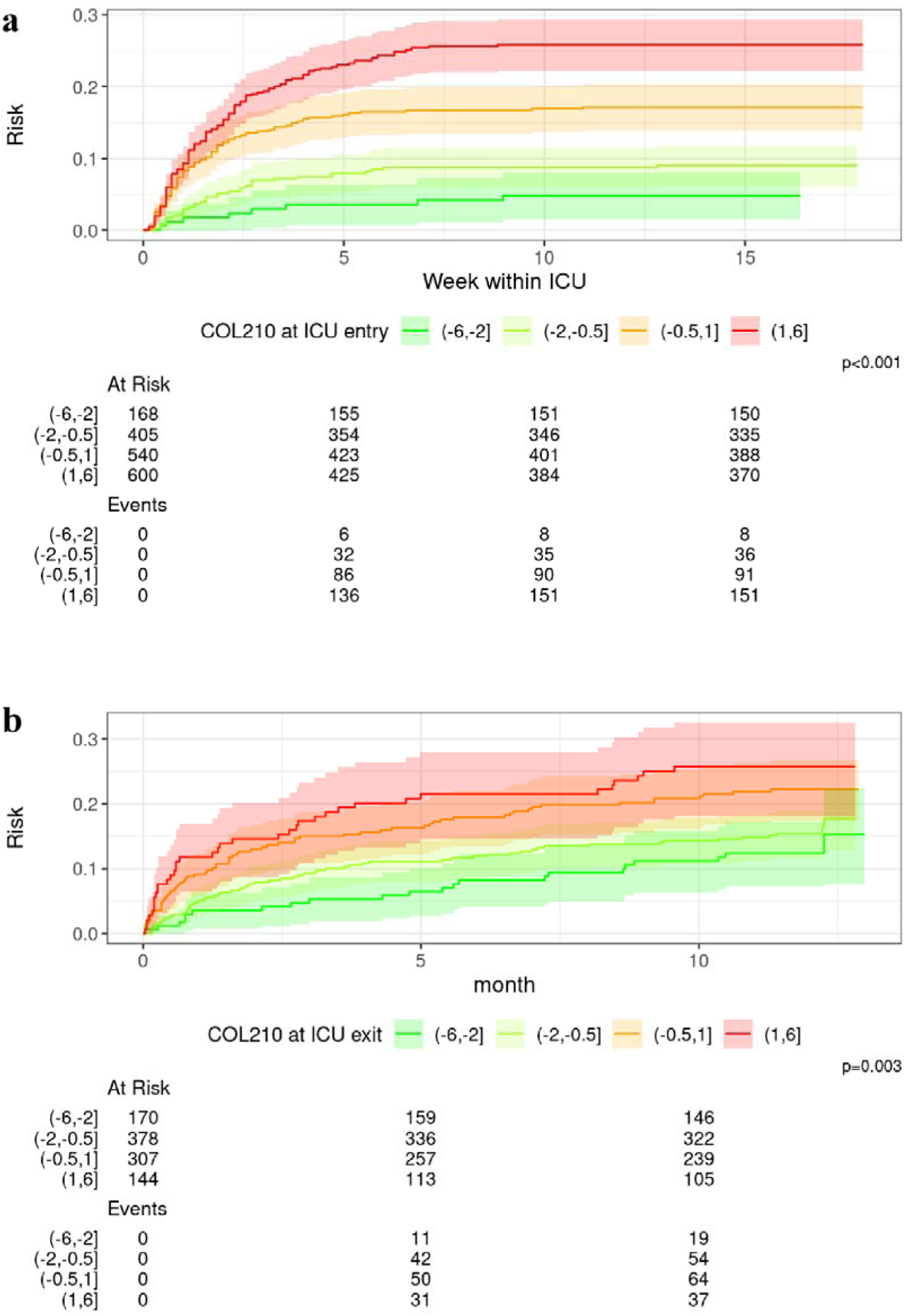
Survival curve depicting short-term, in ICU (a) and long-term, post-ICU mortality (b) in patients with critical condition stratified into quartiles based on the COL210 score.

### COL210 predictive value in non-critical patients (non-ICU)

To further expand our analysis, we subsequently investigated if COL210 is predictive of future death in subjects without a critical condition. The numbers of subjects within the non-ICU population per study split based on mortality events are summarised in **Supplementary Table 2**. Of the 7,474 subjects, 625 (8·4%) died during a median follow-up of 47 months. Survival curves depicting overall mortality within the non-ICU group stratified based on the COL210 score strata are shown in **Figure 4**, with adjusted HR estimated up to 2·20 for the highest COL210 score stratum (95% CI: 1·56 – 3·10; p<0·001; **Supplementary Table 5**). Therefore, in both ICU and non-ICU groups, higher COL210 scores are observed for patients that did not survive (**Figure 5A**), with a negative correlation of COL210 score with time to mortality (rho= -0·583, p<0·001 **Figure 5B**). As age is a crucial risk factor for death, we investigated the relationship between COL210 and mortality split by age groups (**Figure 6**). Evidently, mortality is increased with both age and COL210.

**Figure 4:**
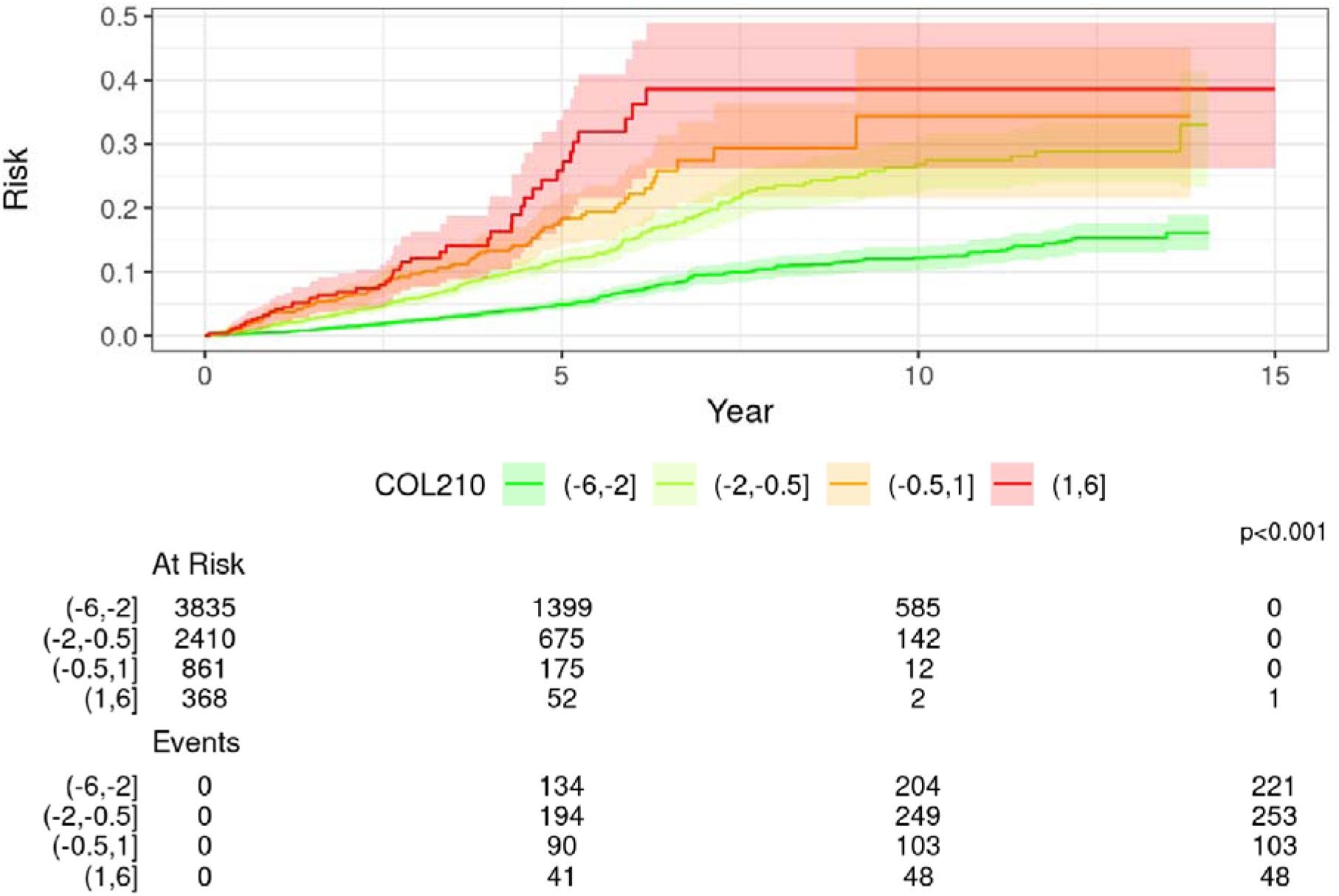
Survival curve depicting overall mortality in non-ICU groups stratified into quartiles based on the COL210 score.

**Figure 5:**
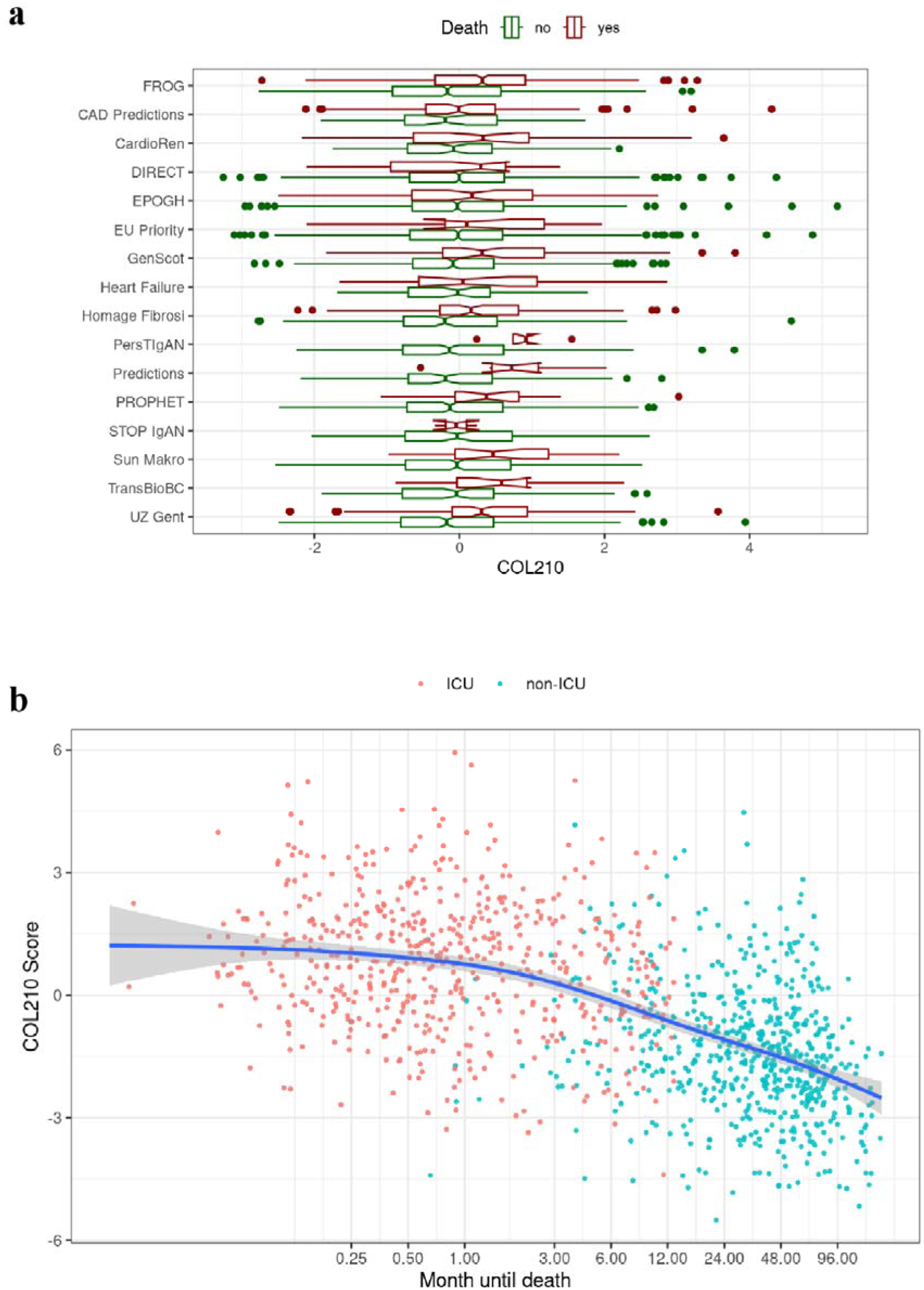
a) Distribution plots of the COL210 score within the different study cohorts. b) COL210 score correlation with the time to death.

**Figure 6:**
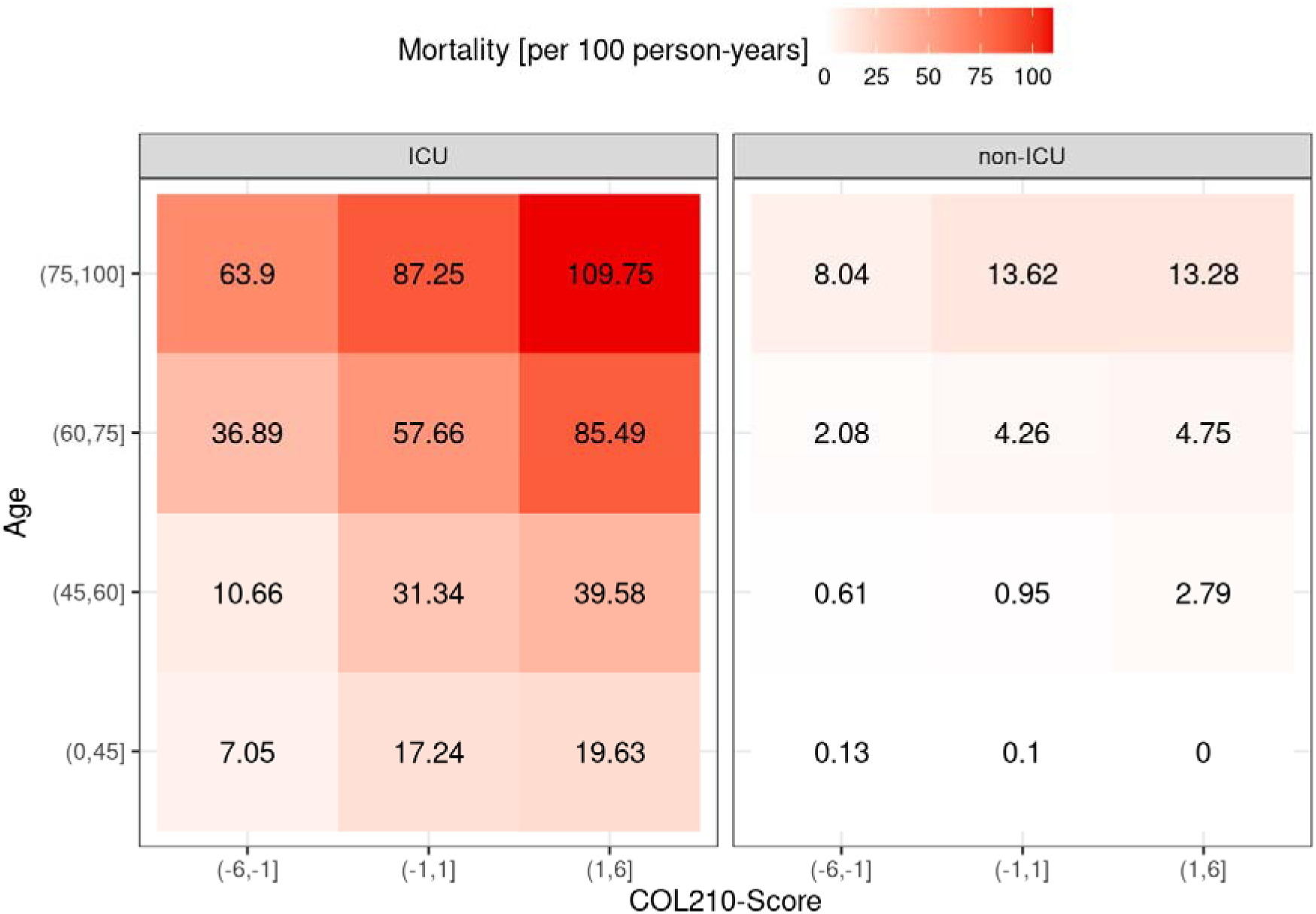
Mortality per person-years for ICU and non-ICU cohorts given age and COL210.

### Association of COL210 with Kidney Fibrosis

To investigate the association between COL210 and kidney fibrosis, datasets from 830 patients with CKD of various aetiologies were analysed. All patients had available data on the degree of fibrosis, assessed by IFTA. The COL210 classification scores were found to correlate significantly with IFTA, showing a Spearman correlation coefficient of rho = 0·253 (p < 0·0001), indicating statistically significant association between the COL210 classifier and the extent of kidney fibrosis.

## DISCUSSION

Recent organ-centric studies and clinical trial data are shifting the research focus from a static perspective of tissue imaging for assessing excessive collagen deposition to a more dynamic investigation of peripheral (serum or urine based) collagen markers. These non-invasive biomarkers can be indicative of fibrosis and further linked to organ failure and subsequent death. Importantly, this evolution is paralleled by a conceptual transition from investigating single diseases in isolation to a broader, integrative approach that identifies common molecular alterations across different organs and disease states. This allows for treatment strategies guided more by shared molecular signatures, such as collagen turnover, than by organ-specific pathophysiology alone. Within kidney fibrosis, multiple urinary collagen derived peptides, especially from COL1A1 and COL3A1, were found significantly associated with CKD progression.^64^ In subsequent studies these findings could be confirmed and expanded, demonstrating significant association of specific urine peptides with kidney fibrosis.^65^ In liver fibrosis, composite biomarker panels quantifying collagen synthesis, like combinations of PRO-C3, PRO-C5, and degradation markers such as C6M, show promise for predicting fibrosis progression, regression, and survival outcomes.^16^ In HF, markers of type I collagen turnover (e.g., urinary COL1A1 peptides) predict adverse cardiovascular events and mortality, demonstrating the clinical relevance of such non-invasive biomarkers for prognostication.^17, 20, 21^

In the context of the above literature, and following previous evidence from our group that collagen fragments naturally excreted in urine can be predictive of organ failure and death in organ-specific studies, in this study we investigated whether we can generalise these findings. Specifically, we hypothesised that an exclusively collagen-based model derived from CE-MS collagen peptides reads in urine are predictive of mortality in a large and diverse population of patients at a critical condition or following chronic diseases depicting a fibrotic environment. We were able to confirm the association of the collagen peptide-based classifier COL210 with the degree of kidney fibrosis in a large cohort of CKD patients.

The data presented are in agreement with multiple previous observations based on CE-MS-based analysis of urinary peptides. In the context of HF, He and colleagues^40^ reported significant reduction of the abundance of COL1A1 peptides from the central part of the collagen molecule associated with HF mortality, while several N- and C-terminal peptides were found increased. This is in line to our results supporting that COL1A1 central and relatively large peptides also associate with mortality. CKD273, a classifier based on 273 peptides significantly associated with CKD, which contains as a major component reduced abundance of several COL1A1 peptides, was also found associated with increased risk of mortality in several studies.^29, 31^ In addition, also in the COV-50 classifier, reduction of the abundance of specific COL1A1 peptides was found associated with mortality, both in patients with SARS-COV-2, but also in patients not infected with SARS-COV-2, further supporting the findings of this study. The COL210 peptide classifier improved the predictive value in comparison with previous peptide panels developed for detection of patients at critical state after SARS-CoV-2 infection.^32, 34^ Of the 34 collagen derived peptides contained in the COV-50 classifier, 20 overlap with the 210 collagen peptides identified here, all with identical trend in regulation. Six of these 20 also overlap with CKD273, further indicating commonality in the underlying biology.

The observation of reduced abundance of larger collagen I fragments associated with mortality is in line with a very recent publication (and data interpretation) from Mina et al.^36^ The authors aimed towards finding an explanation for the simultaneous increase and decrease of certain collagen fragments with CKD, a condition tightly associated with kidney fibrosis. Investigating the distribution of these peptides in detail, the authors presented a model suggesting initial degradation of collagen by endoproteases, likely members of the MMP family, resulting in the generation of larger peptides, which is attenuated in CKD. The same model is also applicable on the observations of this study, which suggests attenuation of collagen degradation, likely resulting in increased fibrosis, as associated with, and possible a significant contributor to increased risk of mortality.

Our data support the concept that intervention, e.g. via therapeutic drugs or lifestyle adjustments, addressing fibrosis may result as an additional benefit in the reduction of mortality. These thoughts are in general in agreement with the different effects observed in the context of sodium glucose cotransporter 2 (SGLT2) inhibition. This class of compounds, initially developed as antidiabetic drugs, has, in addition, demonstrated significant impact on mortality, and also on fibrosis. In a recent report^66^, we could also demonstrate significant increase of several urinary collagen fragments in patients after SGLT2 inhibitor treatment, indicating that these drugs, via inducing a metabolic switch, may result in reduction of highly reactive (oxygen) radicals, reducing collagen modification and crosslinking, and in this way enabling more efficient physiological collagen degradation, ideally even resolution of fibrosis. This hypothesis, although it has to be proven in additional experiments, would explain the above-mentioned observations.

The study presents with limitations, especially due to its retrospective design based on previously collected data. Nevertheless, the large number of datasets, the high number of endpoints assessed, and the high degree of homogeneity of the obtained findings strongly support that the results can be generalized. Along these lines, a strength of this analysis is the inclusion of datasets from multiple studies and across different conditions, indicating a robust basis for the assessment.

## CONCLUSIONS

Collectively, in this study we demonstrate that multiple specific urinary collagen peptides are significantly associated with future death in both patients at critical condition and those without. These peptides can be combined into a classifier, COL210 that enables the detection of “vulnerable” subjects, irrespective of the underlying conditions, of which score was also as correlated with the degree of fibrosis. This supports the rising clinical utility of collagen turnover biomarkers in predicting mortality across various fibrotic conditions and highlights the importance of dynamic markers assessing ECM. By integrating these biomarkers into risk models and leveraging advanced analytical methods like machine learning, predictive accuracy is enhanced. Further research is needed to assess if specific, personalized intervention guided by urinary collagen fragments can significantly improve outcomes and extend lifespan.

## Contributors

All authors read and approved the final version of the manuscript. Conceptualisation: MF, FK, HM, AV; Methodology: MF, FK, AL, HM, JS; Data generation: MF, FK, AL, LC, RW JS, HM; Visualisation: FK, JS, MF; Supervision: JB, AM, PP, JPS, JAS, AV, HM, JS; HR Writing—original draft: MF, JS; Writing—review & editing: MF, FK, AL, JB, AM, AC, DA, PP, JPS, JAS, HR,LC, RW, AV, HM, JS. MF, JS and FK accessed and verified the data.

## Data Availability

Anonymised data and code used in conducting the analyses will be made available by the corresponding author upon request.

## Acknowledgments

All authors are grateful to all patients who donated urine samples. This work was supported in part by funding through the European Union’s Horizon Europe Marie Skłodowska-Curie Actions Doctoral Networks - Industrial Doctorates Programme (HORIZON – MSCA – 2021 – DN-ID, “DisCo-I”, grant No 101072828) to AL, AV, JPS and HM. This study was also funded in part by “SIGNAL” (Federal Ministry of Education and Research, 01KU2307 to JS and HM; Austrian Science Fund (FWF) via Project number I 6464, Grant-DOI 10.55776/I6464 to FK and ANR-22-PERM-0002-06 to JPS), by “UriCov” (Federal Ministry of Health, 2523FSB114) to JS, HM, by “Accurate-CVD” (Federal Ministry for Economic Affairs and Climate Protection, ZIM-KK5560002AP3) to MF, HM, by UPTAKE project (Federal Ministry of Education and Research, 01EK2105A to HR and LC; 01EK2105B to HM and JS and 01EK2105C to RW and JB), and by MULTIR (HORIZON-MISS-2023-CANCER-01-01, project number: 101136926) to MF, AV funded by the European Commission. Views and opinions expressed are however those of the author(s) only and do not necessarily reflect those of the European Union. Neither the European Union nor the granting authorities HORIZON-MISS-2023-CANCER-01-01 and HORIZON – MSCA – 2021 – DN-ID can be held responsible for them.

## Supplementary Tables

**Supplementary Table 1:** Descriptive statistics for the ICU patients considered within this study.

**Supplementary Table 2:** Descriptive statistics for the non-ICU patients considered within this study.

**Supplementary Table 3:** Descriptive statistics for the CKD patients considered within this study.

**Supplementary Table 4.** List of 607 significant collagen peptides

**Supplementary Table 5.** Prediction of short-term mortality in patients with critical condition stratified based on the COL210 score range.

**Supplementary Table 6.** Prediction of mortality in patients with non-critical condition stratified based on underlying co-morbidities.

