## Supplementary tables for "Urinary collagen peptides predict mortality"

**Supplementary Table 1:** Descriptive statistics for the ICU patients considered within this study.

| | Level/Unit | Overall | Death: no | Death: yes | p |
|---|---|---|---|---|---|
| N | | 1719 (100%) | 1139 (66·3%) | 580 (33·7%) | |
| Study | FROG | 1719 (100%) | 1139 (100%) | 580 (100%) | |
| Age | [yrs] | 62 (50, 73) | 58 (46, 69) | 70 (61, 78) | <0·001 |
| Female | yes | 602 (35%) | 414 (36%) | 188 (32%) | 0·11 |
| BMI | [kg/m²] | 26·2 (22·9, 30·0) | 26·2 (22·8, 29·9) | 26·4 (23·1, 30·1) | 0·6 |
| Systolic BP | [mmHg] | 123 (109, 140) | 124 (110, 140) | 120 (107, 139) | 0·01 |
| Diastolic BP | [mmHg] | 64 (55, 75) | 66 (56, 77) | 60 (52, 70) | <0·001 |
| Mean Arterial BP | [mmHg] | 84 (74, 95) | 86 (76, 96) | 80 (71, 92) | <0·001 |
| Hypertension | yes | 979 (57%) | 602 (53%) | 377 (65%) | <0·001 |
| eGFR | [ml/min/1·73 m²] | 87 (48, 127) | 97 (57, 132) | 67 (37, 107) | <0·001 |
| Kidney Disease | yes | 716 (42%) | 378 (33%) | 338 (58%) | <0·001 |
| Diabetes | yes | 280 (16%) | 160 (14%) | 120 (21%) | <0·001 |
| Cardiovascular Disease | yes | 98 (5·7%) | 49 (4·3%) | 49 (8·4%) | <0·001 |
| COV50 | | 1·17 (0·34, 1·83) | 1·01 (0·11, 1·74) | 1·45 (0·72, 1·97) | <0·001 |
| FU Duration | [month] | 12·0 (2·0, 12·1) | 12·0 (12·0, 12·5) | 0·7 (0·3, 2·1) | <0·001 |

Categorical variables are described with absolute (N) and group-wise relative frequencies (%), continuous variables with median (IQR). P-values for group differences result from chi-squared homogeneity tests for categorical and for Wilcoxon rank sum test for continuous variables. Abbreviations: BMI= body mass index; BP= blood pressure; eGFR= estimated glomerular filtration rate; FU= follow-up; ICU= intensive care unit; yrs= years

**Supplementary Table 2:** Descriptive statistics for the Non-ICU patients considered within this study.

| | **Level/Unit** | **Overall** | **Death: no** | **Death: yes** | **p** |
|---|---|---|---|---|---|
| N | | 7474 (100%) | 6849 (91·6%) | 625 (8·4%) | |
| Study | | | | | <0·001 |
| | CAD Predictions | 145 (1·9%) | 50 (0·7%) | 95 (15%) | |
| | CardioRen | 116 (1·6%) | 87 (1·3%) | 29 (4·6%) | |
| | DIRECT | 1487 (20%) | 1448 (21%) | 39 (6·2%) | |
| | EPOGH | 914 (12%) | 850 (12%) | 64 (10%) | |
| | EU Priority | 1,769 (24%) | 1,756 (26%) | 13 (2·1%) | |
| | GenScot | 473 (6·3%) | 417 (6·1%) | 56 (9·0%) | |
| | Heart Failure | 84 (1·1%) | 67 (1·0%) | 17 (2·7%) | |
| | Homage Fibrosis | 354 (4·7%) | 229 (3·3%) | 125 (20%) | |
| | PersTIgAN | 270 (3·6%) | 265 (3·9%) | 5 (0·8%) | |
| | Predictions | 91 (1·2%) | 85 (1·2%) | 6 (1·0%) | |
| | PROPHET | 462 (6·2%) | 444 (6·5%) | 18 (2·9%) | |
| | STOP IgAN | 109 (1·5%) | 107 (1·6%) | 2 (0·3%) | |
| | Sun Makro | 581 (7·8%) | 556 (8·1%) | 25 (4·0%) | |
| | TransBioBC | 131 (1·8%) | 117 (1·7%) | 14 (2·2%) | |
| | UZ Gent | 488 (6·5%) | 371 (5·4%) | 117 (19%) | |
| Age | [yrs] | 60 (48, 68) | 59 (47, 66) | 73 (66, 79) | <0·001 |
| Female | yes | 2857 (38%) | 2657 (39%) | 200 (32%) | <0·001 |
| BMI | [kg/m²] | 27·5 (24·3, 31·2) | 27·6 (24·3, 31·4) | 26·9 (23·7, 30·1) | <0·001 |

| | Level/Unit | Overall | Death: no | Death: yes | p |
|---|---|---|---|---|---|
| Systolic BP | [mmHg] | 132 (121, 145) | 132 (121, 144) | 138 (124, 153) | <0·001 |
| Diastolic BP | [mmHg] | 79 (72, 85) | 79 (73, 85) | 75 (67, 82) | <0·001 |
| Mean Arterial BP | [mmHg] | 97 (90, 104) | 97 (90, 104) | 97 (88, 105) | 0·3 |
| Hypertension | yes | 3090 (41%) | 2758 (40%) | 332 (53%) | <0·001 |
| eGFR | [ml/min/1·73 m²] | 82 (59, 99) | 84 (62, 100) | 61 (37, 80) | <0·001 |
| Kidney Disease | yes | 2212 (30%) | 1898 (28%) | 314 (50%) | <0·001 |
| Diabetes | yes | 4101 (55%) | 3938 (57%) | 163 (26%) | <0·001 |
| Cardiovascular Disease | yes | 1357 (18%) | 983 (14%) | 374 (60%) | <0·001 |
| COV50 | | -1·88 (-2·33, -1·27) | -1·89 (-2·34, -1·30) | -1·69 (-2·26, -0·94) | <0·001 |
| FU Duration | [month] | 47 (29, 67) | 48 (29, 67) | 38 (19, 62) | <0·001 |

Categorical variables are described with absolute (N) and group-wise relative frequencies (%), continuous variables with median (IQR). P-values for group differences result from chi-squared homogeneity tests for categorical and for Wilcoxon rank sum test for continuous variables. Abbreviations: BMI= body mass index; BP= blood pressure; eGFR= estimated glomerular filtration rate; FU= follow-up; yrs= years

**Supplementary Table 3:** Descriptive statistics for the CKD patients considered within this study**.**

| | **Level/Unit** | **Overall** |
|---|---|---|
| N | | 830 (100%) |
| Age | [yrs] | 60 (47, 72) |
| Female | yes | 311 (37%) |
| BMI | [kg/m²] | 27·9 (24·3, 32·1) |
| Systolic BP | [mmHg] | 134 (120, 149) |
| Diastolic BP | [mmHg] | 79 (70, 84) |
| eGFR | [ml/min/1·73 m²] | 29 (14, 52) |
| IFTA | [%] | 20 (7·5, 35) |

Categorical variables are described with absolute (N) and group-wise relative frequencies (%), continuous variables with median (IQR). Abbreviations: BMI= body mass index; BP= blood pressure; eGFR= estimated glomerular filtration rate; yrs= years, IFTA=interstitial fibrosis and tubular atrophy

**Supplementary Table 4:** List of 607 significant collagen peptides.

| Peptide ID | Mass [Da] | CE-time [Min] | Sequence | UniprotID | Symbol | Protein name | p-value | adj_p-value (BH) | average_cases | average_controls | Fold change |
|---|---|---|---|---|---|---|---|---|---|---|---|
| e09627 | 1962·81 | 21·88 | PpGPpGKNGDDGEAGKpGRp | P02452 | COL1A1 | Collagen alpha-1(I) chain | 6·35E-08 | 1·58E-06 | 4·66 | 64·70 | 0·07 |
| e16641 | 3117·45 | 31·20 | GADGQPGAKGEpGDAGAKGDAGPPGPAGPAGPPGPIG | P02452 | COL1A1 | Collagen alpha-1(I) chain | 2·57E-03 | 1·26E-02 | 2·83 | 17·58 | 0·16 |
| e15230 | 2824·29 | 29·28 | AGpPGApGApGApGPVGPAGKSGDRGETGPAGP | P02452 | COL1A1 | Collagen alpha-1(I) chain | 1·44E-04 | 1·15E-03 | 22·25 | 130·71 | 0·17 |
| e13850 | 2587·19 | 21·13 | PpGKNGDDGEAGKPGRpGERGppGPQ | P02452 | COL1A1 | Collagen alpha-1(I) chain | 4·47E-09 | 1·74E-07 | 24·47 | 134·77 | 0·18 |
| e16966 | 3193·43 | 22·57 | PpGESGREGApGAEGSpGRDGSPGAKGDRGETGP | P02452 | COL1A1 | Collagen alpha-1(I) chain | 4·95E-17 | 1·13E-13 | 114·97 | 578·06 | 0·20 |
| e01399 | 1050·48 | 26·93 | DGRpGPpGPpG | P02452 | COL1A1 | Collagen alpha-1(I) chain | 4·38E-09 | 1·73E-07 | 55·58 | 247·84 | 0·22 |
| e18705 | 3660·71 | 33·06 | GADGQPGAKGEpGDAGAKGDAGPPGpAGPAGpPGPIGNVGApG | P02452 | COL1A1 | Collagen alpha-1(I) chain | 6·01E-05 | 5·48E-04 | 6·33 | 27·08 | 0·23 |
| e17162 | 3238·50 | 30·87 | GPAGFAGpPGADGQPGAKGEPGDAGAKGDAGPpGPAGP | P02452 | COL1A1 | Collagen alpha-1(I) chain | 3·66E-03 | 1·70E-02 | 6·22 | 26·17 | 0·24 |
| e05294 | 1474·65 | 20·07 | GRDGSpGAKGDRGET | P02452 | COL1A1 | Collagen alpha-1(I) chain | 1·08E-05 | 1·19E-04 | 10·98 | 38·79 | 0·28 |
| e17768 | 3381·56 | 31·78 | GAPGAEGSPGRDGSPGAKGDRGETGPAGPPGApGApGAp | P02452 | COL1A1 | Collagen alpha-1(I) chain | 7·12E-05 | 6·39E-04 | 12·79 | 41·05 | 0·31 |
| e17690 | 3359·58 | 31·68 | pPGADGQPGAKGEpGDAGAKGDAGppGPAGPAGPPGPIG | P02452 | COL1A1 | Collagen alpha-1(I) chain | 1·52E-15 | 1·16E-12 | 204·20 | 655·21 | 0·31 |
| e19431 | 3946·86 | 21·87 | SEGPQGVRGEPGppGpAGAAGPAGNPGADGQPGAKGAn-GAPGIAGA | P02452 | COL1A1 | Collagen alpha-1(I) chain | 8·33E-11 | 6·00E-09 | 87·67 | 265·38 | 0·33 |
| e02592 | 1186·53 | 22·31 | DDGEAGKpGRpG | P02452 | COL1A1 | Collagen alpha-1(I) chain | 1·94E-07 | 3·80E-06 | 71·82 | 213·21 | 0·34 |
| e00220 | 860·36 | 25·66 | DDGEAGKpG | P02452 | COL1A1 | Collagen alpha-1(I) chain | 5·93E-07 | 9·37E-06 | 63·16 | 182·97 | 0·35 |
| e16708 | 3133·47 | 31·24 | GADGQPGAKGEpGDAGAKGDAGPpGPAGPAGPPGPIG | P02452 | COL1A1 | Collagen alpha-1(I) chain | 1·54E-06 | 2·08E-05 | 34·33 | 96·86 | 0·35 |
| e17825 | 3400·59 | 32·15 | GPpGADGQPGAKGEPGDAGAKGDAGPPGpAGPAGPpGPIG | P02452 | COL1A1 | Collagen alpha-1(I) chain | 3·12E-09 | 1·26E-07 | 107·53 | 299·71 | 0·36 |
| e18006 | 3448·59 | 32·10 | PpGpAGFAGpPGADGQPGAKGEPGDAGAKGDAGPPGPAGP | P02452 | COL1A1 | Collagen alpha-1(I) chain | 2·63E-08 | 7·61E-07 | 75·88 | 208·89 | 0·36 |

| Pep-tide ID | Mass [Da] | CE-time [Min] | Sequence | UniprotID | Symbol | Protein name | p-value | adj_p-value (BH) | aver-age_cases | aver-age_con-trols | Fold change |
|---|---|---|---|---|---|---|---|---|---|---|---|
| e18580 | 3618·69 | 32·79 | FAGpPGADGQPGAKGEpGDAGAKGDAGpPGPAGPAGPPGPIG | P02452 | COL1A1 | Collagen alpha-1(I) chain | 3·53E-06 | 4·32E-05 | 43·60 | 119·33 | 0·37 |
| e17947 | 3433·55 | 31·83 | GSEGPQGVRGEPGpPGPAGAAGPAGNPGADGQPGAKGANG | P02452 | COL1A1 | Collagen alpha-1(I) chain | 2·32E-13 | 4·08E-11 | 322·54 | 865·92 | 0·37 |
| e17626 | 3343·57 | 31·67 | pPGADGQPGAKGEpGDAGAKGDAGPpGPAGPAGPPGPIG | P02452 | COL1A1 | Collagen alpha-1(I) chain | 3·67E-08 | 9·89E-07 | 47·96 | 127·74 | 0·38 |
| e16845 | 3165·47 | 31·32 | GADGQpGAKGEpGDAGAKGDAGPpGPAGPAGPpGPIG | P02452 | COL1A1 | Collagen alpha-1(I) chain | 5·38E-09 | 2·02E-07 | 102·48 | 253·52 | 0·40 |
| e16611 | 3109·44 | 36·22 | RGAPGDRGEPGPPGPAGFAGPpGADGQpGAKGEP | P02452 | COL1A1 | Collagen alpha-1(I) chain | 2·56E-05 | 2·62E-04 | 27·49 | 66·37 | 0·41 |
| e01983 | 1121·49 | 27·63 | DGRpGPpGPpGA | P02452 | COL1A1 | Collagen alpha-1(I) chain | 3·46E-03 | 1·63E-02 | 9·97 | 23·97 | 0·42 |
| e14876 | 2767·35 | 21·71 | KEGGKGPRGETGPAGRpGEVGPpGPpGPAG | P02452 | COL1A1 | Collagen alpha-1(I) chain | 2·12E-09 | 9·62E-08 | 100·21 | 238·41 | 0·42 |
| e08795 | 1865·80 | 32·98 | DAGPAGPKGEpGSpGENGApG | P02452 | COL1A1 | Collagen alpha-1(I) chain | 1·46E-04 | 1·16E-03 | 30·88 | 72·11 | 0·43 |
| e17047 | 3209·43 | 22·73 | PpGESGREGApGAEGSpGRDGSpGAKGDRGETGP | P02452 | COL1A1 | Collagen alpha-1(I) chain | 6·11E-14 | 1·75E-11 | 1363·50 | 3137·90 | 0·43 |
| e13830 | 2585·15 | 22·96 | RDGSPGAKGDRGETGPAGPPGApGApGAP | P02452 | COL1A1 | Collagen alpha-1(I) chain | 1·27E-02 | 4·55E-02 | 12·82 | 29·36 | 0·44 |
| e11537 | 2233·05 | 20·52 | GKNGDDGEAGKPGRpGERGPpGP | P02452 | COL1A1 | Collagen alpha-1(I) chain | 6·85E-08 | 1·65E-06 | 123·25 | 270·47 | 0·46 |
| e17949 | 3434·56 | 31·78 | GEPGPPGPAGFAGPPGADGQPGAKGEPGDAGAKGDAGPpG | P02452 | COL1A1 | Collagen alpha-1(I) chain | 2·63E-08 | 7·61E-07 | 221·26 | 466·93 | 0·47 |
| e05228 | 1466·64 | 28·47 | GPpGSpGEQGPSGASGP | P02452 | COL1A1 | Collagen alpha-1(I) chain | 1·18E-02 | 4·28E-02 | 18·78 | 39·21 | 0·48 |
| e05274 | 1471·63 | 22·10 | DDGEAGKpGRpGER | P02452 | COL1A1 | Collagen alpha-1(I) chain | 9·55E-10 | 4·86E-08 | 251·50 | 519·77 | 0·48 |
| e14445 | 2687·23 | 28·98 | KDGEAGAQGPPGPAGpAGERGEQGPAGSpG | P02452 | COL1A1 | Collagen alpha-1(I) chain | 2·37E-07 | 4·52E-06 | 53·30 | 108·81 | 0·49 |
| e16771 | 3149·46 | 31·28 | GADGQpGAKGEpGDAGAKGDAGPPGPAGPAGPpGPIG | P02452 | COL1A1 | Collagen alpha-1(I) chain | 2·14E-09 | 9·62E-08 | 191·94 | 388·98 | 0·49 |
| e18625 | 3633·74 | 27·04 | AAGEpGKAGERGVPGppGAVGPAGKDGEAGAQGpPGPAGPAG | P02452 | COL1A1 | Collagen alpha-1(I) chain | 1·19E-07 | 2·66E-06 | 83·71 | 164·91 | 0·51 |
| e18536 | 3603·69 | 32·96 | ADGQPGAKGEpGDAGAKGDAGPPGPAGpAGpPGPIGNVGApG | P02452 | COL1A1 | Collagen alpha-1(I) chain | 1·49E-03 | 7·97E-03 | 32·82 | 64·59 | 0·51 |
| e17946 | 3432·59 | 31·95 | PpGPAGFAGPPGADGQPGAKGEpGDAGAKGDAGPPGPAGP | P02452 | COL1A1 | Collagen alpha-1(I) chain | 8·68E-10 | 4·67E-08 | 744·52 | 1455·89 | 0·51 |

| Peptide ID | Mass [Da] | CE-time [Min] | Sequence | UniprotID | Symbol | Protein name | p-value | adj_p-value (BH) | average_cases | average_controls | Fold change |
|---|---|---|---|---|---|---|---|---|---|---|---|
| e18740 | 3671·71 | 22·41 | DKGETGEQGDRGIKGHRGFSGLQGppGPPGSPGEQGP | P02452 | COL1A1 | Collagen alpha-1(I) chain | 1·31E-02 | 4·63E-02 | 9·04 | 17·35 | 0·52 |
| e00467 | 911·44 | 25·83 | DGKTGPpGPA | P02452 | COL1A1 | Collagen alpha-1(I) chain | 2·80E-04 | 1·95E-03 | 161·29 | 309·30 | 0·52 |
| e12661 | 2377·11 | 20·84 | GKNGDDGEAGKPGRpGERGPpGpQ | P02452 | COL1A1 | Collagen alpha-1(I) chain | 2·47E-10 | 1·67E-08 | 582·46 | 1110·93 | 0·52 |
| e17221 | 3250·40 | 22·62 | GPpGESGREGAPGAEGSpGRDGSPGAKGDRGETGp | P02452 | COL1A1 | Collagen alpha-1(I) chain | 2·90E-06 | 3·69E-05 | 146·01 | 277·21 | 0·53 |
| e05379 | 1486·67 | 23·68 | DGSpGAKGDRGETGPA | P02452 | COL1A1 | Collagen alpha-1(I) chain | 1·91E-06 | 2·54E-05 | 182·18 | 345·37 | 0·53 |
| e16213 | 3022·36 | 24·59 | SGREGApGAEGSpGRDGSPGAKGDRGETGPAGP | P02452 | COL1A1 | Collagen alpha-1(I) chain | 2·75E-07 | 4·99E-06 | 95·56 | 179·02 | 0·53 |
| e17596 | 3337·47 | 22·75 | GPpGESGREGApGAEGSpGRDGSPGAKGDRGETGpA | P02452 | COL1A1 | Collagen alpha-1(I) chain | 2·80E-07 | 5·02E-06 | 251·38 | 469·49 | 0·54 |
| e18909 | 3736·69 | 32·21 | PAGKDGEAGAQGPpGpAGPAGERGEQGPAGSPGFQGLPGPAG | P02452 | COL1A1 | Collagen alpha-1(I) chain | 4·93E-06 | 5·83E-05 | 102·22 | 190·83 | 0·54 |
| e06176 | 1578·76 | 40·28 | VGPpGPpGPPGPpGPPSA | P02452 | COL1A1 | Collagen alpha-1(I) chain | 6·26E-03 | 2·61E-02 | 186·05 | 345·40 | 0·54 |
| e17348 | 3280·45 | 22·62 | ppGESGREGAPGAEGSpGRDGSpGAKGDRGETGPA | P02452 | COL1A1 | Collagen alpha-1(I) chain | 1·11E-12 | 1·70E-10 | 184·76 | 335·10 | 0·55 |
| e17890 | 3416·59 | 31·96 | GPpGADGQPGAKGEpGDAGAKGDAGPPGpAGPAGPPGpIG | P02452 | COL1A1 | Collagen alpha-1(I) chain | 1·44E-09 | 6·86E-08 | 686·62 | 1243·97 | 0·55 |
| e12211 | 2320·08 | 20·79 | KNGDDGEAGKpGRPGERGppGPQ | P02452 | COL1A1 | Collagen alpha-1(I) chain | 1·32E-04 | 1·08E-03 | 57·42 | 103·39 | 0·56 |
| e08755 | 1860·83 | 21·44 | EGSpGRDGSpGAKGDRGET | P02452 | COL1A1 | Collagen alpha-1(I) chain | 7·23E-07 | 1·10E-05 | 265·81 | 476·88 | 0·56 |
| e11903 | 2281·98 | 35·26 | GppGESGREGApGAEGSPGRDGSPG | P02452 | COL1A1 | Collagen alpha-1(I) chain | 7·43E-03 | 2·95E-02 | 93·05 | 163·41 | 0·57 |
| e16606 | 3108·46 | 31·19 | ADGQpGAKGEpGDAGAKGDAGpPGPAGPAGPpGPIG | P02452 | COL1A1 | Collagen alpha-1(I) chain | 5·91E-08 | 1·50E-06 | 293·56 | 515·33 | 0·57 |
| e16548 | 3092·47 | 31·13 | ADGQpGAKGEpGDAGAKGDAGPpGPAGPAGPPGPIG | P02452 | COL1A1 | Collagen alpha-1(I) chain | 1·86E-09 | 8·69E-08 | 273·68 | 476·01 | 0·57 |
| e16177 | 3013·34 | 22·23 | ESGREGApGAEGSPGRDGSpGAKGDRGETGpA | P02452 | COL1A1 | Collagen alpha-1(I) chain | 8·77E-10 | 4·67E-08 | 1755·75 | 3043·02 | 0·58 |
| e05423 | 1491·73 | 39·89 | VGpPGPPGpPGPPGPPS | P02452 | COL1A1 | Collagen alpha-1(I) chain | 6·64E-04 | 4·05E-03 | 178·91 | 308·44 | 0·58 |
| e05259 | 1469·67 | 23·62 | DGQPGAKGEpGDAGAK | P02452 | COL1A1 | Collagen alpha-1(I) chain | 1·26E-08 | 4·35E-07 | 734·20 | 1264·53 | 0·58 |

| Peptide ID | Mass [Da] | CE-time [Min] | Sequence | UniprotID | Symbol | Protein name | p-value | adj_p-value (BH) | average_cases | average_controls | Fold change |
|---|---|---|---|---|---|---|---|---|---|---|---|
| e18906 | 3735·70 | 32·37 | GERGVQGPpGpAGpRGANGAPGNDGAKGDAGAPGAPGSQGAPG | P02452 | COL1A1 | Collagen alpha-1(I) chain | 5·54E-05 | 5·09E-04 | 184·37 | 314·29 | 0·59 |
| e05369 | 1485·68 | 23·77 | DGQpGAKGEpGDAGAK | P02452 | COL1A1 | Collagen alpha-1(I) chain | 3·63E-06 | 4·40E-05 | 226·82 | 384·15 | 0·59 |
| e11527 | 2232·01 | 33·67 | IGPpGpAGApGDKGESGPSGPAGPTG | P02452 | COL1A1 | Collagen alpha-1(I) chain | 1·76E-04 | 1·33E-03 | 58·21 | 97·74 | 0·60 |
| e18578 | 3617·75 | 27·06 | AAGEpGKAGERGVpGPPGAVGPAGKDGEAGAQGPpGPAGPAG | P02452 | COL1A1 | Collagen alpha-1(I) chain | 1·95E-05 | 2·05E-04 | 36·11 | 59·15 | 0·61 |
| e03142 | 1247·52 | 22·07 | DKGETGEQGDRG | P02452 | COL1A1 | Collagen alpha-1(I) chain | 2·86E-07 | 5·09E-06 | 518·46 | 844·82 | 0·61 |
| e05136 | 1453·68 | 23·64 | DGQPGAKGEPGDAGAK | P02452 | COL1A1 | Collagen alpha-1(I) chain | 3·86E-03 | 1·78E-02 | 27·37 | 44·35 | 0·62 |
| e07275 | 1697·74 | 31·08 | NGApGNDGAKGDAGApGApG | P02452 | COL1A1 | Collagen alpha-1(I) chain | 7·73E-07 | 1·14E-05 | 423·82 | 685·09 | 0·62 |
| e05353 | 1483·66 | 22·63 | GPpGKNGDDGEAGKpG | P02452 | COL1A1 | Collagen alpha-1(I) chain | 5·08E-06 | 5·97E-05 | 211·02 | 340·27 | 0·62 |
| e13065 | 2446·09 | 28·29 | ADGQpGAKGEpGDAGAKGDAGPpGPAGP | P02452 | COL1A1 | Collagen alpha-1(I) chain | 1·82E-06 | 2·44E-05 | 133·12 | 213·21 | 0·62 |
| e14893 | 2770·25 | 29·27 | GPpGADGQpGAKGEpGDAGAKGDAGpPGPAGP | P02452 | COL1A1 | Collagen alpha-1(I) chain | 1·29E-02 | 4·57E-02 | 31·17 | 49·84 | 0·63 |
| e11002 | 2156·97 | 22·19 | AEGSpGRDGSpGAKGDRGETGPA | P02452 | COL1A1 | Collagen alpha-1(I) chain | 4·75E-07 | 7·72E-06 | 377·17 | 601·75 | 0·63 |
| e18627 | 3634·70 | 32·70 | FAGpPGADGQPGAKGEpGDAGAKGDAGPpGPAGPAGPpGPIG | P02452 | COL1A1 | Collagen alpha-1(I) chain | 3·43E-03 | 1·62E-02 | 53·96 | 84·66 | 0·64 |
| e16474 | 3076·46 | 31·08 | ADGQPGAKGEpGDAGAKGDAGpPGPAGPAGPPGPIG | P02452 | COL1A1 | Collagen alpha-1(I) chain | 2·04E-03 | 1·04E-02 | 27·57 | 43·00 | 0·64 |
| e17280 | 3264·57 | 25·70 | AAGEpGKAGERGVpGPpGAVGPAGKDGEAGAQGPPGP | P02452 | COL1A1 | Collagen alpha-1(I) chain | 2·66E-08 | 7·61E-07 | 733·37 | 1114·14 | 0·66 |
| e12949 | 2423·09 | 27·70 | LDGAKGDAGPAGpKGEpGSpGENGApG | P02452 | COL1A1 | Collagen alpha-1(I) chain | 8·04E-05 | 7·00E-04 | 145·89 | 221·55 | 0·66 |
| e16170 | 3011·39 | 29·69 | LTGSpGSpGpDGKTGPPGPAGQDGRPGPpGppG | P02452 | COL1A1 | Collagen alpha-1(I) chain | 9·75E-06 | 1·09E-04 | 1008·58 | 1493·95 | 0·68 |
| e07807 | 1750·79 | 23·87 | GpPGpPGKNGDDGEAGKpG | P02452 | COL1A1 | Collagen alpha-1(I) chain | 3·08E-05 | 3·08E-04 | 500·93 | 740·77 | 0·68 |
| e11777 | 2265·98 | 33·80 | ANGApGNDGAKGDAGApGApGSQGAPG | P02452 | COL1A1 | Collagen alpha-1(I) chain | 5·85E-04 | 3·65E-03 | 210·81 | 309·47 | 0·68 |
| e06309 | 1594·76 | 40·28 | VGPpGPpGPpGPpGPPSA | P02452 | COL1A1 | Collagen alpha-1(I) chain | 1·83E-04 | 1·36E-03 | 708·57 | 1032·95 | 0·69 |
| e11264 | 2194·95 | 33·38 | NGApGNDGAKGDAGAPGApGSQGApG | P02452 | COL1A1 | Collagen alpha-1(I) chain | 1·19E-03 | 6·57E-03 | 64·85 | 94·36 | 0·69 |

| Peptide ID | Mass [Da] | CE-time [Min] | Sequence | UniprotID | Symbol | Protein name | p-value | adj_p-value (BH) | average_cases | average_controls | Fold change |
|---|---|---|---|---|---|---|---|---|---|---|---|
| e10032 | 2014·89 | 21·88 | EGSpGRDGSpGAKGDRGETGP | P02452 | COL1A1 | Collagen alpha-1(I) chain | 1·22E-07 | 2·72E-06 | 1441·26 | 2086·25 | 0·69 |
| e11415 | 2216·03 | 33·83 | IGPpGPAGApGDKGESGPSGPAGPTG | P02452 | COL1A1 | Collagen alpha-1(I) chain | 1·21E-06 | 1·71E-05 | 323·80 | 467·94 | 0·69 |
| e13825 | 2584·24 | 35·08 | LTGPIGppGPAGAPGDKGESGPSGPAGPTG | P02452 | COL1A1 | Collagen alpha-1(I) chain | 3·14E-06 | 3·93E-05 | 363·10 | 510·26 | 0·71 |
| e05551 | 1507·73 | 40·19 | VGPpGPPGPpGPpGPPS | P02452 | COL1A1 | Collagen alpha-1(I) chain | 2·44E-03 | 1·20E-02 | 1416·21 | 1971·36 | 0·72 |
| e13277 | 2488·11 | 27·95 | PpGADGQpGAKGEpGDAGAKGDAGPpGP | P02452 | COL1A1 | Collagen alpha-1(I) chain | 6·21E-04 | 3·81E-03 | 81·22 | 112·67 | 0·72 |
| e05682 | 1523·73 | 40·32 | VGPpGPpGPpGPpGPPS | P02452 | COL1A1 | Collagen alpha-1(I) chain | 7·40E-03 | 2·95E-02 | 4233·04 | 5871·64 | 0·72 |
| e17505 | 3318·52 | 30·97 | AGRpGEVGPpGPpGPAGEKGSPGADGPAGAPGTpGPQG | P02452 | COL1A1 | Collagen alpha-1(I) chain | 1·08E-05 | 1·19E-04 | 154·75 | 213·52 | 0·72 |
| e03025 | 1235·55 | 26·86 | GQDGRpGPpGpPG | P02452 | COL1A1 | Collagen alpha-1(I) chain | 1·99E-03 | 1·02E-02 | 1011·25 | 1384·35 | 0·73 |
| e13342 | 2503·13 | 28·32 | GADGQpGAKGEpGDAGAKGDAGPpGPAGP | P02452 | COL1A1 | Collagen alpha-1(I) chain | 1·26E-02 | 4·51E-02 | 36·28 | 49·63 | 0·73 |
| e10483 | 2076·95 | 21·79 | GPpGPpGKNGDDGEAGKpGRpG | P02452 | COL1A1 | Collagen alpha-1(I) chain | 1·59E-04 | 1·23E-03 | 657·94 | 897·20 | 0·73 |
| e15778 | 2926·30 | 22·17 | ESGREGAPGAEGSPGRDGSpGAKGDRGETGp | P02452 | COL1A1 | Collagen alpha-1(I) chain | 3·61E-06 | 4·40E-05 | 274·52 | 369·30 | 0·74 |
| e15863 | 2942·30 | 22·19 | ESGREGApGAEGSpGRDGSpGAKGDRGETGP | P02452 | COL1A1 | Collagen alpha-1(I) chain | 3·66E-06 | 4·41E-05 | 1822·49 | 2448·32 | 0·74 |
| e11991 | 2294·03 | 28·29 | DGAKGDAGPAGPKGEpGSpGENGApG | P02452 | COL1A1 | Collagen alpha-1(I) chain | 4·26E-03 | 1·93E-02 | 26·39 | 35·30 | 0·75 |
| e15333 | 2841·26 | 24·45 | GAPGpQGFQGppGEPGEPGASGPMGPRGPPG | P02452 | COL1A1 | Collagen alpha-1(I) chain | 1·02E-05 | 1·14E-04 | 714·17 | 954·91 | 0·75 |
| e11375 | 2210·96 | 33·69 | NGApGNDGAKGDAGApGApGSQGApG | P02452 | COL1A1 | Collagen alpha-1(I) chain | 2·28E-04 | 1·64E-03 | 1916·39 | 2548·84 | 0·75 |
| e11213 | 2188·98 | 26·86 | ADGQPGAKGEpGDAGAKGDAGPPGP | P02452 | COL1A1 | Collagen alpha-1(I) chain | 1·49E-04 | 1·18E-03 | 497·40 | 646·73 | 0·77 |
| e12851 | 2407·09 | 27·69 | LDGAKGDAGPAGPKGEpGSpGENGApG | P02452 | COL1A1 | Collagen alpha-1(I) chain | 1·46E-04 | 1·16E-03 | 470·90 | 609·78 | 0·77 |
| e17438 | 3302·51 | 30·84 | AGRpGEVGpPGpPGPAGEKGSPGADGPAGAPGTPGPQG | P02452 | COL1A1 | Collagen alpha-1(I) chain | 2·10E-05 | 2·20E-04 | 210·63 | 272·02 | 0·77 |
| e01758 | 1096·48 | 26·17 | ApGDRGEpGPp | P02452 | COL1A1 | Collagen alpha-1(I) chain | 4·75E-03 | 2·11E-02 | 2288·46 | 2954·48 | 0·77 |
| e03890 | 1321·59 | 28·38 | ApGDRGEpGPpGPA | P02452 | COL1A1 | Collagen alpha-1(I) chain | 4·65E-03 | 2·08E-02 | 5422·22 | 6992·30 | 0·78 |

| Peptide ID | Mass [Da] | CE-time [Min] | Sequence | UniprotID | Symbol | Protein name | p-value | adj_p-value (BH) | average_cases | average_controls | Fold change |
|---|---|---|---|---|---|---|---|---|---|---|---|
| e19279 | 3871·78 | 27·55 | QGpRGSEGPQGVRGEPGPPGPAGAAGPAGNPGADGQP-GAKGANG | P02452 | COL1A1 | Collagen alpha-1(I) chain | 9·83E-03 | 3·69E-02 | 279·16 | 353·73 | 0·79 |
| e05844 | 1542·69 | 24·01 | DGQpGAKGEpGDAGAKG | P02452 | COL1A1 | Collagen alpha-1(I) chain | 7·03E-03 | 2·83E-02 | 28·18 | 35·70 | 0·79 |
| e10657 | 2101·91 | 22·00 | AEGSpGRDGSpGAKGDRGETGp | P02452 | COL1A1 | Collagen alpha-1(I) chain | 1·24E-06 | 1·73E-05 | 607·67 | 768·39 | 0·79 |
| e08320 | 1809·82 | 20·97 | GPpGKNGDDGEAGKpGRpG | P02452 | COL1A1 | Collagen alpha-1(I) chain | 1·34E-02 | 4·68E-02 | 68·41 | 86·19 | 0·79 |
| e17419 | 3295·53 | 25·47 | KGESGPSGPAGPTGARGAPGDRGEPGPpGpAGFAGPP | P02452 | COL1A1 | Collagen alpha-1(I) chain | 3·64E-05 | 3·55E-04 | 195·95 | 246·77 | 0·79 |
| e10266 | 2047·92 | 21·93 | NGDDGEAGKpGRpGERGPPGP | P02452 | COL1A1 | Collagen alpha-1(I) chain | 7·51E-04 | 4·48E-03 | 7907·75 | 9659·54 | 0·82 |
| e12366 | 2339·00 | 34·00 | GANGApGNDGAKGDAGApGApGSQGApG | P02452 | COL1A1 | Collagen alpha-1(I) chain | 7·05E-03 | 2·83E-02 | 325·19 | 396·47 | 0·82 |
| e11536 | 2233·01 | 22·45 | NGDDGEAGKPGRpGERGPpGPQG | P02452 | COL1A1 | Collagen alpha-1(I) chain | 1·66E-04 | 1·26E-03 | 313·53 | 381·90 | 0·82 |
| e07823 | 1752·78 | 19·61 | PpGKNGDDGEAGKpGRpG | P02452 | COL1A1 | Collagen alpha-1(I) chain | 4·78E-04 | 3·10E-03 | 59·01 | 71·78 | 0·82 |
| e10554 | 2085·93 | 21·99 | EGSpGRDGSpGAKGDRGETGPA | P02452 | COL1A1 | Collagen alpha-1(I) chain | 1·00E-03 | 5·68E-03 | 5389·29 | 6529·83 | 0·83 |
| e14211 | 2644·23 | 21·20 | GPpGKNGDDGEAGKPGRpGERGPpGpQ | P02452 | COL1A1 | Collagen alpha-1(I) chain | 9·53E-03 | 3·59E-02 | 308·23 | 370·94 | 0·83 |
| e19885 | 4169·93 | 33·60 | ERGEQGPAGSpGFQGLpGpAGppGEAGKpGEQGVPGDLGAPGPSG | P02452 | COL1A1 | Collagen alpha-1(I) chain | 1·08E-02 | 4·00E-02 | 1537·09 | 1842·68 | 0·83 |
| e09408 | 1933·88 | 21·64 | GDDGEAGKpGRPGERGPPGp | P02452 | COL1A1 | Collagen alpha-1(I) chain | 6·73E-04 | 4·09E-03 | 745·75 | 882·75 | 0·84 |
| e02530 | 1179·52 | 26·93 | pGDRGEpGPpGP | P02452 | COL1A1 | Collagen alpha-1(I) chain | 7·61E-03 | 3·00E-02 | 207·35 | 242·15 | 0·86 |
| e03723 | 1306·56 | 27·33 | GQDGRPGppGpPGA | P02452 | COL1A1 | Collagen alpha-1(I) chain | 4·39E-03 | 1·97E-02 | 1216·91 | 1417·79 | 0·86 |
| e05651 | 1519·69 | 22·58 | GQDGRPGppGpPGARG | P02452 | COL1A1 | Collagen alpha-1(I) chain | 1·66E-03 | 8·74E-03 | 101·26 | 117·81 | 0·86 |
| e18806 | 3696·77 | 27·01 | ARGNDGATGAAGPpGPTGPAGpPGFpGAVGAKGEAGpQGPRG | P02452 | COL1A1 | Collagen alpha-1(I) chain | 1·58E-05 | 1·68E-04 | 195·59 | 225·58 | 0·87 |
| e17381 | 3287·49 | 30·80 | GpAGpRGANGApGNDGAKGDAGAPGAPGSQGAPGLQGM | P02452 | COL1A1 | Collagen alpha-1(I) chain | 1·44E-02 | 4·96E-02 | 2321·29 | 2662·85 | 0·87 |
| e13322 | 2500·18 | 20·94 | GPpGKNGDDGEAGKpGRpGERGPPGP | P02452 | COL1A1 | Collagen alpha-1(I) chain | 1·31E-02 | 4·63E-02 | 81·78 | 92·36 | 0·89 |
| e08916 | 1876·87 | 22·17 | DDGEAGKPGRpGERGpPGP | P02452 | COL1A1 | Collagen alpha-1(I) chain | 6·24E-03 | 2·61E-02 | 1213·24 | 1352·87 | 0·90 |

| Peptide ID | Mass [Da] | CE-time [Min] | Sequence | UniprotID | Symbol | Protein name | p-value | adj_p-value (BH) | average_cases | average_controls | Fold change |
|---|---|---|---|---|---|---|---|---|---|---|---|
| e17288 | 3266·43 | 22·62 | GPpGESGREGAPGAEGSpGRDGSpGAKGDRGETGp | P02452 | COL1A1 | Collagen alpha-1(I) chain | 1·11E-02 | 4·08E-02 | 2632·14 | 2933·76 | 0·90 |
| e13200 | 2471·16 | 34·76 | TGPIGpPGPAGAPGDKGESGpSGPAGPTG | P02452 | COL1A1 | Collagen alpha-1(I) chain | 1·10E-02 | 4·06E-02 | 211·02 | 233·05 | 0·91 |
| e10883 | 2137·99 | 25·72 | NSGEPGApGSKGDTGAKGEPGPVG | P02452 | COL1A1 | Collagen alpha-1(I) chain | 6·67E-03 | 2·74E-02 | 103·96 | 112·69 | 0·92 |
| e11193 | 2185·99 | 25·94 | NSGEpGApGSKGDTGAkGEpGPVG | P02452 | COL1A1 | Collagen alpha-1(I) chain | 1·86E-03 | 9·64E-03 | 1181·59 | 1270·81 | 0·93 |
| e11128 | 2176·03 | 20·48 | KNGDDGEAGKPGRPGERGPpGp | P02452 | COL1A1 | Collagen alpha-1(I) chain | 1·41E-03 | 7·59E-03 | 19·45 | 20·87 | 0·93 |
| e09059 | 1892·87 | 22·13 | DDGEAGKPGRpGERGpPGp | P02452 | COL1A1 | Collagen alpha-1(I) chain | 5·41E-03 | 2·33E-02 | 318·33 | 335·20 | 0·95 |
| e09542 | 1949·89 | 21·61 | GDDGEAGKpGRpGERGpPGP | P02452 | COL1A1 | Collagen alpha-1(I) chain | 8·04E-04 | 4·71E-03 | 250·26 | 243·76 | 1·03 |
| e03533 | 1286·54 | 29·33 | DGQpGAKGEpGDAG | P02452 | COL1A1 | Collagen alpha-1(I) chain | 1·23E-02 | 4·41E-02 | 73·35 | 70·23 | 1·04 |
| e12101 | 2308·01 | 27·33 | ADGQpGAKGEpGDAGAKGDAGPpGpA | P02452 | COL1A1 | Collagen alpha-1(I) chain | 2·01E-03 | 1·03E-02 | 166·78 | 156·59 | 1·07 |
| e09125 | 1899·87 | 21·34 | SpGRDGSpGAKGDRGETGPA | P02452 | COL1A1 | Collagen alpha-1(I) chain | 6·47E-03 | 2·66E-02 | 190·40 | 173·38 | 1·10 |
| e01511 | 1065·50 | 25·57 | GPDGKTGpPGPA | P02452 | COL1A1 | Collagen alpha-1(I) chain | 4·78E-03 | 2·12E-02 | 56·48 | 51·30 | 1·10 |
| e11940 | 2287·01 | 22·41 | GESGREGApGAEGSpGRDGSpGAKG | P02452 | COL1A1 | Collagen alpha-1(I) chain | 1·43E-02 | 4·95E-02 | 43·01 | 37·59 | 1·14 |
| e14190 | 2639·29 | 21·44 | KEGGKGPRGETGPAGRpGEVGPpGPpGP | P02452 | COL1A1 | Collagen alpha-1(I) chain | 1·07E-05 | 1·19E-04 | 389·91 | 322·90 | 1·21 |
| e14501 | 2697·23 | 29·13 | pPGADGQPGAKGEpGDAGAKGDAGpPGPAGP | P02452 | COL1A1 | Collagen alpha-1(I) chain | 1·17E-02 | 4·25E-02 | 111·98 | 89·15 | 1·26 |
| e06697 | 1635·79 | 40·52 | VGPpGpPGPpGPPGPPSAG | P02452 | COL1A1 | Collagen alpha-1(I) chain | 1·20E-02 | 4·34E-02 | 704·95 | 535·61 | 1·32 |
| e05705 | 1526·69 | 23·89 | DGQPGAKGEpGDAGAKG | P02452 | COL1A1 | Collagen alpha-1(I) chain | 1·38E-02 | 4·79E-02 | 73·55 | 55·57 | 1·32 |
| e13223 | 2476·17 | 22·83 | GSpGPAGPKGSpGEAGRpGEAGLpGAKG | P02452 | COL1A1 | Collagen alpha-1(I) chain | 4·39E-03 | 1·97E-02 | 79·77 | 60·26 | 1·32 |
| e06859 | 1651·79 | 40·75 | VGpPGpPGPpGPpGPPSAG | P02452 | COL1A1 | Collagen alpha-1(I) chain | 2·80E-03 | 1·36E-02 | 2431·82 | 1745·61 | 1·39 |
| e17213 | 3248·56 | 25·78 | AAGEPGKAGERGVpGpPGAVGPAGKDGEAGAQGPPGP | P02452 | COL1A1 | Collagen alpha-1(I) chain | 2·84E-03 | 1·37E-02 | 914·33 | 627·05 | 1·46 |
| e14583 | 2713·23 | 29·12 | pPGADGQpGAKGEpGDAGAKGDAGPpGPAGP | P02452 | COL1A1 | Collagen alpha-1(I) chain | 4·18E-03 | 1·90E-02 | 44·46 | 30·02 | 1·48 |

| Peptide ID | Mass [Da] | CE-time [Min] | Sequence | UniprotID | Symbol | Protein name | p-value | adj_p-value (BH) | average_cases | average_controls | Fold change |
|---|---|---|---|---|---|---|---|---|---|---|---|
| e20139 | 4343·06 | 25·14 | VGPpGPpGpPGPPGPPSAGFDFSFLPQPPQEKAHDGGRYYRA | P02452 | COL1A1 | Collagen alpha-1(I) chain | 5·38E-04 | 3·41E-03 | 37·91 | 25·28 | 1·50 |
| e19111 | 3802·73 | 32·21 | ANGApGNDGAKGDAGApGApGSQGApGLQGMpGERGAAGLPGp | P02452 | COL1A1 | Collagen alpha-1(I) chain | 1·38E-04 | 1·12E-03 | 1200·80 | 761·83 | 1·58 |
| e16895 | 3177·44 | 22·62 | pPGESGREGAPGAEGSPGRDGSpGAKGDRGETGP | P02452 | COL1A1 | Collagen alpha-1(I) chain | 3·43E-07 | 5·86E-06 | 99·56 | 61·94 | 1·61 |
| e12488 | 2352·04 | 26·81 | KGDRGETGpAGPPGApGAPGAPGPVGP | P02452 | COL1A1 | Collagen alpha-1(I) chain | 3·72E-03 | 1·73E-02 | 1093·24 | 650·23 | 1·68 |
| e05174 | 1458·62 | 28·04 | SpGENGApGQmGPRG | P02452 | COL1A1 | Collagen alpha-1(I) chain | 1·15E-02 | 4·17E-02 | 294·18 | 169·23 | 1·74 |
| e13817 | 2583·20 | 28·30 | AGPpGAPGApGAPGpVGPAGKSGDRGETGP | P02452 | COL1A1 | Collagen alpha-1(I) chain | 5·08E-04 | 3·26E-03 | 1167·30 | 657·32 | 1·78 |
| e10135 | 2029·94 | 21·76 | SGNAGPpGPpGPAGKEGGKGPRG | P02452 | COL1A1 | Collagen alpha-1(I) chain | 1·22E-02 | 4·40E-02 | 391·15 | 219·18 | 1·78 |
| e18791 | 3689·76 | 20·75 | LQGMpGERGAAGLPGpKGDRGDAGPKGADGSpGKDGVRG | P02452 | COL1A1 | Collagen alpha-1(I) chain | 3·99E-05 | 3·84E-04 | 114·38 | 62·66 | 1·83 |
| e19228 | 3847·85 | 27·44 | PGLPGPSGEpGKQGpSGASGERGPPGPMGPPGLAGppGESGR | P02452 | COL1A1 | Collagen alpha-1(I) chain | 3·07E-06 | 3·88E-05 | 308·03 | 149·75 | 2·06 |
| e04405 | 1376·68 | 20·00 | PpGPAGKEGGKGPRG | P02452 | COL1A1 | Collagen alpha-1(I) chain | 2·45E-03 | 1·21E-02 | 1060·92 | 502·87 | 2·11 |
| e10002 | 2009·96 | 24·86 | GPSGEpGKQGpSGASGERGpPG | P02452 | COL1A1 | Collagen alpha-1(I) chain | 6·67E-06 | 7·76E-05 | 349·68 | 162·35 | 2·15 |
| e16638 | 3115·45 | 25·06 | QGPGGPpGPKGNSGEpGApGSKGDTGAKGEPGpVG | P02452 | COL1A1 | Collagen alpha-1(I) chain | 1·23E-04 | 1·01E-03 | 113·52 | 50·84 | 2·23 |
| e09923 | 1997·91 | 25·16 | NSGEPGApGSKGDTGAKGEpGP | P02452 | COL1A1 | Collagen alpha-1(I) chain | 8·99E-03 | 3·45E-02 | 581·27 | 259·73 | 2·24 |
| e03111 | 1244·56 | 21·70 | GRDGSpGAKGDRG | P02452 | COL1A1 | Collagen alpha-1(I) chain | 2·32E-03 | 1·15E-02 | 57·78 | 23·35 | 2·47 |
| e18831 | 3703·71 | 22·41 | DKGETGEQGDRGIKGHRGFSGLQGppGppGSPGEQGP | P02452 | COL1A1 | Collagen alpha-1(I) chain | 1·40E-04 | 1·13E-03 | 3532·74 | 1418·12 | 2·49 |
| e16605 | 3108·44 | 20·64 | RGPpGPpGKNGDDGEAGKPGRpGERGPpGPQG | P02452 | COL1A1 | Collagen alpha-1(I) chain | 5·48E-03 | 2·36E-02 | 87·39 | 33·45 | 2·61 |
| e16036 | 2979·40 | 29·96 | LTGSpGSpGPDGKTGPpGPAGQDGRPGPPGPpG | P02452 | COL1A1 | Collagen alpha-1(I) chain | 5·51E-03 | 2·36E-02 | 496·79 | 185·36 | 2·68 |
| e15755 | 2923·38 | 20·41 | RGPpGPpGKNGDDGEAGKPGRpGERGPpGP | P02452 | COL1A1 | Collagen alpha-1(I) chain | 7·12E-10 | 4·37E-08 | 3317·48 | 1126·56 | 2·94 |
| e15015 | 2790·39 | 20·22 | PGPAGPPGEAGKPGEQGVPGDLGAPGPSGARG | P02452 | COL1A1 | Collagen alpha-1(I) chain | 2·67E-04 | 1·87E-03 | 3276·63 | 1102·30 | 2·97 |

| Peptide ID | Mass [Da] | CE-time [Min] | Sequence | UniprotID | Symbol | Protein name | p-value | adj_p-value (BH) | average_cases | average_controls | Fold change |
|---|---|---|---|---|---|---|---|---|---|---|---|
| e09308 | 1921·85 | 31·98 | SAGPpGATGFpGAAGRVGPpGP | P02452 | COL1A1 | Collagen alpha-1(I) chain | 2·94E-04 | 2·03E-03 | 142·09 | 47·19 | 3·01 |
| e05480 | 1498·66 | 29·13 | GEAGKpGEQGVpGDLG | P02452 | COL1A1 | Collagen alpha-1(I) chain | 1·55E-03 | 8·24E-03 | 327·37 | 107·23 | 3·05 |
| e17471 | 3310·44 | 25·12 | AGAPGDKGESGPSGPAGpTGARGAPGDRGEpGPpGPAG | P02452 | COL1A1 | Collagen alpha-1(I) chain | 2·49E-04 | 1·76E-03 | 237·24 | 74·42 | 3·19 |
| e12904 | 2414·15 | 22·94 | SGNAGPpGPPGpAGKEGGKGPRGETGP | P02452 | COL1A1 | Collagen alpha-1(I) chain | 2·90E-07 | 5·11E-06 | 738·47 | 228·65 | 3·23 |
| e11146 | 2178·95 | 32·66 | GEPGSpGENGApGQMGPRGLPGE | P02452 | COL1A1 | Collagen alpha-1(I) chain | 5·54E-04 | 3·48E-03 | 154·69 | 47·19 | 3·28 |
| e12542 | 2359·07 | 27·50 | DGQpGAKGEPGDAGAKGDAGPpGPAGP | P02452 | COL1A1 | Collagen alpha-1(I) chain | 4·95E-11 | 4·05E-09 | 292·32 | 88·48 | 3·30 |
| e04274 | 1361·58 | 27·83 | NSGEpGApGSKGDTG | P02452 | COL1A1 | Collagen alpha-1(I) chain | 5·17E-03 | 2·25E-02 | 46·28 | 13·51 | 3·42 |
| e14887 | 2769·31 | 20·18 | RGPpGPpGKNGDDGEAGKpGRpGERGPP | P02452 | COL1A1 | Collagen alpha-1(I) chain | 9·69E-06 | 1·09E-04 | 151·22 | 44·09 | 3·43 |
| e00191 | 854·41 | 23·29 | PpGERGGpG | P02452 | COL1A1 | Collagen alpha-1(I) chain | 1·34E-02 | 4·68E-02 | 111·04 | 31·39 | 3·54 |
| e14935 | 2776·31 | 24·06 | GPpGPKGNSGEpGApGSKGDTGAKGEpGPVG | P02452 | COL1A1 | Collagen alpha-1(I) chain | 1·46E-04 | 1·16E-03 | 253·08 | 70·42 | 3·59 |
| e13118 | 2456·12 | 27·82 | PpGADGQPGAKGEpGDAGAKGDAGPPGP | P02452 | COL1A1 | Collagen alpha-1(I) chain | 8·07E-03 | 3·15E-02 | 91·31 | 24·02 | 3·80 |
| e02424 | 1168·54 | 26·34 | KpGEQGVPGDLG | P02452 | COL1A1 | Collagen alpha-1(I) chain | 4·69E-08 | 1·24E-06 | 296·28 | 72·96 | 4·06 |
| e05160 | 1456·70 | 22·56 | ApGSKGDTGAKGEpGP | P02452 | COL1A1 | Collagen alpha-1(I) chain | 1·20E-04 | 9·93E-04 | 241·16 | 57·26 | 4·21 |
| e11918 | 2284·02 | 33·27 | DGAKGDAGAPGApGSQGApGLQGmpG | P02452 | COL1A1 | Collagen alpha-1(I) chain | 1·69E-04 | 1·28E-03 | 265·44 | 62·72 | 4·23 |
| e14827 | 2759·34 | 23·79 | GFPGpKGAAGEpGKAGERGVpGPpGAVGPAG | P02452 | COL1A1 | Collagen alpha-1(I) chain | 7·95E-04 | 4·68E-03 | 68·54 | 16·16 | 4·24 |
| e06870 | 1652·76 | 30·26 | GSpGSpGPDGKTGppGPAG | P02452 | COL1A1 | Collagen alpha-1(I) chain | 5·69E-03 | 2·41E-02 | 392·90 | 81·08 | 4·85 |
| e07514 | 1721·81 | 23·71 | ApGPVGPAGKSGDRGETGP | P02452 | COL1A1 | Collagen alpha-1(I) chain | 6·15E-07 | 9·59E-06 | 151·58 | 30·87 | 4·91 |
| e05202 | 1462·68 | 22·27 | DGRpGPpGPpGARGQ | P02452 | COL1A1 | Collagen alpha-1(I) chain | 1·31E-06 | 1·82E-05 | 995·55 | 200·12 | 4·97 |
| e05309 | 1476·67 | 29·51 | TGPIGPpGPAGApGDKG | P02452 | COL1A1 | Collagen alpha-1(I) chain | 7·48E-03 | 2·96E-02 | 352·46 | 70·68 | 4·99 |
| e05509 | 1501·73 | 20·17 | GSpGRDGSpGAKGDRG | P02452 | COL1A1 | Collagen alpha-1(I) chain | 7·61E-07 | 1·14E-05 | 2756·49 | 535·52 | 5·15 |
| e14709 | 2738·23 | 28·44 | GPpGADGQPGAKGEPGDAGAKGDAGPpGPAGP | P02452 | COL1A1 | Collagen alpha-1(I) chain | 1·50E-03 | 8·02E-03 | 149·39 | 28·98 | 5·15 |

| Peptide ID | Mass [Da] | CE-time [Min] | Sequence | UniprotID | Symbol | Protein name | p-value | adj_p-value (BH) | average_cases | average_controls | Fold change |
|---|---|---|---|---|---|---|---|---|---|---|---|
| e06054 | 1566·70 | 29·50 | GpAGNpGADGQPGAKGAN | P02452 | COL1A1 | Collagen alpha-1(I) chain | 6·36E-03 | 2·63E-02 | 277·54 | 51·51 | 5·39 |
| e02069 | 1130·49 | 27·54 | KGEpGSpGENGA | P02452 | COL1A1 | Collagen alpha-1(I) chain | 9·25E-03 | 3·51E-02 | 50·65 | 9·36 | 5·41 |
| e15006 | 2788·28 | 21·65 | ESGREGApGAEGSpGRDGSpGAKGDRGET | P02452 | COL1A1 | Collagen alpha-1(I) chain | 6·69E-03 | 2·74E-02 | 54·04 | 9·98 | 5·42 |
| e00992 | 993·48 | 24·84 | VRGEpGPpGP | P02452 | COL1A1 | Collagen alpha-1(I) chain | 1·13E-02 | 4·14E-02 | 58·67 | 10·63 | 5·52 |
| e09562 | 1952·86 | 25·43 | GEPGApGSKGDTGAKGEpGPVG | P02452 | COL1A1 | Collagen alpha-1(I) chain | 4·69E-03 | 2·09E-02 | 173·41 | 30·41 | 5·70 |
| e09799 | 1981·94 | 25·16 | NSGEPGApGSKGDTGAKGEPGP | P02452 | COL1A1 | Collagen alpha-1(I) chain | 3·92E-16 | 4·49E-13 | 1351·14 | 229·98 | 5·88 |
| e08392 | 1816·88 | 24·38 | SGNAGPpGPpGPAGKEGGKGP | P02452 | COL1A1 | Collagen alpha-1(I) chain | 7·19E-07 | 1·10E-05 | 296·73 | 49·79 | 5·96 |
| e01025 | 998·48 | 21·21 | DGEAGKpGRP | P02452 | COL1A1 | Collagen alpha-1(I) chain | 1·87E-03 | 9·69E-03 | 31·03 | 5·12 | 6·06 |
| e09953 | 2002·94 | 25·44 | GQPGAKGEpGDAGAKGDAGPPGP | P02452 | COL1A1 | Collagen alpha-1(I) chain | 5·43E-04 | 3·43E-03 | 101·08 | 16·36 | 6·18 |
| e08234 | 1798·88 | 41·05 | VGPpGPpGPpGPpGPPSAGF | P02452 | COL1A1 | Collagen alpha-1(I) chain | 4·53E-07 | 7·52E-06 | 1038·75 | 164·46 | 6·32 |
| e13730 | 2567·20 | 28·23 | AGPpGAPGApGAPGPVGPAGKSGDRGETGP | P02452 | COL1A1 | Collagen alpha-1(I) chain | 3·37E-07 | 5·84E-06 | 2406·05 | 380·52 | 6·32 |
| e13648 | 2554·14 | 23·13 | GPpGESGREGApGAEGSpGRDGSpGAKG | P02452 | COL1A1 | Collagen alpha-1(I) chain | 3·83E-03 | 1·77E-02 | 195·24 | 30·59 | 6·38 |
| e05415 | 1490·74 | 22·88 | EpGKAGERGVPGPPGA | P02452 | COL1A1 | Collagen alpha-1(I) chain | 4·98E-03 | 2·18E-02 | 104·57 | 16·19 | 6·46 |
| e06985 | 1667·79 | 40·64 | TGDAGpVGPpGPpGPPGPP | P02452 | COL1A1 | Collagen alpha-1(I) chain | 6·95E-04 | 4·20E-03 | 139·55 | 20·89 | 6·68 |
| e00383 | 893·44 | 23·55 | GPpGPpGARG | P02452 | COL1A1 | Collagen alpha-1(I) chain | 2·98E-05 | 2·99E-04 | 5·97 | 0·88 | 6·77 |
| e09978 | 2006·91 | 32·30 | GpAGPpGEAGKpGEQGVpGDLG | P02452 | COL1A1 | Collagen alpha-1(I) chain | 4·13E-11 | 3·50E-09 | 688·38 | 99·43 | 6·92 |
| e07720 | 1742·85 | 23·74 | LGApGPSGARGERGFpGE | P02452 | COL1A1 | Collagen alpha-1(I) chain | 1·42E-08 | 4·64E-07 | 421·41 | 60·35 | 6·98 |
| e08522 | 1833·88 | 24·05 | GEPGKQGPSGASGERGPpGP | P02452 | COL1A1 | Collagen alpha-1(I) chain | 3·70E-11 | 3·26E-09 | 8608·69 | 1204·19 | 7·15 |
| e14018 | 2612·26 | 23·41 | GPpGPAGEEGKRGARGEPGpTGLpGPpG | P02452 | COL1A1 | Collagen alpha-1(I) chain | 9·36E-03 | 3·54E-02 | 150·07 | 20·08 | 7·48 |
| e12792 | 2398·11 | 22·80 | GNAGPpGPpGPAGKEGGKGPRGETGPA | P02452 | COL1A1 | Collagen alpha-1(I) chain | 2·44E-04 | 1·74E-03 | 268·53 | 32·73 | 8·20 |
| e10219 | 2042·93 | 32·24 | PGAVGPAGKDGEAGAQGPpGpAGP | P02452 | COL1A1 | Collagen alpha-1(I) chain | 8·87E-07 | 1·29E-05 | 75·03 | 8·93 | 8·40 |

| Peptide ID | Mass [Da] | CE-time [Min] | Sequence | UniprotID | Symbol | Protein name | p-value | adj_p-value (BH) | average_cases | average_controls | Fold change |
|---|---|---|---|---|---|---|---|---|---|---|---|
| e00271 | 870·48 | 23·82 | DGVRGLTGP | P02452 | COL1A1 | Collagen alpha-1(I) chain | 1·59E-04 | 1·23E-03 | 210·37 | 24·93 | 8·44 |
| e08666 | 1849·86 | 24·75 | GEPGKQGpSGASGERGPpGP | P02452 | COL1A1 | Collagen alpha-1(I) chain | 3·63E-03 | 1·69E-02 | 724·28 | 80·23 | 9·03 |
| e06027 | 1563·75 | 23·05 | GEpGKAGERGVPGPpGA | P02452 | COL1A1 | Collagen alpha-1(I) chain | 1·37E-06 | 1·88E-05 | 729·54 | 77·38 | 9·43 |
| e02899 | 1221·58 | 26·91 | IGPpGPAGApGDKG | P02452 | COL1A1 | Collagen alpha-1(I) chain | 6·14E-04 | 3·80E-03 | 198·06 | 21·00 | 9·43 |
| e03982 | 1331·62 | 27·87 | RpGEVGPpGPpGPA | P02452 | COL1A1 | Collagen alpha-1(I) chain | 6·35E-05 | 5·77E-04 | 203·79 | 18·36 | 11·10 |
| e15680 | 2907·38 | 20·45 | RGPpGPPGKNGDDGEAGKPGRPGERGPpGp | P02452 | COL1A1 | Collagen alpha-1(I) chain | 5·60E-04 | 3·51E-03 | 1128·87 | 99·15 | 11·39 |
| e14750 | 2745·33 | 21·49 | ERGSPGPAGPKGSpGEAGRpGEAGLpGAKG | P02452 | COL1A1 | Collagen alpha-1(I) chain | 6·91E-06 | 7·99E-05 | 305·19 | 26·05 | 11·71 |
| e12709 | 2384·08 | 22·89 | GApGAEGSpGRDGSpGAKGDRGETGP | P02452 | COL1A1 | Collagen alpha-1(I) chain | 2·87E-08 | 8·07E-07 | 183·03 | 15·15 | 12·08 |
| e09896 | 1993·98 | 21·54 | GPKGSpGEAGRpGEAGLpGAKG | P02452 | COL1A1 | Collagen alpha-1(I) chain | 7·63E-04 | 4·53E-03 | 331·60 | 23·58 | 14·06 |
| e03905 | 1322·60 | 27·62 | GSpGPDGKTGPpGPA | P02452 | COL1A1 | Collagen alpha-1(I) chain | 1·01E-03 | 5·71E-03 | 1559·31 | 73·74 | 21·15 |
| e11048 | 2165·00 | 21·93 | ERGSpGpAGpKGSPGEAGRPGEA | P02452 | COL1A1 | Collagen alpha-1(I) chain | 4·73E-07 | 7·72E-06 | 1092·69 | 48·14 | 22·70 |
| e09075 | 1893·91 | 21·22 | NGDDGEAGKPGRpGERGPp | P02452 | COL1A1 | Collagen alpha-1(I) chain | 1·06E-04 | 8·91E-04 | 466·46 | 16·11 | 28·95 |
| e16540 | 3091·46 | 28·24 | GPAGAAGARGNDGQPGPAGPPGPVGpAGGpGFpGAPG | P02458 | COL2A1 | Collagen alpha-1(II) chain | 1·36E-11 | 1·48E-09 | 50·44 | 212·99 | 0·24 |
| e18281 | 3530·65 | 26·13 | ppGSNGNpGPPGPPGPSGKDGPKGARGDSGPPGRAGEPG | P02458 | COL2A1 | Collagen alpha-1(II) chain | 2·24E-06 | 2·93E-05 | 85·71 | 255·65 | 0·34 |
| e11713 | 2256·97 | 33·66 | GETGAAGpPGPAGPAGERGEQGAPGP | P02458 | COL2A1 | Collagen alpha-1(II) chain | 7·43E-05 | 6·62E-04 | 219·08 | 440·47 | 0·50 |
| e06794 | 1645·76 | 20·76 | DGPKGASGPAGPpGAQGpP | P02458 | COL2A1 | Collagen alpha-1(II) chain | 1·27E-05 | 1·37E-04 | 25·51 | 48·09 | 0·53 |
| e07685 | 1738·76 | 24·95 | GPAGApGpQGFQGNpGEPG | P02458 | COL2A1 | Collagen alpha-1(II) chain | 1·87E-07 | 3·72E-06 | 196·39 | 367·00 | 0·54 |
| e09547 | 1950·84 | 32·03 | GPpGEGGKpGDQGVpGEAGApG | P02458 | COL2A1 | Collagen alpha-1(II) chain | 7·65E-03 | 3·01E-02 | 14·44 | 26·70 | 0·54 |
| e05675 | 1523·62 | 23·69 | GSPGPAGASGNpGTDGIP | P02458 | COL2A1 | Collagen alpha-1(II) chain | 4·04E-08 | 1·08E-06 | 90·65 | 167·03 | 0·54 |
| e04199 | 1353·66 | 25·88 | PVGpSGKDGANGIpG | P02458 | COL2A1 | Collagen alpha-1(II) chain | 2·22E-06 | 2·93E-05 | 69·61 | 125·51 | 0·55 |
| e12658 | 2376·95 | 33·92 | AGppGEKGEPGDDGpSGAEGPpGPQG | P02458 | COL2A1 | Collagen alpha-1(II) chain | 2·82E-04 | 1·95E-03 | 114·24 | 195·12 | 0·59 |

| Peptide ID | Mass [Da] | CE-time [Min] | Sequence | UniprotID | Symbol | Protein name | p-value | adj_p-value (BH) | average_cases | average_controls | Fold change |
|---|---|---|---|---|---|---|---|---|---|---|---|
| e16656 | 3121·43 | 30·16 | FAGpPGADGQpGAKGEQGEAGQKGDAGApGpQGP | P02458 | COL2A1 | Collagen alpha-1(II) chain | 1·37E-08 | 4·55E-07 | 378·87 | 622·21 | 0·61 |
| e18044 | 3458·58 | 31·40 | AGGPGFPGApGAKGEAGpTGARGpEGAQGPRGEPGTPGS | P02458 | COL2A1 | Collagen alpha-1(II) chain | 8·74E-08 | 2·04E-06 | 9323·34 | 13797·94 | 0·68 |
| e04531 | 1392·63 | 21·80 | PGTpGSPGPAGASGNPG | P02458 | COL2A1 | Collagen alpha-1(II) chain | 4·94E-05 | 4·62E-04 | 2289·92 | 3288·88 | 0·70 |
| e17184 | 3242·48 | 22·89 | GPAGFAGPPGADGQPGAKGEQGEAGQKGDAGAPGPQG | P02458 | COL2A1 | Collagen alpha-1(II) chain | 5·57E-06 | 6·51E-05 | 141·05 | 200·73 | 0·70 |
| e12714 | 2385·05 | 33·96 | pAGPPGEKGEPGDDGPSGAEGPpGPQ | P02458 | COL2A1 | Collagen alpha-1(II) chain | 7·27E-04 | 4·37E-03 | 306·99 | 433·05 | 0·71 |
| e01661 | 1084·43 | 25·18 | DGpSGAEGpPGp | P02458 | COL2A1 | Collagen alpha-1(II) chain | 1·22E-06 | 1·72E-05 | 1257·52 | 1760·05 | 0·71 |
| e12007 | 2296·95 | 27·05 | pGADGQpGAKGEQGEAGQKGDAGAp | P02458 | COL2A1 | Collagen alpha-1(II) chain | 2·11E-04 | 1·53E-03 | 177·15 | 241·79 | 0·73 |
| e17377 | 3286·53 | 25·40 | GARGAPGERGETGPPGpAGFAGpPGADGQPGAKGEQ | P02458 | COL2A1 | Collagen alpha-1(II) chain | 4·29E-03 | 1·94E-02 | 95·27 | 129·79 | 0·73 |
| e18922 | 3740·67 | 32·25 | FAGppGADGQPGAKGEQGEAGQKGDAGAPGPQGPSGAPGPQG | P02458 | COL2A1 | Collagen alpha-1(II) chain | 1·62E-03 | 8·54E-03 | 88·15 | 118·59 | 0·74 |
| e19780 | 4113·85 | 24·55 | GAPGERGETGPpGpAGFAGpPGADGQPGAKGEQGEAGQKG-DAGAPG | P02458 | COL2A1 | Collagen alpha-1(II) chain | 1·34E-07 | 2·89E-06 | 270·06 | 360·11 | 0·75 |
| e13634 | 2551·15 | 34·75 | PGVKGESGSpGENGSpGpMGPRGLPGE | P02458 | COL2A1 | Collagen alpha-1(II) chain | 6·19E-04 | 3·81E-03 | 84·73 | 108·73 | 0·78 |
| e13033 | 2442·07 | 34·10 | GpAGPPGEKGEPGDDGPSGAEGPpGPQ | P02458 | COL2A1 | Collagen alpha-1(II) chain | 4·93E-03 | 2·17E-02 | 779·94 | 964·10 | 0·81 |
| e13127 | 2458·08 | 34·20 | GpAGpPGEKGEPGDDGPSGAEGpPGPQ | P02458 | COL2A1 | Collagen alpha-1(II) chain | 1·33E-02 | 4·67E-02 | 32·12 | 36·27 | 0·89 |
| e04196 | 1353·59 | 21·57 | PSGDQGASGpAGPSGP | P02458 | COL2A1 | Collagen alpha-1(II) chain | 7·38E-03 | 2·95E-02 | 228·35 | 162·65 | 1·40 |
| e20089 | 4306·00 | 25·07 | SPGADGPPGRDGAAGVKGDRGETGAVGAPGAPGPpGSpGpAG-PTGKQGD | P02458 | COL2A1 | Collagen alpha-1(II) chain | 1·70E-04 | 1·28E-03 | 1846·64 | 1168·97 | 1·58 |
| e09072 | 1893·84 | 32·04 | pPGEGGKpGDQGVpGEAGApG | P02458 | COL2A1 | Collagen alpha-1(II) chain | 3·59E-04 | 2·42E-03 | 92·38 | 50·25 | 1·84 |
| e19022 | 3774·72 | 22·71 | ERGETGPPGPAGFAGPpGADGQpGAKGEQGEAGQKGDAGAp | P02458 | COL2A1 | Collagen alpha-1(II) chain | 9·84E-12 | 1·19E-09 | 5876·69 | 2260·94 | 2·60 |
| e17318 | 3274·47 | 22·99 | GpAGFAGPPGADGQPGAKGEQGEAGQKGDAGAPGPQG | P02458 | COL2A1 | Collagen alpha-1(II) chain | 8·86E-03 | 3·41E-02 | 218·28 | 47·57 | 4·59 |
| e15593 | 2889·36 | 24·09 | GpSGpAGARGIQGpQGPRGDKGEAGEPGER | P02458 | COL2A1 | Collagen alpha-1(II) chain | 1·45E-02 | 4·98E-02 | 385·56 | 65·55 | 5·88 |

| Pep-tide ID | Mass [Da] | CE-time [Min] | Sequence | UniprotID | Symbol | Protein name | p-value | adj_p-value (BH) | aver-age_cases | aver-age_con-trols | Fold change |
|---|---|---|---|---|---|---|---|---|---|---|---|
| e05303 | 1475·78 | 22·81 | GRpGPpGPQGARGQp | P02458 | COL2A1 | Collagen alpha-1(II) chain | 1·58E-08 | 4·97E-07 | 583·49 | 54·84 | 10·64 |
| e06181 | 1579·65 | 39·92 | GETGAVGApGApGPpGSPG | P02458 | COL2A1 | Collagen alpha-1(II) chain | 2·40E-04 | 1·72E-03 | 42·80 | 1·92 | 22·28 |
| e06611 | 1626·72 | 30·52 | EGGKpGDQGVpGEAGApG | P02458 | COL2A1 | Collagen alpha-1(II) chain | 7·53E-03 | 2·97E-02 | 1414·06 | 32·10 | 44·05 |
| e12451 | 2347·04 | 21·91 | SPGVSGpKGDAGQpGEKGSPGAQGPP | P02461 | COL3A1 | Collagen alpha-1(III) chain | 1·38E-02 | 4·78E-02 | 1·87 | 9·81 | 0·19 |
| e20018 | 4255·98 | 25·73 | FPGSPGAKGEVGPAGSPGSNGAPGQRGE-PGPQGHAGAQGPpGppGIN | P02461 | COL3A1 | Collagen alpha-1(III) chain | 8·07E-11 | 6·00E-09 | 104·21 | 522·08 | 0·20 |
| e01767 | 1097·50 | 21·08 | PGpTGPGGDkGD | P02461 | COL3A1 | Collagen alpha-1(III) chain | 3·63E-03 | 1·69E-02 | 5·13 | 22·42 | 0·23 |
| e17112 | 3226·48 | 22·73 | PSGSPGKDGPpGpAGNTGAPGSPGVSGPKGDAGQPGE | P02461 | COL3A1 | Collagen alpha-1(III) chain | 7·75E-03 | 3·04E-02 | 3·48 | 13·56 | 0·26 |
| e16231 | 3025·40 | 29·89 | pPGKNGETGPQGppGPTGPGGDKGDTGPPGpQG | P02461 | COL3A1 | Collagen alpha-1(III) chain | 9·27E-05 | 7·89E-04 | 11·09 | 41·81 | 0·27 |
| e18864 | 3718·72 | 32·42 | GpAGPPGPPGPPGTSGHPGSPGSPGYQGPPGEPGQAGPSGPPG | P02461 | COL3A1 | Collagen alpha-1(III) chain | 5·59E-13 | 9·14E-11 | 116·62 | 383·80 | 0·30 |
| e01648 | 1082·50 | 23·81 | GpEGGKGAAGPpG | P02461 | COL3A1 | Collagen alpha-1(III) chain | 1·23E-04 | 1·01E-03 | 45·43 | 129·02 | 0·35 |
| e14821 | 2758·25 | 28·98 | KNGETGPQGppGPTGPGGDKGDTGPpGPQG | P02461 | COL3A1 | Collagen alpha-1(III) chain | 7·38E-08 | 1·74E-06 | 32·55 | 86·06 | 0·38 |
| e15129 | 2809·20 | 24·38 | ERGEAGIpGVpGAKGEDGKDGSPGEpGANG | P02461 | COL3A1 | Collagen alpha-1(III) chain | 1·09E-11 | 1·25E-09 | 164·64 | 390·81 | 0·42 |
| e01935 | 1115·51 | 21·60 | DGESGRpGRpG | P02461 | COL3A1 | Collagen alpha-1(III) chain | 2·51E-07 | 4·68E-06 | 78·69 | 167·08 | 0·47 |
| e17270 | 3262·50 | 22·36 | APGAPGHPGPPGPVGPAGKSGDRGESGPAGPAGAPGPAG | P02461 | COL3A1 | Collagen alpha-1(III) chain | 2·56E-03 | 1·25E-02 | 23·24 | 49·12 | 0·47 |
| e08188 | 1794·80 | 24·01 | GNDGApGKNGERGGpGGpGP | P02461 | COL3A1 | Collagen alpha-1(III) chain | 1·70E-11 | 1·69E-09 | 546·54 | 1127·58 | 0·48 |
| e06841 | 1649·73 | 22·75 | GPpGpAGQpGDKGEGGAPG | P02461 | COL3A1 | Collagen alpha-1(III) chain | 8·90E-11 | 6·18E-09 | 387·63 | 792·57 | 0·49 |
| e08165 | 1790·81 | 23·85 | GESGRPGppGpSGPRGQPG | P02461 | COL3A1 | Collagen alpha-1(III) chain | 3·05E-07 | 5·33E-06 | 71·15 | 139·82 | 0·51 |
| e03938 | 1326·54 | 29·12 | SpGGpGSDGKpGPpG | P02461 | COL3A1 | Collagen alpha-1(III) chain | 4·70E-03 | 2·09E-02 | 28·79 | 55·35 | 0·52 |
| e17170 | 3239·49 | 30·85 | NTGApGSPGVSGpKGDAGQpGEKGSPGAQGPPGAPGP | P02461 | COL3A1 | Collagen alpha-1(III) chain | 9·97E-03 | 3·73E-02 | 26·49 | 49·67 | 0·53 |
| e17866 | 3409·61 | 31·29 | NTGApGSpGVSGPKGDAGQPGEKGSpGAQGPPGAPGPLG | P02461 | COL3A1 | Collagen alpha-1(III) chain | 1·06E-06 | 1·53E-05 | 164·18 | 296·60 | 0·55 |

| Peptide ID | Mass [Da] | CE-time [Min] | Sequence | UniprotID | Symbol | Protein name | p-value | adj_p-value (BH) | average_cases | average_controls | Fold change |
|---|---|---|---|---|---|---|---|---|---|---|---|
| e15411 | 2854·37 | 34·67 | PQGPpGPTGpGGDKGDTGPpGPQGLQGLpGT | P02461 | COL3A1 | Collagen alpha-1(III) chain | 1·27E-08 | 4·35E-07 | 1170·70 | 2075·98 | 0·56 |
| e16388 | 3058·39 | 24·82 | GSPGKDGppGpAGNTGAPGSPGVSGPKGDAGQPGE | P02461 | COL3A1 | Collagen alpha-1(III) chain | 2·88E-06 | 3·68E-05 | 103·64 | 180·77 | 0·57 |
| e17238 | 3255·49 | 30·76 | NTGApGSpGVSGPKGDAGQpGEKGSpGAQGPPGAPGP | P02461 | COL3A1 | Collagen alpha-1(III) chain | 2·02E-07 | 3·92E-06 | 162·21 | 282·86 | 0·57 |
| e14315 | 2663·21 | 23·57 | NRGERGSEGSPGHpGQpGPPGpPGApGP | P02461 | COL3A1 | Collagen alpha-1(III) chain | 1·54E-05 | 1·65E-04 | 189·46 | 313·82 | 0·60 |
| e14224 | 2647·21 | 23·52 | NRGERGSEGSPGHpGQPGPpGPPGApGP | P02461 | COL3A1 | Collagen alpha-1(III) chain | 7·41E-03 | 2·95E-02 | 35·27 | 57·68 | 0·61 |
| e17254 | 3258·47 | 22·83 | ENGKPGEpGpKGDAGApGApGGKGDAGApGERGpPG | P02461 | COL3A1 | Collagen alpha-1(III) chain | 2·88E-09 | 1·20E-07 | 1067·62 | 1745·05 | 0·61 |
| e14644 | 2726·26 | 28·92 | KNGETGPQGPPGPTGPGGDKGDTGPpGPQG | P02461 | COL3A1 | Collagen alpha-1(III) chain | 2·53E-05 | 2·59E-04 | 110·01 | 176·11 | 0·62 |
| e13662 | 2557·17 | 28·23 | KNGETGPQGPPGpTGPGGDKGDTGPpGP | P02461 | COL3A1 | Collagen alpha-1(III) chain | 1·50E-07 | 3·16E-06 | 83·64 | 133·64 | 0·63 |
| e12496 | 2353·99 | 27·28 | AGIpGVpGAKGEDGKDGSpGEpGANG | P02461 | COL3A1 | Collagen alpha-1(III) chain | 8·33E-06 | 9·49E-05 | 86·64 | 136·96 | 0·63 |
| e18041 | 3457·61 | 31·46 | NTGAPGSpGVSGpKGDAGQpGEKGSpGAQGppGAPGPLG | P02461 | COL3A1 | Collagen alpha-1(III) chain | 1·79E-08 | 5·40E-07 | 11914·80 | 18291·13 | 0·65 |
| e09629 | 1962·88 | 31·97 | QGLpGTGGPpGENGKpGEpGP | P02461 | COL3A1 | Collagen alpha-1(III) chain | 6·30E-03 | 2·62E-02 | 63·16 | 96·33 | 0·66 |
| e09856 | 1989·88 | 32·53 | SNGNPGppGPSGSpGKDGPPGp | P02461 | COL3A1 | Collagen alpha-1(III) chain | 1·17E-04 | 9·75E-04 | 154·97 | 234·43 | 0·66 |
| e13697 | 2563·15 | 21·25 | ApGPAGSRGApGPQGpRGDKGETGERG | P02461 | COL3A1 | Collagen alpha-1(III) chain | 3·55E-07 | 6·03E-06 | 678·20 | 1019·52 | 0·67 |
| e18092 | 3473·60 | 31·47 | NTGApGSpGVSGpKGDAGQpGEKGSpGAQGpPGAPGpLG | P02461 | COL3A1 | Collagen alpha-1(III) chain | 1·69E-07 | 3·40E-06 | 2587·84 | 3888·82 | 0·67 |
| e16132 | 3001·44 | 34·97 | YDVKSGVAVGGLAGYpGpAGPPGPPGPPGTSGH | P02461 | COL3A1 | Collagen alpha-1(III) chain | 4·86E-05 | 4·58E-04 | 5512·16 | 8257·45 | 0·67 |
| e16085 | 2991·41 | 20·97 | SPGGPGAAGFPGARGLPGPPGSNGNPGPPGPSGSp | P02461 | COL3A1 | Collagen alpha-1(III) chain | 8·64E-03 | 3·34E-02 | 118·96 | 176·06 | 0·68 |
| e17922 | 3425·61 | 31·29 | NTGApGSPGVSGPKGDAGQpGEKGSpGAQGpPGAPGPLG | P02461 | COL3A1 | Collagen alpha-1(III) chain | 5·47E-07 | 8·76E-06 | 1594·62 | 2348·01 | 0·68 |
| e11780 | 2266·03 | 22·25 | QNGEpGGKGERGApGEKGEGGPpG | P02461 | COL3A1 | Collagen alpha-1(III) chain | 7·66E-07 | 1·14E-05 | 2716·01 | 3928·14 | 0·69 |
| e02160 | 1140·48 | 21·09 | DKGDTGPPGPQG | P02461 | COL3A1 | Collagen alpha-1(III) chain | 4·94E-05 | 4·62E-04 | 1301·13 | 1818·67 | 0·72 |

| Peptide ID | Mass [Da] | CE-time [Min] | Sequence | UniprotID | Symbol | Protein name | p-value | adj_p-value (BH) | average_cases | average_controls | Fold change |
|---|---|---|---|---|---|---|---|---|---|---|---|
| e17311 | 3271·52 | 30·76 | NTGApGSpGVSGpKGDAGQpGEKGSPGAQGPPGAPGp | P02461 | COL3A1 | Collagen alpha-1(III) chain | 6·90E-07 | 1·07E-05 | 756·09 | 1043·56 | 0·72 |
| e02041 | 1127·52 | 20·81 | ApGKNGERGGpG | P02461 | COL3A1 | Collagen alpha-1(III) chain | 1·36E-02 | 4·73E-02 | 16·63 | 22·79 | 0·73 |
| e16220 | 3023·41 | 20·91 | SPGGPGAAGFPGARGLPGPPGSNGNpGppGPSGSP | P02461 | COL3A1 | Collagen alpha-1(III) chain | 3·55E-05 | 3·48E-04 | 252·46 | 341·90 | 0·74 |
| e15237 | 2825·28 | 24·45 | ERGEAGIpGVpGAKGEDGKDGSpGEpGANG | P02461 | COL3A1 | Collagen alpha-1(III) chain | 4·84E-05 | 4·58E-04 | 9479·35 | 12183·74 | 0·78 |
| e10881 | 2137·93 | 21·77 | NGEpGGKGERGAPGEKGEGGppG | P02461 | COL3A1 | Collagen alpha-1(III) chain | 1·00E-03 | 5·68E-03 | 410·45 | 524·63 | 0·78 |
| e10257 | 2046·92 | 32·75 | GSNGNpGpPGPSGSpGKDGPpGP | P02461 | COL3A1 | Collagen alpha-1(III) chain | 3·89E-04 | 2·59E-03 | 1245·56 | 1558·11 | 0·80 |
| e07676 | 1737·78 | 23·79 | NDGAPGKNGERGGpGGpGp | P02461 | COL3A1 | Collagen alpha-1(III) chain | 2·55E-04 | 1·80E-03 | 2568·59 | 3209·83 | 0·80 |
| e09891 | 1993·88 | 32·19 | SEGSPGHpGQPGpPGpPGApGP | P02461 | COL3A1 | Collagen alpha-1(III) chain | 1·34E-02 | 4·68E-02 | 185·71 | 230·51 | 0·81 |
| e15323 | 2839·36 | 24·20 | GPAGPRGPVGPSGPPGKDGTSGHPGPIGppGP | P02461 | COL3A1 | Collagen alpha-1(III) chain | 2·52E-06 | 3·28E-05 | 574·73 | 702·42 | 0·82 |
| e14735 | 2742·25 | 28·93 | KNGETGPQGPPGpTGPGGDKGDTGPpGPQG | P02461 | COL3A1 | Collagen alpha-1(III) chain | 9·62E-03 | 3·62E-02 | 1386·97 | 1693·26 | 0·82 |
| e20087 | 4305·93 | 28·79 | ARGNDGARGSDGQpGpPGPpGTAGFpGSpGAKGEVGpAGSpGSN-GApG | P02461 | COL3A1 | Collagen alpha-1(III) chain | 5·22E-03 | 2·27E-02 | 2448·30 | 2973·08 | 0·82 |
| e04649 | 1405·70 | 23·33 | DGVPGKDGPRGPTGP | P02461 | COL3A1 | Collagen alpha-1(III) chain | 7·04E-03 | 2·83E-02 | 90·96 | 109·56 | 0·83 |
| e17856 | 3405·56 | 25·85 | ARGNDGARGSDGQPGPpGppGTAGFpGSpGAKGEVGP | P02461 | COL3A1 | Collagen alpha-1(III) chain | 1·77E-04 | 1·33E-03 | 1924·16 | 2316·69 | 0·83 |
| e06220 | 1583·70 | 23·30 | NDGApGKNGERGGpGGp | P02461 | COL3A1 | Collagen alpha-1(III) chain | 1·42E-07 | 3·02E-06 | 170·44 | 194·53 | 0·88 |
| e14253 | 2654·20 | 23·89 | ERGEAGIpGVpGAKGEDGKDGSpGEpGA | P02461 | COL3A1 | Collagen alpha-1(III) chain | 2·29E-05 | 2·38E-04 | 189·63 | 201·39 | 0·94 |
| e06598 | 1624·73 | 24·42 | NDGApGKNGERGGpGGPG | P02461 | COL3A1 | Collagen alpha-1(III) chain | 1·44E-02 | 4·96E-02 | 226·93 | 218·43 | 1·04 |
| e10386 | 2062·93 | 26·46 | DAGAPGApGGKGDAGApGERGpPG | P02461 | COL3A1 | Collagen alpha-1(III) chain | 9·28E-04 | 5·30E-03 | 373·28 | 358·57 | 1·04 |
| e03194 | 1251·62 | 22·45 | DGVPGKDGPRGPT | P02461 | COL3A1 | Collagen alpha-1(III) chain | 4·24E-03 | 1·92E-02 | 272·28 | 249·61 | 1·09 |
| e14956 | 2779·23 | 28·94 | HRGFpGNPGAPGSPGPAGQqGAIGSpGPAGP | P02461 | COL3A1 | Collagen alpha-1(III) chain | 6·32E-03 | 2·62E-02 | 71·08 | 59·60 | 1·19 |
| e01923 | 1114·49 | 25·71 | SpGERGETGPp | P02461 | COL3A1 | Collagen alpha-1(III) chain | 3·24E-03 | 1·53E-02 | 3320·91 | 2588·47 | 1·28 |

| Peptide ID | Mass [Da] | CE-time [Min] | Sequence | UniprotID | Symbol | Protein name | p-value | adj_p-value (BH) | average_cases | average_controls | Fold change |
|---|---|---|---|---|---|---|---|---|---|---|---|
| e20132 | 4338·00 | 25·14 | LQGLPGTGGppGENGKpGEpGpKGDAGApGApGGKG-DAGApGERGpPG | P02461 | COL3A1 | Collagen alpha-1(III) chain | 2·18E-04 | 1·57E-03 | 991·05 | 728·76 | 1·36 |
| e13816 | 2583·17 | 23·63 | ERGEAGIpGVpGAKGEDGKDGSpGEpG | P02461 | COL3A1 | Collagen alpha-1(III) chain | 3·10E-05 | 3·09E-04 | 275·19 | 184·79 | 1·49 |
| e19159 | 3822·88 | 24·17 | QQGAIGSPGPAGPRGPVGpSGpPGKDGTSGHpGPIGPPGpRG | P02461 | COL3A1 | Collagen alpha-1(III) chain | 1·52E-04 | 1·19E-03 | 62·84 | 41·23 | 1·52 |
| e20109 | 4321·99 | 25·14 | LQGLpGTGGppGENGKpGEPGpKGDAGAPGAPGGKG-DAGApGERGppG | P02461 | COL3A1 | Collagen alpha-1(III) chain | 6·56E-08 | 1·62E-06 | 10890·25 | 6544·08 | 1·66 |
| e05577 | 1510·67 | 23·44 | GGAGEpGKNGAKGEpGP | P02461 | COL3A1 | Collagen alpha-1(III) chain | 1·89E-03 | 9·77E-03 | 248·28 | 137·29 | 1·81 |
| e19801 | 4121·91 | 23·26 | GEPGRDGVPGGPGMRGMPGSPGGPGSDGKPGPpGSQGESGRpGpP | P02461 | COL3A1 | Collagen alpha-1(III) chain | 5·41E-03 | 2·33E-02 | 2821·21 | 1312·82 | 2·15 |
| e17824 | 3400·55 | 30·92 | GEVGPAGSPGSNGApGQRGEpGpQGHAGAQGPPGPPGI | P02461 | COL3A1 | Collagen alpha-1(III) chain | 3·67E-04 | 2·48E-03 | 515·32 | 232·61 | 2·22 |
| e16173 | 3012·38 | 22·19 | LGSPGpKGDKGEpGGpGADGVPGKDGPRGPTGP | P02461 | COL3A1 | Collagen alpha-1(III) chain | 9·46E-04 | 5·39E-03 | 681·50 | 306·11 | 2·23 |
| e12891 | 2412·11 | 27·18 | RGGAGPPGpEGGKGAAGPpGpPGAAGTpG | P02461 | COL3A1 | Collagen alpha-1(III) chain | 4·48E-03 | 2·01E-02 | 653·57 | 293·39 | 2·23 |
| e05814 | 1538·78 | 20·42 | GEpGKNGAKGEPGpRG | P02461 | COL3A1 | Collagen alpha-1(III) chain | 2·82E-03 | 1·37E-02 | 133·97 | 59·19 | 2·26 |
| e03633 | 1296·61 | 21·80 | RGApGEKGEGGpPG | P02461 | COL3A1 | Collagen alpha-1(III) chain | 3·10E-03 | 1·48E-02 | 56·43 | 23·70 | 2·38 |
| e19548 | 4008·81 | 23·32 | GEPGRDGVPGGPGMRGMPGSPGGPGSDGKPGPPGSQGESGRpGp | P02461 | COL3A1 | Collagen alpha-1(III) chain | 1·47E-08 | 4·66E-07 | 10437·78 | 4272·68 | 2·44 |
| e11599 | 2241·01 | 33·65 | PpGSNGNpGPpGPSGSPGKDGPpGP | P02461 | COL3A1 | Collagen alpha-1(III) chain | 5·53E-03 | 2·36E-02 | 102·91 | 41·18 | 2·50 |
| e19321 | 3890·78 | 24·34 | SNGNPGPPGPSGSPGKDGPPGpAGNTGApGSpGVSGPKGDAGQPG | P02461 | COL3A1 | Collagen alpha-1(III) chain | 2·83E-15 | 1·62E-12 | 2909·11 | 941·85 | 3·09 |
| e05918 | 1551·69 | 29·71 | GTGGPpGENGKpGEpGP | P02461 | COL3A1 | Collagen alpha-1(III) chain | 1·16E-04 | 9·68E-04 | 640·59 | 199·55 | 3·21 |
| e15095 | 2802·31 | 19·60 | AGERGAPGFRGPAGPNGIPGEKGpAGERGA | P02461 | COL3A1 | Collagen alpha-1(III) chain | 3·93E-03 | 1·81E-02 | 31·45 | 8·79 | 3·58 |
| e04723 | 1412·62 | 29·58 | SpGAPGApGHpGPpGP | P02461 | COL3A1 | Collagen alpha-1(III) chain | 9·82E-04 | 5·58E-03 | 44·24 | 12·05 | 3·67 |
| e09966 | 2004·93 | 25·02 | GARGNDGARGSDGQPGPpGPpG | P02461 | COL3A1 | Collagen alpha-1(III) chain | 5·69E-10 | 3·72E-08 | 799·24 | 216·68 | 3·69 |
| e10019 | 2012·95 | 25·18 | RGGAGPpGPEGGKGAAGPpGPpGA | P02461 | COL3A1 | Collagen alpha-1(III) chain | 3·92E-06 | 4·68E-05 | 99·57 | 26·27 | 3·79 |
| e00184 | 852·42 | 23·26 | DGPRGPTGP | P02461 | COL3A1 | Collagen alpha-1(III) chain | 2·09E-03 | 1·06E-02 | 22·18 | 5·70 | 3·89 |

| Peptide ID | Mass [Da] | CE-time [Min] | Sequence | UniprotID | Symbol | Protein name | p-value | adj_p-value (BH) | average_cases | average_controls | Fold change |
|---|---|---|---|---|---|---|---|---|---|---|---|
| e10046 | 2016·02 | 21·43 | GIpGEKGPAGERGApGPAGPRG | P02461 | COL3A1 | Collagen alpha-1(III) chain | 7·62E-04 | 4·53E-03 | 4267·02 | 1018·91 | 4·19 |
| e14434 | 2684·27 | 21·39 | GEpGRDGNpGSDGLpGRDGSpGGKGDRG | P02461 | COL3A1 | Collagen alpha-1(III) chain | 9·30E-03 | 3·53E-02 | 118·28 | 26·91 | 4·40 |
| e03927 | 1325·56 | 21·47 | GEpGKNGAKGEpGP | P02461 | COL3A1 | Collagen alpha-1(III) chain | 8·51E-04 | 4·95E-03 | 201·41 | 43·33 | 4·65 |
| e05434 | 1492·71 | 22·74 | GESGRpGPpGPSGPRG | P02461 | COL3A1 | Collagen alpha-1(III) chain | 6·08E-04 | 3·77E-03 | 76·05 | 16·30 | 4·66 |
| e02000 | 1123·50 | 26·03 | GGAGPpGPEGGKGA | P02461 | COL3A1 | Collagen alpha-1(III) chain | 3·92E-04 | 2·60E-03 | 106·03 | 20·36 | 5·21 |
| e08819 | 1867·92 | 21·32 | ApGPQGpRGDKGETGERGA | P02461 | COL3A1 | Collagen alpha-1(III) chain | 1·12E-05 | 1·23E-04 | 297·05 | 56·18 | 5·29 |
| e00837 | 968·45 | 26·63 | KGDTGPpGPQ | P02461 | COL3A1 | Collagen alpha-1(III) chain | 8·13E-03 | 3·17E-02 | 49·63 | 9·34 | 5·31 |
| e06617 | 1627·70 | 29·59 | MpGSpGGpGSDGKpGpPG | P02461 | COL3A1 | Collagen alpha-1(III) chain | 8·38E-05 | 7·25E-04 | 6322·27 | 1040·89 | 6·07 |
| e13241 | 2481·12 | 27·35 | NTGApGSpGVSGpKGDAGQPGEKGSPGA | P02461 | COL3A1 | Collagen alpha-1(III) chain | 7·03E-08 | 1·68E-06 | 298·26 | 47·71 | 6·25 |
| e01416 | 1052·48 | 25·16 | GGAGPpGPEGGKG | P02461 | COL3A1 | Collagen alpha-1(III) chain | 7·46E-04 | 4·47E-03 | 258·37 | 40·21 | 6·43 |
| e04135 | 1347·68 | 21·86 | GRPGppGPSGPRGQ | P02461 | COL3A1 | Collagen alpha-1(III) chain | 3·24E-06 | 3·99E-05 | 106·39 | 16·02 | 6·64 |
| e13362 | 2507·13 | 22·82 | ApGQNGEPGGkGERGAPGEkGEGGPpG | P02461 | COL3A1 | Collagen alpha-1(III) chain | 6·62E-04 | 4·05E-03 | 639·13 | 94·24 | 6·78 |
| e14880 | 2768·26 | 24·19 | ERGEAGIpGVpGAKGEDGKDGSpGEpGAN | P02461 | COL3A1 | Collagen alpha-1(III) chain | 2·06E-03 | 1·05E-02 | 142·63 | 19·30 | 7·39 |
| e12612 | 2369·07 | 34·14 | PpGSNGNpGpPGPSGSpGKDGPPGPAG | P02461 | COL3A1 | Collagen alpha-1(III) chain | 7·45E-10 | 4·37E-08 | 187·57 | 24·75 | 7·58 |
| e03325 | 1265·61 | 21·80 | ApGKNGERGGPGGP | P02461 | COL3A1 | Collagen alpha-1(III) chain | 8·39E-11 | 6·00E-09 | 390·08 | 51·07 | 7·64 |
| e02344 | 1160·54 | 26·41 | SpGPAGPRGPVGP | P02461 | COL3A1 | Collagen alpha-1(III) chain | 1·28E-02 | 4·56E-02 | 166·72 | 21·79 | 7·65 |
| e14469 | 2691·26 | 24·02 | IGSpGPAGPRGPVGpSGPPGKDGTSGHpGP | P02461 | COL3A1 | Collagen alpha-1(III) chain | 3·28E-08 | 8·95E-07 | 79·72 | 10·00 | 7·97 |
| e13352 | 2505·20 | 20·93 | GEpGPRGERGEAGIPGVpGAKGEDGK | P02461 | COL3A1 | Collagen alpha-1(III) chain | 6·84E-03 | 2·79E-02 | 169·67 | 20·47 | 8·29 |
| e14566 | 2710·30 | 28·62 | RGGAGpPGPEGGKGAAGPpGPpGAAGTpGLQG | P02461 | COL3A1 | Collagen alpha-1(III) chain | 1·71E-08 | 5·22E-07 | 838·96 | 97·60 | 8·60 |
| e06066 | 1567·69 | 29·54 | NGSpGAPGApGHPGPpGP | P02461 | COL3A1 | Collagen alpha-1(III) chain | 8·19E-06 | 9·38E-05 | 398·57 | 39·61 | 10·06 |
| e05726 | 1529·70 | 39·89 | GPpGFTGppGPPGPPGP | P02462 | COL4A1 | Collagen alpha-1(IV) chain | 1·58E-04 | 1·22E-03 | 193·82 | 363·87 | 0·53 |

| Peptide ID | Mass [Da] | CE-time [Min] | Sequence | UniprotID | Symbol | Protein name | p-value | adj_p-value (BH) | average_cases | average_controls | Fold change |
|---|---|---|---|---|---|---|---|---|---|---|---|
| e05864 | 1545·70 | 40·28 | GppGFTGPpGpPGPPGP | P02462 | COL4A1 | Collagen alpha-1(IV) chain | 4·86E-04 | 3·14E-03 | 1315·50 | 1995·33 | 0·66 |
| e17983 | 3441·61 | 31·36 | DGAPGQKGEMGPAGPTGPRGFpGppGPDGLPGSMGPP | P02462 | COL4A1 | Collagen alpha-1(IV) chain | 1·23E-07 | 2·72E-06 | 6488·86 | 9407·16 | 0·69 |
| e12114 | 2309·19 | 19·56 | GEKGEKGSIGIPGMPGSPGLKGSpG | P02462 | COL4A1 | Collagen alpha-1(IV) chain | 4·27E-04 | 2·79E-03 | 18·98 | 9·65 | 1·97 |
| e18981 | 3759·86 | 19·39 | GMpGVGEKGEpGKPGPRGKPGKDGDKGEKGSPGFPGEPG | P02462 | COL4A1 | Collagen alpha-1(IV) chain | 2·84E-03 | 1·37E-02 | 4743·92 | 1851·78 | 2·56 |
| e05900 | 1549·69 | 40·72 | QGpQGEKGEAGppGPP | P02462 | COL4A1 | Collagen alpha-1(IV) chain | 1·05E-02 | 3·91E-02 | 82·58 | 30·49 | 2·71 |
| e16099 | 2995·38 | 29·72 | TpSQTTDERGPPGEQGPPGPPGPPGVPGIDG | P20849 | COL9A1 | Collagen alpha-1(IX) chain | 2·68E-11 | 2·55E-09 | 38·52 | 120·41 | 0·32 |
| e06111 | 1572·67 | 39·90 | pGADGLTGPDGSpGSIGS | P20849 | COL9A1 | Collagen alpha-1(IX) chain | 5·85E-03 | 2·47E-02 | 30·47 | 23·17 | 1·31 |
| e09264 | 1916·85 | 24·65 | LpGAPGDQGQRGPpGEAGPK | P20849 | COL9A1 | Collagen alpha-1(IX) chain | 8·65E-04 | 5·00E-03 | 70·96 | 11·49 | 6·17 |
| e08146 | 1787·93 | 24·63 | LGRVGPVGDpGRRGppGP | P20849 | COL9A1 | Collagen alpha-1(IX) chain | 7·50E-04 | 4·48E-03 | 95·94 | 8·92 | 10·76 |
| e01721 | 1092·46 | 36·37 | GPpGPpGpAGEp | P20849 | COL9A1 | Collagen alpha-1(IX) chain | 7·79E-03 | 3·05E-02 | 258·07 | 18·14 | 14·23 |
| e18967 | 3756·69 | 32·37 | GPPGEPGPAGQDGPPGDKGDDGEPGQTGSPGPTGEPGPSGPP | P20908 | COL5A1 | Collagen alpha-1(V) chain | 2·51E-07 | 4·68E-06 | 84·97 | 168·65 | 0·50 |
| e18060 | 3463·59 | 30·87 | PGEKGETGDVGQMGPPGPPGPRGpSGApGADGpQGPPG | P20908 | COL5A1 | Collagen alpha-1(V) chain | 9·17E-05 | 7·84E-04 | 842·68 | 1278·51 | 0·66 |
| e05011 | 1441·68 | 39·08 | pGPPGEPGPAGQDGPP | P20908 | COL5A1 | Collagen alpha-1(V) chain | 3·12E-03 | 1·48E-02 | 391·40 | 546·14 | 0·72 |
| e19013 | 3772·71 | 32·53 | GpPGEPGPAGQDGPPGDKGDDGEPGQTGSPGPTGEPGPSGPP | P20908 | COL5A1 | Collagen alpha-1(V) chain | 1·94E-04 | 1·43E-03 | 78·16 | 107·00 | 0·73 |
| e18346 | 3549·51 | 31·23 | DpGPPGEPGPAGQDGPPGDKGDDGEPGQTGSPGPTGEPG | P20908 | COL5A1 | Collagen alpha-1(V) chain | 1·71E-03 | 8·94E-03 | 434·00 | 502·91 | 0·86 |
| e06015 | 1562·69 | 22·48 | GApGADGpQGpPGGIGNP | P20908 | COL5A1 | Collagen alpha-1(V) chain | 5·23E-03 | 2·27E-02 | 1733·44 | 1633·26 | 1·06 |
| e12971 | 2428·09 | 27·47 | VVGpQGPTGETGpMGERGHpGPpGP | P20908 | COL5A1 | Collagen alpha-1(V) chain | 1·27E-02 | 4·55E-02 | 38·34 | 29·97 | 1·28 |
| e03053 | 1238·54 | 27·56 | GEAGHPGPpGPPGP | P20908 | COL5A1 | Collagen alpha-1(V) chain | 9·40E-05 | 7·97E-04 | 86·81 | 40·22 | 2·16 |
| e09005 | 1885·84 | 32·28 | PGppGpRGPSGAPGADGpQGP | P20908 | COL5A1 | Collagen alpha-1(V) chain | 1·01E-02 | 3·77E-02 | 62·03 | 27·11 | 2·29 |
| e11963 | 2291·01 | 33·46 | ppGpPGVVGpQGPTGETGPMGERG | P20908 | COL5A1 | Collagen alpha-1(V) chain | 1·01E-07 | 2·33E-06 | 152·46 | 61·16 | 2·49 |

| Peptide ID | Mass [Da] | CE-time [Min] | Sequence | UniprotID | Symbol | Protein name | p-value | adj_p-value (BH) | average_cases | average_controls | Fold change |
|---|---|---|---|---|---|---|---|---|---|---|---|
| e11907 | 2282·05 | 25·63 | GADGPPGHPGKEGPpGEKGGqGppG | P20908 | COL5A1 | Collagen alpha-1(V) chain | 8·78E-04 | 5·05E-03 | 98·77 | 26·23 | 3·76 |
| e16400 | 3060·53 | 21·10 | PQGPTGETGPMGERGHpGpPGPPGEQGLPGLA | P20908 | COL5A1 | Collagen alpha-1(V) chain | 1·46E-08 | 4·66E-07 | 49·35 | 10·73 | 4·60 |
| e12651 | 2375·21 | 22·51 | KGGQGpPGPQGpIGYPGPRGVKGAD | P20908 | COL5A1 | Collagen alpha-1(V) chain | 2·72E-05 | 2·76E-04 | 34095·59 | 6628·88 | 5·14 |
| e17131 | 3231·42 | 22·51 | PPGDPGLMGERGEDGpAGNGTEGFpGFPGYPGN | P12109 | COL6A1 | Collagen alpha-1(VI) chain | 8·67E-09 | 3·20E-07 | 82·16 | 212·17 | 0·39 |
| e17202 | 3247·38 | 22·46 | PpGDpGLMGERGEDGpAGNGTEGFPGFPGYPGN | P12109 | COL6A1 | Collagen alpha-1(VI) chain | 2·73E-09 | 1·17E-07 | 305·87 | 632·88 | 0·48 |
| e17637 | 3347·45 | 30·65 | EAGRpGSSGpSGDEGQPGEPGPPGEKGEAGDEGNPG | P12109 | COL6A1 | Collagen alpha-1(VI) chain | 7·27E-04 | 4·37E-03 | 44·18 | 85·44 | 0·52 |
| e19631 | 4043·79 | 33·01 | GEpGADGEAGRpGSSGpSGDEGQPGEPGPPGEKGEAGDEGNPGP | P12109 | COL6A1 | Collagen alpha-1(VI) chain | 2·65E-06 | 3·41E-05 | 233·81 | 387·36 | 0·60 |
| e17705 | 3363·44 | 30·78 | EAGRpGSSGpSGDEGQpGEPGPPGEKGEAGDEGNPG | P12109 | COL6A1 | Collagen alpha-1(VI) chain | 2·19E-03 | 1·10E-02 | 155·08 | 230·82 | 0·67 |
| e20326 | 4467·97 | 28·94 | GSEGARGAPGPAGPPGDPGLMGERGEDGPAGNG-TEGFpGFpGYpGNR | P12109 | COL6A1 | Collagen alpha-1(VI) chain | 5·02E-04 | 3·23E-03 | 160·14 | 236·31 | 0·68 |
| e19757 | 4100·77 | 33·06 | PGADGEAGRPGSSGpSGDEGQpGEpGPPGEKGEAGDEGNPGPDGA | P12109 | COL6A1 | Collagen alpha-1(VI) chain | 1·03E-04 | 8·67E-04 | 129·91 | 185·16 | 0·70 |
| e16718 | 3136·41 | 30·22 | DPGLMGERGEDGpAGNGTEGFpGFPGYPGNR | P12109 | COL6A1 | Collagen alpha-1(VI) chain | 3·94E-04 | 2·61E-03 | 124·00 | 154·65 | 0·80 |
| e17447 | 3304·39 | 22·57 | GPpGDpGLMGERGEDGpAGNGTEGFPGFPGYPGN | P12109 | COL6A1 | Collagen alpha-1(VI) chain | 1·79E-04 | 1·34E-03 | 491·68 | 593·84 | 0·83 |
| e16281 | 3037·34 | 22·30 | GADGEAGRPGSSGPSGDEGQPGEpGppGEKGEA | P12109 | COL6A1 | Collagen alpha-1(VI) chain | 4·00E-05 | 3·84E-04 | 388·84 | 440·23 | 0·88 |
| e11761 | 2263·91 | 33·50 | ADGEAGRPGSSGPSGDEGQPGEPGP | P12109 | COL6A1 | Collagen alpha-1(VI) chain | 1·86E-03 | 9·64E-03 | 31·75 | 9·45 | 3·36 |
| e02718 | 1200·54 | 26·31 | GEKGEAGDpGRp | P12109 | COL6A1 | Collagen alpha-1(VI) chain | 3·17E-06 | 3·95E-05 | 526·59 | 93·88 | 5·61 |
| e16018 | 2975·44 | 20·97 | QGPPGSATAKGERGFPGADGRpGSPGRAGNPG | Q02388 | COL7A1 | Collagen alpha-1(VII) chain | 6·46E-04 | 3·96E-03 | 10·45 | 38·76 | 0·27 |
| e20262 | 4414·02 | 28·99 | FDGQPGPKGDQGEKGERGTPGIGGFPGPSGNDGSAG-PPGPpGSVGpR | Q02388 | COL7A1 | Collagen alpha-1(VII) chain | 1·69E-08 | 5·22E-07 | 67·04 | 162·45 | 0·41 |
| e16156 | 3007·42 | 21·01 | QGPpGSATAKGERGFpGADGRpGSPGRAGNPG | Q02388 | COL7A1 | Collagen alpha-1(VII) chain | 3·18E-05 | 3·15E-04 | 202·78 | 322·08 | 0·63 |

| Peptide ID | Mass [Da] | CE-time [Min] | Sequence | UniprotID | Symbol | Protein name | p-value | adj_p-value (BH) | average_cases | average_controls | Fold change |
|---|---|---|---|---|---|---|---|---|---|---|---|
| e05288 | 1473·63 | 22·33 | FPGQTGPRGEMGQp | Q02388 | COL7A1 | Collagen alpha-1(VII) chain | 8·02E-04 | 4·71E-03 | 237·84 | 343·28 | 0·69 |
| e20082 | 4304·04 | 23·05 | GDRGFPGPLGEAGEKGERGPPGPAGSRGLpGVAGRp-GAKGpEGPPG | Q02388 | COL7A1 | Collagen alpha-1(VII) chain | 2·42E-04 | 1·73E-03 | 51·46 | 17·21 | 2·99 |
| e13387 | 2511·14 | 27·32 | KGEKGDSEDGApGLpGQPGSpGEQGp | Q02388 | COL7A1 | Collagen alpha-1(VII) chain | 1·70E-03 | 8·92E-03 | 220·43 | 64·20 | 3·43 |
| e14783 | 2751·35 | 21·71 | LPGQVGETGKpGApGRDGASGKDGDRGSP | Q02388 | COL7A1 | Collagen alpha-1(VII) chain | 3·78E-04 | 2·54E-03 | 118·24 | 29·77 | 3·97 |
| e09719 | 1971·94 | 24·77 | RGDPGPQGppGLALGERGpP | Q02388 | COL7A1 | Collagen alpha-1(VII) chain | 9·00E-03 | 3·45E-02 | 870·05 | 207·60 | 4·19 |
| e11417 | 2216·08 | 22·36 | KGEKGDSGASGREGFPGVpGGTGP | Q02388 | COL7A1 | Collagen alpha-1(VII) chain | 1·08E-13 | 2·24E-11 | 2764·09 | 538·29 | 5·13 |
| e00748 | 954·49 | 24·73 | KGDKGEAGPP | Q02388 | COL7A1 | Collagen alpha-1(VII) chain | 7·63E-05 | 6·73E-04 | 44·86 | 6·59 | 6·81 |
| e03037 | 1236·56 | 27·18 | GDRGEpGpPGPpG | Q02388 | COL7A1 | Collagen alpha-1(VII) chain | 5·36E-08 | 1·40E-06 | 195·91 | 26·51 | 7·39 |
| e07939 | 1764·83 | 24·47 | pGpLGEAGEKGERGpPGp | Q02388 | COL7A1 | Collagen alpha-1(VII) chain | 3·47E-04 | 2·35E-03 | 91·43 | 9·89 | 9·25 |
| e07215 | 1691·79 | 31·85 | GSPGppGpAGIATKGLNGP | Q03692 | COL10A1 | Collagen alpha-1(X) chain | 9·31E-03 | 3·53E-02 | 15·74 | 32·05 | 0·49 |
| e07224 | 1692·77 | 23·47 | GIpGFPGSKGDPGSPGPPG | Q03692 | COL10A1 | Collagen alpha-1(X) chain | 4·01E-04 | 2·65E-03 | 224·84 | 160·34 | 1·40 |
| e07216 | 1691·81 | 23·67 | KGDPGSPGPpGpAGIATKG | Q03692 | COL10A1 | Collagen alpha-1(X) chain | 3·42E-07 | 5·86E-06 | 62·12 | 4·11 | 15·13 |
| e02502 | 1176·56 | 26·86 | PGPQGPTGpSGpP | Q03692 | COL10A1 | Collagen alpha-1(X) chain | 2·92E-08 | 8·07E-07 | 87·10 | 3·62 | 24·04 |
| e02940 | 1226·53 | 21·11 | SGGDGppGpPGER | P12107 | COL11A1 | Collagen alpha-1(XI) chain | 1·95E-12 | 2·63E-10 | 175·00 | 446·62 | 0·39 |
| e18973 | 3757·74 | 32·85 | PGEPGpAGQDGVGGDKGEDGDpGQPGPPGPSGEAGPPGPPGK | P12107 | COL11A1 | Collagen alpha-1(XI) chain | 7·76E-03 | 3·04E-02 | 19·10 | 42·43 | 0·45 |
| e12945 | 2423·00 | 33·98 | DGpQGPpGSVGSVGGVGEKGEPGEAGN | P12107 | COL11A1 | Collagen alpha-1(XI) chain | 1·97E-04 | 1·45E-03 | 101·17 | 149·24 | 0·68 |
| e07389 | 1708·78 | 32·15 | GpAGEKGApGEKGpQGPAG | P12107 | COL11A1 | Collagen alpha-1(XI) chain | 4·96E-03 | 2·18E-02 | 191·11 | 271·70 | 0·70 |
| e04847 | 1425·59 | 22·34 | GGDkGEDGDPGQPGp | P12107 | COL11A1 | Collagen alpha-1(XI) chain | 9·64E-06 | 1·09E-04 | 1496·82 | 2101·28 | 0·71 |
| e17283 | 3265·44 | 35·90 | GPAGQDGVGGDKGEDGDpGQpGPPGPSGEAGPPGpPG | P12107 | COL11A1 | Collagen alpha-1(XI) chain | 8·36E-03 | 3·25E-02 | 201·77 | 254·40 | 0·79 |
| e05053 | 1445·67 | 28·49 | GPMGPPGppGpRGPQ | P12107 | COL11A1 | Collagen alpha-1(XI) chain | 4·68E-04 | 3·05E-03 | 442·57 | 178·38 | 2·48 |
| e17634 | 3346·67 | 19·25 | EGQSGEKGALGppGpQGPIGYPGPRGVKGADGVRG | P12107 | COL11A1 | Collagen alpha-1(XI) chain | 2·97E-03 | 1·43E-02 | 166·79 | 44·10 | 3·78 |

| Peptide ID | Mass [Da] | CE-time [Min] | Sequence | UniprotID | Symbol | Protein name | p-value | adj_p-value (BH) | average_cases | average_controls | Fold change |
|---|---|---|---|---|---|---|---|---|---|---|---|
| e02106 | 1134·60 | 21·20 | pGPpGKRGPpGA | P12107 | COL11A1 | Collagen alpha-1(XI) chain | 2·19E-09 | 9·65E-08 | 571·81 | 117·82 | 4·85 |
| e16701 | 3131·45 | 31·20 | QGPpGPqGpNGLSIPGEQGRQGmKGDAGEPGL | Q99715 | COL12A1 | Collagen alpha-1(XII) chain | 5·65E-03 | 2·40E-02 | 18·34 | 31·76 | 0·58 |
| e16294 | 3039·40 | 34·39 | QPGDKGERGAAGEQGpDGpKGSKGEPGKGEM | Q5TAT6 | COL13A1 | Collagen alpha-1(XIII) chain | 6·38E-03 | 2·63E-02 | 2502·06 | 2883·20 | 0·87 |
| e07594 | 1730·77 | 30·12 | PGHPGpKGDMGLTGpPGQ | Q5TAT6 | COL13A1 | Collagen alpha-1(XIII) chain | 1·16E-02 | 4·20E-02 | 571·73 | 362·33 | 1·58 |
| e07106 | 1681·74 | 30·91 | EAGEKGNPGAEGLQGVpG | Q5TAT6 | COL13A1 | Collagen alpha-1(XIII) chain | 1·02E-03 | 5·73E-03 | 1111·95 | 469·34 | 2·37 |
| e08610 | 1844·91 | 41·11 | SQASIQGppGPPGPpGPSGp | Q5TAT6 | COL13A1 | Collagen alpha-1(XIII) chain | 1·24E-09 | 6·05E-08 | 512·04 | 104·66 | 4·89 |
| e17892 | 3417·50 | 22·03 | KGERGAAGEQGpDGPKGSKGEPGKGEMVDYNGNI | Q5TAT6 | COL13A1 | Collagen alpha-1(XIII) chain | 2·69E-05 | 2·73E-04 | 57·00 | 9·57 | 5·96 |
| e15200 | 2820·31 | 23·85 | GIpGGVGSpGRDGSpGQRGLPGKDGSSGPpG | Q05707 | COL14A1 | Collagen alpha-1(XIV) chain | 4·15E-07 | 6·94E-06 | 509·07 | 215·42 | 2·36 |
| e17139 | 3232·52 | 22·12 | KGEKGNpGVGTQGPRGPPGPAGPSGESRPGSPGPP | Q05707 | COL14A1 | Collagen alpha-1(XIV) chain | 3·36E-04 | 2·29E-03 | 79·40 | 22·31 | 3·56 |
| e06381 | 1603·74 | 30·02 | VQGppGEPGRPGSpGAP | Q05707 | COL14A1 | Collagen alpha-1(XIV) chain | 7·23E-03 | 2·90E-02 | 335·23 | 74·14 | 4·52 |
| e15611 | 2892·33 | 34·39 | QKGEQGFEGSKGETGEKGEQGEKGDpAL | Q14993 | COL19A1 | Collagen alpha-1(XIX) chain | 1·04E-07 | 2·37E-06 | 384·24 | 773·02 | 0·50 |
| e19753 | 4098·99 | 21·17 | GPEGPSGKpGINGKDGIPGAQGImGKpGDRGpKGERGDQGIP | Q14993 | COL19A1 | Collagen alpha-1(XIX) chain | 1·02E-07 | 2·33E-06 | 3599·92 | 1873·29 | 1·92 |
| e03306 | 1263·56 | 26·70 | GSDGPPGKpGppGP | Q14993 | COL19A1 | Collagen alpha-1(XIX) chain | 5·33E-04 | 3·38E-03 | 52·25 | 6·42 | 8·13 |
| e17504 | 3318·40 | 22·73 | TDVFMGPpGSpGEDGPAGEPGPPGPEGQPGVDGATG | P39059 | COL15A1 | Collagen alpha-1(XV) chain | 1·45E-06 | 1·98E-05 | 61·49 | 124·23 | 0·49 |
| e06922 | 1659·74 | 29·44 | GPpGFGRpGDpGPPGPPG | P39059 | COL15A1 | Collagen alpha-1(XV) chain | 8·80E-03 | 3·40E-02 | 1114·93 | 724·22 | 1·54 |
| e12388 | 2340·21 | 26·00 | GEKGDPGNRGLPGPPGKKGQAGPpG | P39059 | COL15A1 | Collagen alpha-1(XV) chain | 3·41E-05 | 3·36E-04 | 722·59 | 381·07 | 1·90 |
| e17239 | 3255·50 | 36·06 | GppGLPGIPGKPGTDVFMGPPGSPGEDGpAGEpGP | P39059 | COL15A1 | Collagen alpha-1(XV) chain | 3·88E-03 | 1·79E-02 | 62·99 | 24·21 | 2·60 |
| e02207 | 1145·48 | 37·24 | GDPGpPGPpGPpG | P39059 | COL15A1 | Collagen alpha-1(XV) chain | 8·92E-03 | 3·43E-02 | 5·33 | 2·03 | 2·63 |
| e07459 | 1715·77 | 30·45 | FGRpGDpGPPGPPGppGP | P39059 | COL15A1 | Collagen alpha-1(XV) chain | 5·50E-03 | 2·36E-02 | 238·55 | 83·24 | 2·87 |
| e01845 | 1106·48 | 36·92 | PGVMGpPGppGP | P39059 | COL15A1 | Collagen alpha-1(XV) chain | 9·13E-10 | 4·76E-08 | 354·20 | 55·66 | 6·36 |

| Pep-tide ID | Mass [Da] | CE-time [Min] | Sequence | UniprotID | Symbol | Protein name | p-value | adj_p-value (BH) | aver-age_cases | aver-age_con-trols | Fold change |
|---|---|---|---|---|---|---|---|---|---|---|---|
| e05421 | 1491·65 | 23·57 | HpGppGEPGTDGAAGK | Q07092 | COL16A1 | Collagen alpha-1(XVI) chain | 2·71E-04 | 1·89E-03 | 29·33 | 70·72 | 0·41 |
| e10130 | 2029·85 | 20·34 | SAGEkGEPGPPGSEGLPGppGP | Q07092 | COL16A1 | Collagen alpha-1(XVI) chain | 1·29E-03 | 7·03E-03 | 25·08 | 51·81 | 0·48 |
| e11495 | 2226·96 | 33·46 | GNSGEKGDQGFQGQPGFPGPpGP | Q07092 | COL16A1 | Collagen alpha-1(XVI) chain | 5·11E-03 | 2·23E-02 | 62·97 | 76·76 | 0·82 |
| e15408 | 2854·31 | 21·56 | pAGERGHPGAPGPSGSPGLPGVPGSMGDMVN | Q07092 | COL16A1 | Collagen alpha-1(XVI) chain | 1·55E-04 | 1·20E-03 | 109·59 | 119·45 | 0·92 |
| e17903 | 3421·57 | 26·00 | AGERGHPGApGPSGSPGLpGVPGSMGDMVNYDEIK | Q07092 | COL16A1 | Collagen alpha-1(XVI) chain | 1·51E-03 | 8·03E-03 | 532·62 | 563·06 | 0·95 |
| e10847 | 2131·99 | 32·55 | KGEPGppGQPGYPGATGPpGLPG | Q07092 | COL16A1 | Collagen alpha-1(XVI) chain | 1·62E-03 | 8·54E-03 | 222·42 | 76·68 | 2·90 |
| e09708 | 1970·91 | 24·92 | AAGKEGPPGKQGFYGPpGpK | Q07092 | COL16A1 | Collagen alpha-1(XVI) chain | 3·21E-06 | 3·98E-05 | 1422·60 | 367·06 | 3·88 |
| e07614 | 1731·82 | 40·63 | GIAGENGLPGppGPQGppG | Q07092 | COL16A1 | Collagen alpha-1(XVI) chain | 9·19E-09 | 3·34E-07 | 497·71 | 88·22 | 5·64 |
| e06980 | 1666·83 | 23·69 | GPPGPAGERGHPGApGPSG | Q07092 | COL16A1 | Collagen alpha-1(XVI) chain | 2·40E-04 | 1·72E-03 | 1555·12 | 206·28 | 7·54 |
| e12694 | 2382·15 | 22·71 | GPAGPRGERGPQGNSGEKGDQGFQ | Q07092 | COL16A1 | Collagen alpha-1(XVI) chain | 1·41E-04 | 1·13E-03 | 273·84 | 31·69 | 8·64 |
| e08891 | 1874·86 | 24·43 | QRGEEGPPGMRGSPGPpGP | Q07092 | COL16A1 | Collagen alpha-1(XVI) chain | 5·42E-11 | 4·28E-09 | 617·26 | 56·32 | 10·96 |
| e04769 | 1417·64 | 20·06 | pGPpGpKGDQGPpGP | Q9UMD9 | COL17A1 | Collagen alpha-1(XVII) chain | 3·99E-03 | 1·83E-02 | 310·57 | 372·57 | 0·83 |
| e06856 | 1651·77 | 23·31 | PGSFLSNSETFLSGpP | Q9UMD9 | COL17A1 | Collagen alpha-1(XVII) chain | 6·16E-03 | 2·59E-02 | 48·38 | 51·14 | 0·95 |
| e12384 | 2340·04 | 22·58 | EGpmGQRGREGPMGpRGEAGPpG | Q9UMD9 | COL17A1 | Collagen alpha-1(XVII) chain | 1·21E-02 | 4·34E-02 | 460·78 | 152·38 | 3·02 |
| e11512 | 2228·08 | 22·45 | SGPPGPPGPPGpKGDQGpPGPRGH | Q9UMD9 | COL17A1 | Collagen alpha-1(XVII) chain | 1·10E-02 | 4·06E-02 | 46·01 | 13·12 | 3·51 |
| e15902 | 2951·44 | 22·27 | AGEPGPHGPPGVPGSVGPKGSSGSPGPQGPPGPV | Q9UMD9 | COL17A1 | Collagen alpha-1(XVII) chain | 1·54E-11 | 1·60E-09 | 2304·97 | 183·22 | 12·58 |
| e13779 | 2577·25 | 24·62 | DDILASPPRLPEPQPYPGAPHHSS | P39060 | COL18A1 | Collagen alpha-1(XVIII) chain | 2·71E-03 | 1·32E-02 | 151·58 | 130·76 | 1·16 |
| e13289 | 2490·24 | 24·64 | DDILASPPRLPEPQPYPGAPHHS | P39060 | COL18A1 | Collagen alpha-1(XVIII) chain | 3·30E-04 | 2·26E-03 | 334·60 | 155·70 | 2·15 |
| e16025 | 2976·44 | 20·46 | pAGpALQTVpGPQGPPGPPGRDGTPGRDGEP | P39060 | COL18A1 | Collagen alpha-1(XVIII) chain | 2·79E-05 | 2·81E-04 | 93·08 | 37·28 | 2·50 |
| e00977 | 990·55 | 20·58 | NVAKGIRSF | P39060 | COL18A1 | Collagen alpha-1(XVIII) chain | 2·11E-03 | 1·06E-02 | 55·91 | 17·15 | 3·26 |
| e05744 | 1531·69 | 40·62 | pGGGGFFGSSLpGpPGP | P39060 | COL18A1 | Collagen alpha-1(XVIII) chain | 7·63E-05 | 6·73E-04 | 52·47 | 13·09 | 4·01 |

| Pep-tide ID | Mass [Da] | CE-time [Min] | Sequence | UniprotID | Symbol | Protein name | p-value | adj_p-value (BH) | aver-age_cases | aver-age_con-trols | Fold change |
|---|---|---|---|---|---|---|---|---|---|---|---|
| e08853 | 1871·83 | 31·17 | PGDPGEDGKpGDTGpQGFp | P39060 | COL18A1 | Collagen alpha-1(XVIII) chain | 1·06E-02 | 3·94E-02 | 239·99 | 34·34 | 6·99 |
| e07751 | 1745·78 | 24·11 | TSAESpDApEENIAGVGA | P39060 | COL18A1 | Collagen alpha-1(XVIII) chain | 5·94E-04 | 3·69E-03 | 123·41 | 9·50 | 12·99 |
| e05725 | 1529·69 | 29·50 | MRGMPGpPGpPGPPGP | P39060 | COL18A1 | Collagen alpha-1(XVIII) chain | 7·46E-03 | 2·96E-02 | 181·79 | 13·68 | 13·29 |
| e05830 | 1540·74 | 39·81 | GPpGVPGpPGpGGSPGLP | Q8NFW1 | COL22A1 | Collagen alpha-1(XXII) chain | 3·84E-04 | 2·57E-03 | 2000·07 | 2966·42 | 0·67 |
| e16303 | 3041·38 | 29·95 | pGppGIAGPQGSQGERGADGEVGQKGDQGHPG | Q8NFW1 | COL22A1 | Collagen alpha-1(XXII) chain | 5·40E-05 | 5·00E-04 | 920·48 | 1262·79 | 0·73 |
| e05677 | 1523·67 | 22·09 | pGKDGDTGPTGPQGPQ | Q8NFW1 | COL22A1 | Collagen alpha-1(XXII) chain | 3·06E-03 | 1·46E-02 | 2777·91 | 3552·76 | 0·78 |
| e05961 | 1556·74 | 40·01 | pGVpGPPGPGGSPGLPGE | Q8NFW1 | COL22A1 | Collagen alpha-1(XXII) chain | 1·21E-02 | 4·34E-02 | 455·12 | 510·23 | 0·89 |
| e15160 | 2814·34 | 20·02 | SpGSRGLPGFPGPQGPAGRDGAPGNPGERG | Q8NFW1 | COL22A1 | Collagen alpha-1(XXII) chain | 1·55E-07 | 3·21E-06 | 401·42 | 342·50 | 1·17 |
| e05644 | 1518·75 | 22·90 | KpGPpGpTGpPGKDGP | Q8NFW1 | COL22A1 | Collagen alpha-1(XXII) chain | 2·41E-03 | 1·19E-02 | 38·98 | 18·49 | 2·11 |
| e20946 | 5508·53 | 25·31 | GTEGKKGEAGPPGLPGPpGIAGpQGSQGERGADG-EVGQKGDQGHPGVPGFMGPPGNPGP | Q8NFW1 | COL22A1 | Collagen alpha-1(XXII) chain | 4·14E-04 | 2·73E-03 | 637·20 | 280·55 | 2·27 |
| e09908 | 1995·95 | 24·98 | GPTGPQGPQGPRGPPGKNGSPG | Q8NFW1 | COL22A1 | Collagen alpha-1(XXII) chain | 4·77E-15 | 2·18E-12 | 553·02 | 121·64 | 4·55 |
| e19272 | 3869·82 | 20·69 | GPPGPPGVPGPPGPGGSPGLPGEIGFPGKPGPPGpTGpPGKDGP | Q8NFW1 | COL22A1 | Collagen alpha-1(XXII) chain | 8·79E-05 | 7·54E-04 | 1582·89 | 341·92 | 4·63 |
| e10073 | 2019·96 | 25·52 | GLpGFpGpQGPAGRDGAPGNPG | Q8NFW1 | COL22A1 | Collagen alpha-1(XXII) chain | 2·11E-03 | 1·06E-02 | 1028·54 | 199·80 | 5·15 |
| e05046 | 1444·72 | 28·93 | GPpGKDGpNGpPGPPG | Q8NFW1 | COL22A1 | Collagen alpha-1(XXII) chain | 3·69E-14 | 1·21E-11 | 205·86 | 22·66 | 9·09 |
| e14558 | 2708·19 | 21·85 | GKKGDDGTPSQpGppGPKGEpGSMGPRG | Q86Y22 | COL23A1 | Collagen alpha-1(XXIII) chain | 2·34E-05 | 2·42E-04 | 149·02 | 19·84 | 7·51 |
| e18759 | 3677·85 | 26·69 | pGRTGLAGApGpPGVKGSSGLPGSPGIQGpKGEQGLPGQP | Q17RW2 | COL24A1 | Collagen alpha-1(XXIV) chain | 2·38E-07 | 4·52E-06 | 530·97 | 191·80 | 2·77 |
| e01856 | 1107·50 | 36·82 | pGLQGpSGPpGP | Q17RW2 | COL24A1 | Collagen alpha-1(XXIV) chain | 4·38E-03 | 1·97E-02 | 14·88 | 5·10 | 2·92 |
| e10311 | 2054·05 | 21·58 | RGPKGDTGPPGPPGPAGIpGpSG | Q17RW2 | COL24A1 | Collagen alpha-1(XXIV) chain | 7·63E-07 | 1·14E-05 | 493·08 | 128·41 | 3·84 |
| e19485 | 3981·92 | 22·14 | GEKGVMGYPGPPGVPGpIGPLGLPGHVGARGPPGSQGPKGQR | Q17RW2 | COL24A1 | Collagen alpha-1(XXIV) chain | 3·35E-03 | 1·58E-02 | 3462·69 | 220·94 | 15·67 |
| e09548 | 1950·85 | 35·51 | GTDGpMGpHGpAGPKGERGE | Q9BXS0 | COL25A1 | Collagen alpha-1(XXV) chain | 6·85E-08 | 1·65E-06 | 62·55 | 187·73 | 0·33 |

| Peptide ID | Mass [Da] | CE-time [Min] | Sequence | UniprotID | Symbol | Protein name | p-value | adj_p-value (BH) | average_cases | average_controls | Fold change |
|---|---|---|---|---|---|---|---|---|---|---|---|
| e04814 | 1422·62 | 22·68 | KGTDGpMGPHGpAGp | Q9BXS0 | COL25A1 | Collagen alpha-1(XXV) chain | 2·31E-03 | 1·15E-02 | 182·43 | 233·84 | 0·78 |
| e15228 | 2824·28 | 21·50 | HGPPGpmGPHGLPGPKGTDGPMGPHGpAGp | Q9BXS0 | COL25A1 | Collagen alpha-1(XXV) chain | 1·86E-05 | 1·96E-04 | 124·01 | 27·91 | 4·44 |
| e13619 | 2548·26 | 27·12 | ANGMKGEKGDSGMPGPQGPSIIGPPGp | Q9BXS0 | COL25A1 | Collagen alpha-1(XXV) chain | 8·97E-14 | 2·21E-11 | 443·08 | 91·11 | 4·86 |
| e00142 | 842·37 | 34·86 | SGMPGpQGP | Q9BXS0 | COL25A1 | Collagen alpha-1(XXV) chain | 8·45E-03 | 3·28E-02 | 18·44 | 3·48 | 5·29 |
| e00600 | 935·45 | 23·82 | GRpGPpGPpG | Q96A83 | COL26A1 | Collagen alpha-1(XXVI) chain | 1·31E-03 | 7·13E-03 | 31·89 | 63·31 | 0·50 |
| e06279 | 1591·68 | 39·72 | PGppGPEGFPGDIGpPG | Q8IZC6 | COL27A1 | Collagen alpha-1(XXVII) chain | 1·28E-02 | 4·56E-02 | 52·26 | 16·99 | 3·08 |
| e07469 | 1716·77 | 28·12 | KGDpGDpGpPGTHGNPGI | Q2UY09 | COL28A1 | Collagen alpha-1(XXVIII) chain | 1·19E-03 | 6·57E-03 | 129·39 | 197·00 | 0·66 |
| e14200 | 2641·26 | 19·60 | PMGIpGIGSQGEQGIQGpIGpPGPQGPA | Q2UY09 | COL28A1 | Collagen alpha-1(XXVIII) chain | 2·61E-06 | 3·38E-05 | 3202·62 | 312·54 | 10·25 |
| e10993 | 2154·98 | 33·18 | FAGEKGPSGEAGTAGPpGTpGPQG | P08123 | COL1A2 | Collagen alpha-2(I) chain | 6·17E-04 | 3·81E-03 | 4·78 | 20·43 | 0·23 |
| e17585 | 3334·56 | 31·03 | EPGSAGPQGPPGPSGEEGKRGPNGEAGSAGPPGppGL | P08123 | COL1A2 | Collagen alpha-2(I) chain | 2·90E-08 | 8·07E-07 | 25·53 | 107·78 | 0·24 |
| e17751 | 3375·57 | 31·83 | GEPGSAGPQGPPGPSGEEGKRGPNGEAGSAGPPGpPGL | P08123 | COL1A2 | Collagen alpha-2(I) chain | 9·63E-14 | 2·21E-11 | 165·69 | 568·62 | 0·29 |
| e04647 | 1405·64 | 20·15 | DGPpGRDGQpGHKG | P08123 | COL1A2 | Collagen alpha-2(I) chain | 1·92E-06 | 2·54E-05 | 14·85 | 50·82 | 0·29 |
| e13424 | 2518·09 | 21·13 | EAGRDGNpGNDGPpGRDGQpGHKGE | P08123 | COL1A2 | Collagen alpha-2(I) chain | 2·46E-03 | 1·21E-02 | 2·76 | 8·93 | 0·31 |
| e10635 | 2096·93 | 20·27 | DGppGRDGQpGHKGERGYpG | P08123 | COL1A2 | Collagen alpha-2(I) chain | 2·17E-04 | 1·57E-03 | 3·32 | 10·47 | 0·32 |
| e15108 | 2805·30 | 23·80 | LKGQpGApGVKGEpGApGENGTPGQTGARG | P08123 | COL1A2 | Collagen alpha-2(I) chain | 6·88E-03 | 2·79E-02 | 13·11 | 32·92 | 0·40 |
| e06288 | 1592·74 | 19·54 | EDGHpGKpGRpGERG | P08123 | COL1A2 | Collagen alpha-2(I) chain | 2·30E-05 | 2·39E-04 | 60·84 | 144·24 | 0·42 |
| e19541 | 4005·96 | 22·25 | EVGKpGERGLHGEFGLPGPAGpRGERGpPGESGAAGPTGPIG | P08123 | COL1A2 | Collagen alpha-2(I) chain | 1·47E-03 | 7·91E-03 | 26·53 | 56·09 | 0·47 |
| e18867 | 3719·69 | 32·15 | QGFQGPAGEPGEPGQTGPAGARGPAGppGKAGEDGHpGKP | P08123 | COL1A2 | Collagen alpha-2(I) chain | 5·90E-08 | 1·50E-06 | 278·80 | 538·20 | 0·52 |
| e09557 | 1951·86 | 32·09 | VNGApGEAGRDGNpGNDGPpG | P08123 | COL1A2 | Collagen alpha-2(I) chain | 7·79E-05 | 6·81E-04 | 29·66 | 57·07 | 0·52 |
| e06154 | 1576·75 | 19·51 | EDGHpGKPGRpGERG | P08123 | COL1A2 | Collagen alpha-2(I) chain | 5·65E-07 | 8·99E-06 | 164·10 | 314·00 | 0·52 |

| Peptide ID | Mass [Da] | CE-time [Min] | Sequence | UniprotID | Symbol | Protein name | p-value | adj_p-value (BH) | average_cases | average_controls | Fold change |
|---|---|---|---|---|---|---|---|---|---|---|---|
| e19107 | 3801·79 | 33·48 | DQGPVGRTGEVGAVGPpGFAGEKGpSGEAGTAGPPGTpGPQG | P08123 | COL1A2 | Collagen alpha-2(I) chain | 1·60E-04 | 1·23E-03 | 115·99 | 221·47 | 0·52 |
| e18573 | 3616·72 | 33·07 | DQGPVGRTGEVGAVGpPGFAGEKGPSGEAGTAGPpGTpGP | P08123 | COL1A2 | Collagen alpha-2(I) chain | 9·14E-03 | 3·49E-02 | 51·38 | 95·30 | 0·54 |
| e05074 | 1447·70 | 19·50 | DGHpGKpGRPGERG | P08123 | COL1A2 | Collagen alpha-2(I) chain | 3·86E-04 | 2·58E-03 | 166·08 | 295·51 | 0·56 |
| e19752 | 4098·90 | 24·55 | GEVGPAGpNGFAGpAGAAGQp-GAKGERGAKGPKGENGVVGPTGpVG | P08123 | COL1A2 | Collagen alpha-2(I) chain | 1·82E-08 | 5·42E-07 | 517·66 | 917·25 | 0·56 |
| e05209 | 1463·69 | 19·50 | DGHpGKpGRpGERG | P08123 | COL1A2 | Collagen alpha-2(I) chain | 3·52E-03 | 1·65E-02 | 73·40 | 128·95 | 0·57 |
| e11265 | 2194·97 | 20·12 | NDGPpGRDGQpGHKGERGYpG | P08123 | COL1A2 | Collagen alpha-2(I) chain | 7·78E-05 | 6·81E-04 | 58·69 | 102·96 | 0·57 |
| e19745 | 4097·86 | 24·65 | SKGESGNKGEpGSAGPQGPpGpSGEEGKRGPNGEAGSAGPPGPpG | P08123 | COL1A2 | Collagen alpha-2(I) chain | 1·36E-08 | 4·55E-07 | 791·47 | 1350·19 | 0·59 |
| e06182 | 1579·68 | 23·04 | GpAGPRGERGPpGESGA | P08123 | COL1A2 | Collagen alpha-2(I) chain | 5·21E-07 | 8·41E-06 | 313·20 | 520·18 | 0·60 |
| e16869 | 3169·50 | 25·46 | ATGDRGEAGAAGPAGPAGpRGSPGERGEVGPAGPNG | P08123 | COL1A2 | Collagen alpha-2(I) chain | 8·63E-07 | 1·26E-05 | 55·48 | 90·86 | 0·61 |
| e17869 | 3410·58 | 31·13 | PAGATGDRGEAGAAGPAGPAGpRGSpGERGEVGPAGPNG | P08123 | COL1A2 | Collagen alpha-2(I) chain | 4·00E-05 | 3·84E-04 | 161·33 | 248·93 | 0·65 |
| e18907 | 3735·73 | 27·23 | TGDPGKNGDKGHAGLAGARGAPGpDGNNGAQGppGPQGVQG | P08123 | COL1A2 | Collagen alpha-2(I) chain | 5·23E-05 | 4·87E-04 | 119·90 | 175·24 | 0·68 |
| e05559 | 1508·66 | 24·06 | GpAGpRGERGPPGESG | P08123 | COL1A2 | Collagen alpha-2(I) chain | 9·13E-03 | 3·49E-02 | 60·25 | 86·63 | 0·70 |
| e00644 | 941·44 | 20·27 | EpGEKGPRG | P08123 | COL1A2 | Collagen alpha-2(I) chain | 4·47E-03 | 2·00E-02 | 183·91 | 258·84 | 0·71 |
| e17212 | 3248·55 | 30·60 | RTGEVGAVGPpGFAGEKGPSGEAGTAGpPGTpGPQG | P08123 | COL1A2 | Collagen alpha-2(I) chain | 7·79E-07 | 1·14E-05 | 1386·48 | 1938·84 | 0·72 |
| e10518 | 2080·93 | 20·25 | DGPpGRDGQpGHKGERGYpG | P08123 | COL1A2 | Collagen alpha-2(I) chain | 6·08E-07 | 9·54E-06 | 195·29 | 270·40 | 0·72 |
| e15948 | 2960·36 | 29·70 | LVGEpGPAGSKGESGNKGEpGSAGPQGPpGPSG | P08123 | COL1A2 | Collagen alpha-2(I) chain | 3·34E-03 | 1·58E-02 | 13·36 | 18·22 | 0·73 |
| e03691 | 1303·55 | 27·13 | YDGKGVGLGPGPMG | P08123 | COL1A2 | Collagen alpha-2(I) chain | 1·13E-03 | 6·30E-03 | 895·43 | 1194·72 | 0·75 |
| e16547 | 3092·44 | 36·22 | TGEVGAVGPpGFAGEKGPSGEAGTAGpPGTpGPQG | P08123 | COL1A2 | Collagen alpha-2(I) chain | 6·89E-03 | 2·79E-02 | 146·32 | 188·13 | 0·78 |
| e00421 | 902·42 | 20·86 | DpGKNGDKG | P08123 | COL1A2 | Collagen alpha-2(I) chain | 1·22E-03 | 6·67E-03 | 408·30 | 510·10 | 0·80 |

| Peptide ID | Mass [Da] | CE-time [Min] | Sequence | UniprotID | Symbol | Protein name | p-value | adj_p-value (BH) | average_cases | average_controls | Fold change |
|---|---|---|---|---|---|---|---|---|---|---|---|
| e17279 | 3264·53 | 30·51 | RTGEVGAVGPpGFAGEKGpSGEAGTAGpPGTpGPQG | P08123 | COL1A2 | Collagen alpha-2(I) chain | 5·07E-03 | 2·22E-02 | 406·55 | 484·76 | 0·84 |
| e11186 | 2184·96 | 33·01 | GPpGpAGSRGDGGPpGMTGFpGAAG | P08123 | COL1A2 | Collagen alpha-2(I) chain | 2·28E-03 | 1·14E-02 | 116·20 | 120·20 | 0·97 |
| e16390 | 3058·44 | 30·62 | pNGPPGpAGSRGDGGpPGMTGFPGAAGRTGPpGP | P08123 | COL1A2 | Collagen alpha-2(I) chain | 1·43E-02 | 4·94E-02 | 151·19 | 153·26 | 0·99 |
| e07217 | 1691·81 | 30·35 | GSpGNIGPAGKEGPVGLpG | P08123 | COL1A2 | Collagen alpha-2(I) chain | 1·10E-02 | 4·06E-02 | 1425·90 | 1302·50 | 1·09 |
| e14368 | 2674·20 | 20·11 | EAGRDGNpGNDGpPGRDGQpGHKGER | P08123 | COL1A2 | Collagen alpha-2(I) chain | 8·83E-04 | 5·07E-03 | 27·33 | 24·16 | 1·13 |
| e15400 | 2853·33 | 23·85 | LkGQpGApGVkGEpGApGENGTpGQTGARG | P08123 | COL1A2 | Collagen alpha-2(I) chain | 1·22E-03 | 6·67E-03 | 1202·08 | 937·52 | 1·28 |
| e08442 | 1822·74 | 30·83 | AGEKGPSGEAGTAGPpGTpGP | P08123 | COL1A2 | Collagen alpha-2(I) chain | 2·86E-03 | 1·37E-02 | 308·51 | 223·29 | 1·38 |
| e06165 | 1577·78 | 20·65 | RGLpGERGRVGApGPA | P08123 | COL1A2 | Collagen alpha-2(I) chain | 7·85E-04 | 4·63E-03 | 52·96 | 34·83 | 1·52 |
| e19692 | 4066·97 | 21·12 | GERGAAGIpGGKGEKGEpGLRGEIGNpGRDGARGAPGAVGApGP | P08123 | COL1A2 | Collagen alpha-2(I) chain | 5·87E-03 | 2·47E-02 | 293·46 | 181·47 | 1·62 |
| e02795 | 1209·53 | 26·33 | GPpGPDGNKGEpG | P08123 | COL1A2 | Collagen alpha-2(I) chain | 1·72E-03 | 8·95E-03 | 1014·71 | 590·18 | 1·72 |
| e11629 | 2244·08 | 19·54 | AGPpGKAGEDGHpGKpGRpGERG | P08123 | COL1A2 | Collagen alpha-2(I) chain | 2·00E-04 | 1·46E-03 | 444·11 | 237·17 | 1·87 |
| e13891 | 2593·33 | 19·91 | ARGPAGPpGKAGEDGHPGKPGRpGERG | P08123 | COL1A2 | Collagen alpha-2(I) chain | 8·29E-05 | 7·19E-04 | 61·79 | 31·25 | 1·98 |
| e08482 | 1828·01 | 24·57 | KEGPVGLpGIDGRPGPIGP | P08123 | COL1A2 | Collagen alpha-2(I) chain | 1·04E-03 | 5·84E-03 | 58·94 | 28·22 | 2·09 |
| e17105 | 3224·66 | 25·40 | TGAKGAAGLpGVAGApGLpGPRGIpGPVGAAGATGARG | P08123 | COL1A2 | Collagen alpha-2(I) chain | 1·14E-08 | 4·00E-07 | 699·32 | 310·27 | 2·25 |
| e19324 | 3891·77 | 24·39 | EPGSAGPQGPPGPSGEEGKRGPNGEAGSAGPPGppGLRGSpGS | P08123 | COL1A2 | Collagen alpha-2(I) chain | 1·66E-04 | 1·26E-03 | 3573·68 | 1492·25 | 2·39 |
| e13572 | 2542·19 | 23·08 | HGEFGLpGPAGPRGERGpPGESGAAGP | P08123 | COL1A2 | Collagen alpha-2(I) chain | 7·28E-10 | 4·37E-08 | 150·10 | 58·67 | 2·56 |
| e20265 | 4416·15 | 22·51 | GpAGKDGRTGHPGTVGPAGIRGPQGHQGPAG-PPGPPGPPGPPGVSGGGY | P08123 | COL1A2 | Collagen alpha-2(I) chain | 1·40E-07 | 2·99E-06 | 22284·79 | 6957·90 | 3·20 |
| e18344 | 3548·69 | 21·50 | KGPTGDpGKNGDKGHAGLAGARGApGPDGNNGAQGPpGP | P08123 | COL1A2 | Collagen alpha-2(I) chain | 3·90E-06 | 4·68E-05 | 299·74 | 89·43 | 3·35 |
| e10513 | 2079·96 | 25·58 | RGSpGSRGLpGADGRAGVMGPP | P08123 | COL1A2 | Collagen alpha-2(I) chain | 6·11E-08 | 1·54E-06 | 2534·44 | 719·53 | 3·52 |
| e10832 | 2129·98 | 32·86 | GVQGGKGEQGPPGPPGFQGLpGP | P08123 | COL1A2 | Collagen alpha-2(I) chain | 1·20E-06 | 1·71E-05 | 132·70 | 36·78 | 3·61 |

| Peptide ID | Mass [Da] | CE-time [Min] | Sequence | UniprotID | Symbol | Protein name | p-value | adj_p-value (BH) | average_cases | average_controls | Fold change |
|---|---|---|---|---|---|---|---|---|---|---|---|
| e09972 | 2005·92 | 25·12 | GEEGKRGPNGEAGSAGPpGPpG | P08123 | COL1A2 | Collagen alpha-2(I) chain | 1·28E-05 | 1·38E-04 | 189·51 | 52·03 | 3·64 |
| e11611 | 2242·03 | 26·40 | IRGPQGHQGPAGPpGPpGPpGPpG | P08123 | COL1A2 | Collagen alpha-2(I) chain | 2·72E-14 | 1·04E-11 | 537·58 | 140·63 | 3·82 |
| e10318 | 2054·95 | 25·44 | GEpGPAGSKGESGNKGEpGSAGP | P08123 | COL1A2 | Collagen alpha-2(I) chain | 1·54E-04 | 1·20E-03 | 381·34 | 92·79 | 4·11 |
| e08859 | 1871·93 | 21·28 | SGPpGPpGPAGKEGLRGPRG | P08123 | COL1A2 | Collagen alpha-2(I) chain | 2·90E-11 | 2·66E-09 | 182·21 | 43·02 | 4·24 |
| e05999 | 1560·81 | 23·09 | KEGPVGLpGIDGRPGP | P08123 | COL1A2 | Collagen alpha-2(I) chain | 7·08E-10 | 4·37E-08 | 405·43 | 88·76 | 4·57 |
| e13555 | 2538·23 | 23·10 | ISGPpGPpGPAGKEGLRGPRGDQGPVG | P08123 | COL1A2 | Collagen alpha-2(I) chain | 6·94E-05 | 6·26E-04 | 217·39 | 47·22 | 4·60 |
| e15742 | 2920·37 | 21·93 | SKGESGNKGEpGSAGPQGPPGPSGEEGKRGp | P08123 | COL1A2 | Collagen alpha-2(I) chain | 4·93E-09 | 1·88E-07 | 214·34 | 44·93 | 4·77 |
| e17219 | 3249·55 | 30·71 | RTGEVGAVGPpGFAGEKGpSGEAGTAGpPGTPGPqG | P08123 | COL1A2 | Collagen alpha-2(I) chain | 8·09E-10 | 4·57E-08 | 502·06 | 105·04 | 4·78 |
| e16873 | 3171·59 | 20·85 | VGKpGERGLHGEFGLpGPAGPRGERGppGESG | P08123 | COL1A2 | Collagen alpha-2(I) chain | 4·37E-04 | 2·85E-03 | 73·08 | 13·73 | 5·32 |
| e04648 | 1405·66 | 28·35 | GISGPpGPpGPAGKEG | P08123 | COL1A2 | Collagen alpha-2(I) chain | 4·05E-07 | 6·82E-06 | 368·86 | 68·66 | 5·37 |
| e11510 | 2228·05 | 19·61 | AGPpGKAGEDGHpGKpGRPGERG | P08123 | COL1A2 | Collagen alpha-2(I) chain | 8·64E-05 | 7·44E-04 | 395·70 | 66·45 | 5·95 |
| e12693 | 2382·14 | 19·80 | GPAGpPGKAGEDGHPGKpGRpGERG | P08123 | COL1A2 | Collagen alpha-2(I) chain | 6·40E-03 | 2·64E-02 | 214·83 | 34·97 | 6·14 |
| e01185 | 1020·52 | 25·27 | NAGPAGPAGPRG | P08123 | COL1A2 | Collagen alpha-2(I) chain | 4·51E-05 | 4·29E-04 | 58·31 | 9·33 | 6·25 |
| e11382 | 2212·09 | 19·64 | AGPpGKAGEDGHPGKPGRpGERG | P08123 | COL1A2 | Collagen alpha-2(I) chain | 1·12E-09 | 5·55E-08 | 132·16 | 17·76 | 7·44 |
| e08582 | 1841·85 | 24·22 | VGEPGpAGSKGESGNKGEpG | P08123 | COL1A2 | Collagen alpha-2(I) chain | 2·53E-03 | 1·24E-02 | 121·82 | 15·69 | 7·76 |
| e09366 | 1928·90 | 24·97 | VGEPGpAGSKGESGNKGEpGS | P08123 | COL1A2 | Collagen alpha-2(I) chain | 6·71E-03 | 2·74E-02 | 259·92 | 32·73 | 7·94 |
| e04292 | 1363·64 | 28·57 | KGEPGSAGpQGPpGP | P08123 | COL1A2 | Collagen alpha-2(I) chain | 9·99E-03 | 3·73E-02 | 526·93 | 64·11 | 8·22 |
| e07405 | 1710·75 | 31·05 | EKGpSGEAGTAGPpGTpGP | P08123 | COL1A2 | Collagen alpha-2(I) chain | 4·83E-04 | 3·12E-03 | 188·88 | 20·02 | 9·43 |
| e00157 | 845·40 | 23·40 | SpGERGEV | P08123 | COL1A2 | Collagen alpha-2(I) chain | 2·04E-03 | 1·04E-02 | 110·26 | 10·13 | 10·89 |
| e01938 | 1115·56 | 26·55 | YDGKGVGLGPGP | P08123 | COL1A2 | Collagen alpha-2(I) chain | 8·18E-10 | 4·57E-08 | 220·79 | 16·31 | 13·54 |
| e00536 | 924·45 | 24·12 | GRVGApGPAGA | P08123 | COL1A2 | Collagen alpha-2(I) chain | 5·42E-05 | 5·00E-04 | 245·48 | 11·18 | 21·95 |

| Peptide ID | Mass [Da] | CE-time [Min] | Sequence | UniprotID | Symbol | Protein name | p-value | adj_p-value (BH) | average_cases | average_controls | Fold change |
|---|---|---|---|---|---|---|---|---|---|---|---|
| e02963 | 1228·58 | 27·34 | FpGAAGRTGppGP | P08123 | COL1A2 | Collagen alpha-2(I) chain | 1·63E-07 | 3·33E-06 | 276·41 | 10·08 | 27·42 |
| e02105 | 1134·59 | 23·99 | PIGQEGAPGRPG | P08572 | COL4A2 | Collagen alpha-2(IV) chain | 3·35E-05 | 3·31E-04 | 36·53 | 76·10 | 0·48 |
| e16610 | 3109·40 | 29·91 | DTGNPGAPGTpGTKGWAGDSGpQGRpGVFGLPG | P08572 | COL4A2 | Collagen alpha-2(IV) chain | 5·49E-04 | 3·45E-03 | 33·29 | 66·51 | 0·50 |
| e11768 | 2264·94 | 43·13 | pGFPGAQGEPGSQGEpGDpGLpGP | P08572 | COL4A2 | Collagen alpha-2(IV) chain | 8·54E-04 | 4·95E-03 | 103·97 | 33·56 | 3·10 |
| e15535 | 2877·32 | 24·25 | GETGPHGYKGMVGAIGATGPpGEEGpRGpP | Q14055 | COL9A2 | Collagen alpha-2(IX) chain | 2·57E-07 | 4·74E-06 | 148·90 | 186·25 | 0·80 |
| e12015 | 2297·99 | 33·82 | PpGppGPpGVPGSDGIDGDNGPPGK | Q14055 | COL9A2 | Collagen alpha-2(IX) chain | 1·32E-02 | 4·64E-02 | 290·63 | 327·35 | 0·89 |
| e06314 | 1595·69 | 22·81 | PGpKGEpGKAGpDGpDG | Q14055 | COL9A2 | Collagen alpha-2(IX) chain | 8·71E-04 | 5·03E-03 | 126·90 | 142·09 | 0·89 |
| e14936 | 2776·34 | 21·50 | VGMMGPPGpPGPPGYpGKQGpHGHpGPR | Q14055 | COL9A2 | Collagen alpha-2(IX) chain | 2·15E-13 | 4·08E-11 | 721·23 | 171·16 | 4·21 |
| e06668 | 1632·80 | 30·84 | ERGpPGpPGpPGVpGSD | Q14055 | COL9A2 | Collagen alpha-2(IX) chain | 1·24E-04 | 1·01E-03 | 564·58 | 94·48 | 5·98 |
| e08304 | 1807·80 | 23·88 | pGQpGTKGGpGDQGEPGPQ | Q14055 | COL9A2 | Collagen alpha-2(IX) chain | 1·11E-02 | 4·08E-02 | 81·49 | 13·11 | 6·22 |
| e15532 | 2876·35 | 34·48 | GPTGATGDKGPPGPVGpPGSNGPVGEPGPEGPA | P05997 | COL5A2 | Collagen alpha-2(V) chain | 4·87E-03 | 2·15E-02 | 7·84 | 36·08 | 0·22 |
| e20387 | 4523·06 | 26·21 | PGPVGpPGSNGPVGEPGPEGPAGNDGTPGRDGAVGERG-DRGDPGPAGLPG | P05997 | COL5A2 | Collagen alpha-2(V) chain | 3·80E-05 | 3·69E-04 | 11·91 | 36·71 | 0·32 |
| e18809 | 3697·72 | 27·26 | TGPMGAMGPLGPRGMPGERGRLGpQGApGQRGAHGMpG | P05997 | COL5A2 | Collagen alpha-2(V) chain | 2·07E-03 | 1·05E-02 | 13·36 | 38·29 | 0·35 |
| e06214 | 1583·65 | 39·73 | GSPGTSGppGSAGpPGSpG | P05997 | COL5A2 | Collagen alpha-2(V) chain | 2·39E-07 | 4·52E-06 | 198·82 | 558·01 | 0·36 |
| e16344 | 3049·35 | 29·64 | VGEpGpEGpAGNDGTPGRDGAVGERGDRGDPG | P05997 | COL5A2 | Collagen alpha-2(V) chain | 2·59E-07 | 4·74E-06 | 127·51 | 308·95 | 0·41 |
| e15594 | 2889·37 | 28·80 | PGSRGENGpTGAVGFAGPQGPDGQPGVKGEP | P05997 | COL5A2 | Collagen alpha-2(V) chain | 1·55E-07 | 3·21E-06 | 635·24 | 339·61 | 1·87 |
| e16571 | 3098·48 | 29·41 | LTGNpGVQGPEGKLGPLGApGEDGRpGpPGSIG | P05997 | COL5A2 | Collagen alpha-2(V) chain | 1·34E-02 | 4·68E-02 | 96·96 | 51·53 | 1·88 |
| e15320 | 2838·39 | 24·16 | TQGPPGATGFPGSAGRVGPpGpAGAPGPAGPLG | P05997 | COL5A2 | Collagen alpha-2(V) chain | 1·26E-05 | 1·37E-04 | 2050·94 | 985·83 | 2·08 |
| e03494 | 1283·55 | 27·39 | DKGDpGEDGQPGP | P05997 | COL5A2 | Collagen alpha-2(V) chain | 3·81E-03 | 1·77E-02 | 61·11 | 26·11 | 2·34 |

| Peptide ID | Mass [Da] | CE-time [Min] | Sequence | UniprotID | Symbol | Protein name | p-value | adj_p-value (BH) | average_cases | average_controls | Fold change |
|---|---|---|---|---|---|---|---|---|---|---|---|
| e04424 | 1379·63 | 28·20 | DKGpPGpVGPpGSNG | P05997 | COL5A2 | Collagen alpha-2(V) chain | 1·69E-07 | 3·40E-06 | 357·03 | 52·26 | 6·83 |
| e00028 | 809·32 | 34·76 | GEPGPEGPA | P05997 | COL5A2 | Collagen alpha-2(V) chain | 1·26E-03 | 6·86E-03 | 11·00 | 1·32 | 8·31 |
| e03206 | 1253·55 | 21·34 | PGTSGPpGSAGpPGS | P05997 | COL5A2 | Collagen alpha-2(V) chain | 1·31E-02 | 4·63E-02 | 996·08 | 65·96 | 15·10 |
| e16764 | 3148·46 | 31·26 | KGEpGLPGpPGEGRAGEPGTAGPTGPPGVPGSpGI | P25067 | COL8A2 | Collagen alpha-2(VIII) chain | 1·66E-05 | 1·76E-04 | 33·69 | 135·34 | 0·25 |
| e18758 | 3677·82 | 24·29 | pGAPGQGGAPGPPGLPGPAGLGKPGLDGLPGAPGDKGESGPPG | P25067 | COL8A2 | Collagen alpha-2(VIII) chain | 1·39E-03 | 7·51E-03 | 354·44 | 517·92 | 0·68 |
| e02699 | 1198·54 | 26·12 | GPpGFQGEpGPQ | P25067 | COL8A2 | Collagen alpha-2(VIII) chain | 8·36E-03 | 3·25E-02 | 180·36 | 105·43 | 1·71 |
| e13748 | 2571·16 | 34·64 | EGRAGEPGTAGPTGppGVPGSPGITGPpG | P25067 | COL8A2 | Collagen alpha-2(VIII) chain | 1·14E-02 | 4·16E-02 | 76·73 | 29·38 | 2·61 |
| e15401 | 2853·34 | 28·33 | GTAGPTGPPGVpGSpGITGpPGPPGPPGPPGAPG | P25067 | COL8A2 | Collagen alpha-2(VIII) chain | 3·33E-04 | 2·28E-03 | 534·37 | 97·58 | 5·48 |
| e09792 | 1980·91 | 25·56 | GPTGPKGEpGFTGRpGGPGVAG | P25067 | COL8A2 | Collagen alpha-2(VIII) chain | 1·24E-04 | 1·01E-03 | 4397·02 | 748·19 | 5·88 |
| e03383 | 1271·62 | 21·83 | KGEpGFTGRPGGP | P25067 | COL8A2 | Collagen alpha-2(VIII) chain | 1·28E-12 | 1·84E-10 | 330·17 | 17·58 | 18·78 |
| e17816 | 3397·56 | 31·76 | NGADGPQGPpGGVGNLGppGEKGEPGESGSPGIQGEP | P13942 | COL11A2 | Collagen alpha-2(XI) chain | 7·20E-07 | 1·10E-05 | 9·68 | 119·81 | 0·08 |
| e14918 | 2774·37 | 21·54 | pGPEGPAGLIGPPGIQGNPGPVGDPGERGP | P13942 | COL11A2 | Collagen alpha-2(XI) chain | 1·04E-03 | 5·85E-03 | 138·02 | 131·40 | 1·05 |
| e07199 | 1689·74 | 40·69 | pGPpGQQGTPGTQGLpGP | P13942 | COL11A2 | Collagen alpha-2(XI) chain | 4·82E-03 | 2·13E-02 | 2186·35 | 1623·47 | 1·35 |
| e20509 | 4654·14 | 25·78 | GEHGpPGPPGPIGPVGQPGAAGADGEPGARGPQGHF-GAKGDEGTRGFNGP | P13942 | COL11A2 | Collagen alpha-2(XI) chain | 7·29E-05 | 6·52E-04 | 12302·68 | 8672·79 | 1·42 |
| e09631 | 1962·89 | 41·85 | EPGppGQQGTpGTQGLpGPQ | P13942 | COL11A2 | Collagen alpha-2(XI) chain | 1·94E-03 | 9·97E-03 | 79·21 | 28·49 | 2·78 |
| e18396 | 3560·70 | 26·10 | GLPGPKGELALSGEKGDQGPPGDpGSpGSpGPAGPAGPPG | Q01955 | COL4A3 | Collagen alpha-3(IV) chain | 1·98E-04 | 1·45E-03 | 11·65 | 43·60 | 0·27 |
| e09534 | 1948·99 | 24·80 | EGPRGAQGLpGLNGLKGQQG | Q01955 | COL4A3 | Collagen alpha-3(IV) chain | 5·30E-04 | 3·38E-03 | 4269·30 | 6042·64 | 0·71 |
| e15360 | 2847·26 | 24·37 | GpKGDpGIpGLDRSGFpGETGSPGIPGHQ | Q01955 | COL4A3 | Collagen alpha-3(IV) chain | 7·81E-04 | 4·62E-03 | 332·44 | 456·93 | 0·73 |
| e07481 | 1717·90 | 23·77 | KGPPGDHGLPGYLGSPGI | Q01955 | COL4A3 | Collagen alpha-3(IV) chain | 3·97E-03 | 1·82E-02 | 1183·98 | 1074·49 | 1·10 |
| e09656 | 1964·99 | 24·86 | KpGTPGPAGEKGNKGSKGEPG | Q01955 | COL4A3 | Collagen alpha-3(IV) chain | 1·45E-02 | 4·98E-02 | 986·81 | 405·93 | 2·43 |

| Peptide ID | Mass [Da] | CE-time [Min] | Sequence | UniprotID | Symbol | Protein name | p-value | adj_p-value (BH) | average_cases | average_controls | Fold change |
|---|---|---|---|---|---|---|---|---|---|---|---|
| e10747 | 2115·92 | 32·77 | PGTpGNEGLDGpRGDPGQPGpP | Q01955 | COL4A3 | Collagen alpha-3(IV) chain | 9·23E-03 | 3·51E-02 | 59·12 | 21·19 | 2·79 |
| e18186 | 3502·75 | 19·29 | PGPAGEKGNKGSKGEPGpAGSDGLpGLKGKRGDSGSPA | Q01955 | COL4A3 | Collagen alpha-3(IV) chain | 1·40E-03 | 7·53E-03 | 991·13 | 337·71 | 2·93 |
| e01534 | 1068·47 | 26·76 | PGDpGSpGSpGP | Q01955 | COL4A3 | Collagen alpha-3(IV) chain | 2·70E-03 | 1·32E-02 | 101·03 | 19·41 | 5·21 |
| e18343 | 3548·62 | 26·82 | LPGDMGKKGEmGQPGPpGHLGpAGPEGAPGSPGSpGLP | Q01955 | COL4A3 | Collagen alpha-3(IV) chain | 2·17E-03 | 1·09E-02 | 49·14 | 4·97 | 9·90 |
| e17817 | 3398·57 | 31·83 | ELGEAGPSGEpGVPGDAGMPGERGEAGHRGSAGALGP | Q14050 | COL9A3 | Collagen alpha-3(IX) chain | 1·37E-06 | 1·88E-05 | 16·41 | 99·22 | 0·17 |
| e18022 | 3454·57 | 31·88 | PGpRGNQGDRGDKGAAGAGLDGpEGDQGPQGPQGVPG | Q14050 | COL9A3 | Collagen alpha-3(IX) chain | 1·14E-03 | 6·34E-03 | 27·34 | 94·64 | 0·29 |
| e19394 | 3926·83 | 33·54 | GDKGAAGAGLDGPEGDQGpQGpQGVpGTSKDGQDGAPGEPGPP | Q14050 | COL9A3 | Collagen alpha-3(IX) chain | 2·57E-04 | 1·81E-03 | 12·57 | 36·28 | 0·35 |
| e11916 | 2283·98 | 27·27 | QGDRGDKGAAGAGLDGpEGDQGpQ | Q14050 | COL9A3 | Collagen alpha-3(IX) chain | 1·94E-04 | 1·43E-03 | 119·80 | 185·95 | 0·64 |
| e12993 | 2431·28 | 26·91 | LpGPPGPKGAPGKPGKPGEAGLPGLPG | Q14050 | COL9A3 | Collagen alpha-3(IX) chain | 9·07E-04 | 5·19E-03 | 66·82 | 40·56 | 1·65 |
| e13568 | 2541·23 | 22·96 | GpTGYKGEQGEVGKDGEKGDPGPpGP | Q14050 | COL9A3 | Collagen alpha-3(IX) chain | 2·79E-07 | 5·02E-06 | 1629·80 | 617·22 | 2·64 |
| e09669 | 1965·99 | 24·88 | LGRPGPKGTPGVAGpSGEpGm | Q14050 | COL9A3 | Collagen alpha-3(IX) chain | 5·18E-04 | 3·32E-03 | 510·13 | 161·94 | 3·15 |
| e15047 | 2795·42 | 21·53 | RDGpPGpKGApGERGSLGPPGpPGLGGKGL | Q14050 | COL9A3 | Collagen alpha-3(IX) chain | 2·96E-09 | 1·21E-07 | 913·61 | 70·47 | 12·96 |
| e10547 | 2085·03 | 22·00 | GPpGPPGSIGHpGARGPpGYRG | Q14050 | COL9A3 | Collagen alpha-3(IX) chain | 1·14E-06 | 1·63E-05 | 86·27 | 3·12 | 27·68 |
| e17754 | 3376·63 | 31·72 | IDGSpGEKGDPGDVGGPGPPGASGEPGAPGPPGKRGPS | P25940 | COL5A3 | Collagen alpha-3(V) chain | 1·07E-08 | 3·83E-07 | 65·22 | 310·53 | 0·21 |
| e10797 | 2123·88 | 21·88 | DGSpGEKGDpGDVGGPGPPGASGE | P25940 | COL5A3 | Collagen alpha-3(V) chain | 1·94E-07 | 3·80E-06 | 1453·08 | 2001·57 | 0·73 |
| e01666 | 1084·52 | 21·20 | GSKGDKGDAGPP | P25940 | COL5A3 | Collagen alpha-3(V) chain | 3·19E-04 | 2·19E-03 | 104·27 | 52·77 | 1·98 |
| e18537 | 3603·77 | 19·35 | GAQGpPGSAGPPGYPGPRGVKGTSGNRGLQGEKGEKGE | P25940 | COL5A3 | Collagen alpha-3(V) chain | 2·34E-03 | 1·16E-02 | 518·52 | 148·23 | 3·50 |
| e11834 | 2273·97 | 26·85 | SPGEKGDPGDVGGPGPpGASGEpGAP | P25940 | COL5A3 | Collagen alpha-3(V) chain | 9·20E-03 | 3·51E-02 | 174·69 | 27·73 | 6·30 |
| e05952 | 1555·68 | 29·36 | VGPPGEpGPpGQQGNH | P25940 | COL5A3 | Collagen alpha-3(V) chain | 6·77E-04 | 4·10E-03 | 541·40 | 76·88 | 7·04 |
| e19151 | 3818·73 | 37·77 | GDLGpPGDpGVSGIDGSPGEKGDPGDVGGPGPPGASGEPGAPGP | P25940 | COL5A3 | Collagen alpha-3(V) chain | 3·48E-03 | 1·63E-02 | 414·58 | 31·96 | 12·97 |

| Peptide ID | Mass [Da] | CE-time [Min] | Sequence | UniprotID | Symbol | Protein name | p-value | adj_p-value (BH) | average_cases | average_controls | Fold change |
|---|---|---|---|---|---|---|---|---|---|---|---|
| e06294 | 1593·71 | 22·82 | PFGDDGLpGppGPkGP | P53420 | COL4A4 | Collagen alpha-4(IV) chain | 1·87E-04 | 1·39E-03 | 55·66 | 128·31 | 0·43 |
| e05380 | 1486·69 | 21·29 | PGDQGPPGpDGPRGAP | P53420 | COL4A4 | Collagen alpha-4(IV) chain | 5·93E-05 | 5·44E-04 | 87·61 | 125·54 | 0·70 |
| e08406 | 1818·83 | 24·27 | HEDATpGGKGFPGPLGpPG | P53420 | COL4A4 | Collagen alpha-4(IV) chain | 6·99E-03 | 2·83E-02 | 397·64 | 127·76 | 3·11 |
| e08096 | 1782·76 | 40·37 | GFpGLPGDQGEPGSpGppG | P53420 | COL4A4 | Collagen alpha-4(IV) chain | 1·12E-02 | 4·09E-02 | 47·95 | 11·39 | 4·21 |
| e08942 | 1878·85 | 32·22 | pGFPGTpGLpGmPGHDGAPG | P29400 | COL4A5 | Collagen alpha-5(IV) chain | 4·17E-04 | 2·74E-03 | 99·97 | 103·55 | 0·97 |
| e10156 | 2032·95 | 25·01 | DGIPGPAGQKGEPGQPGFGNPG | P29400 | COL4A5 | Collagen alpha-5(IV) chain | 1·19E-02 | 4·30E-02 | 24·51 | 16·56 | 1·48 |
| e06269 | 1590·67 | 40·03 | GLDGppGpDGLQGpPGP | P29400 | COL4A5 | Collagen alpha-5(IV) chain | 3·04E-03 | 1·45E-02 | 371·05 | 191·42 | 1·94 |
| e00320 | 880·39 | 23·62 | RGLDGPpGP | P29400 | COL4A5 | Collagen alpha-5(IV) chain | 4·20E-03 | 1·91E-02 | 64·33 | 18·34 | 3·51 |
| e07367 | 1706·77 | 40·58 | GDVGPNGQpGPmGPpGLp | P29400 | COL4A5 | Collagen alpha-5(IV) chain | 3·94E-06 | 4·68E-05 | 151·99 | 42·25 | 3·60 |
| e18802 | 3695·77 | 27·01 | GIPGLPGDPGYPGEPGRDGEKGQKGDTGppGppGLVIP | P29400 | COL4A5 | Collagen alpha-5(IV) chain | 3·72E-12 | 4·74E-10 | 242·78 | 61·69 | 3·94 |
| e12852 | 2407·21 | 22·70 | GpSITGVpGpAGLPGPKGEKGYPGIG | Q14031 | COL4A6 | Collagen alpha-6(IV) chain | 2·48E-04 | 1·76E-03 | 165·39 | 94·35 | 1·75 |
| e09844 | 1987·97 | 25·15 | GLDGERGRPGPAGppGppGPS | Q14031 | COL4A6 | Collagen alpha-6(IV) chain | 6·87E-03 | 2·79E-02 | 1122·72 | 575·45 | 1·95 |

**Supplementary Table 5:** Prediction of mortality in patients with critical condition stratified based on the COL210 score range.

| **Characteristic** | **HR**[1] | **95% CI**[1] | **p-value** |
|---|---|---|---|
| COL210 | | | |
| (-6,-2] | — | — | |
| (-2,-0·5] | 1·10 | 0·73, 1·67 | 0·6 |
| (-0·5,1] | 1·58 | 1·07, 2·34 | 0·021 |
| (1,6] | 1·76 | 1·19, 2·60 | 0·004 |
| Gender | | | |
| Male | — | — | |
| Female | 0·84 | 0·71, 1·00 | 0·056 |
| Kidney disease | | | |
| no | — | — | |
| yes | 1·63 | 1·36, 1·95 | <0·001 |
| Diabetes | | | |
| no | — | — | |
| yes | 0·96 | 0·78, 1·19 | 0·7 |
| CVD | | | |

| **Characteristic** | **HR**[1] | **95% CI**[1] | **p-value** |
|---|---|---|---|
| no | — | — | |
| yes | 1·25 | 0·93, 1·68 | 0·14 |
| ht | | | |
| no | — | — | |
| yes | 0·97 | 0·81, 1·17 | 0·8 |
| n = 1,713; N events = 574; statistic.log = 70.4; p.value.log = 0.000; statistic.sc = 69.0; p.value.sc = 0.000; statistic.wald = 67.3; p.value.wald = 0.000; statistic.robust = NA; p.value.robust = NA; $R^2$ = 0.040; r.squared.max = 0.966; c-index = 0.625; c-index SE = 0.013; Log-likelihood = -2,854; AIC = 5,723; BIC = 5,758; No. Obs. = 1,713 | | | |

**Supplementary Table 6:** Prediction of mortality in patients with non-critical condition stratified based on underlying co-morbidities.

| **Characteristic** | **HR[1]** | **95% CI[1]** | **p-value** |
|---|---|---|---|
| COL210 | | | |
| (-6,-2] | — | — | |
| (-2,-0.5] | 1·69 | 1·40, 2·03 | <0·001 |
| (-0.5,1] | 1·91 | 1·48, 2·45 | <0·001 |
| (1,6] | 2·20 | 1·56, 3·10 | <0·001 |
| Gender | | | |
| Male | — | — | |
| Female | 0·74 | 0·63, 0·88 | <0·001 |
| Kidney disease | | | |
| no | — | — | |
| Yes | 1·05 | 0·80, 1·38 | 0·7 |
| Diabetes | | | |
| no | — | — | |
| yes | 1·39 | 1·13, 1·70 | 0·001 |
| CVD | | | |

| Characteristic | HR[1] | 95% CI[1] | p-value |
|---|---|---|---|
| no | — | — | |
| yes | 3·15 | 2·62, 3·78 | <0·001 |
| ht | | | |
| no | — | — | |
| yes | 1·22 | 0·99, 1·51 | 0·062 |
| map | 0·99 | 0·98, 0·99 | <0·001 |
| egfr | 0·99 | 0·99, 1·00 | 0·020 |
| n = 7,474; N events = 625; statistic.log = 295; p.value.log = 0.000; statistic.sc = 302; p.value.sc = 0.000; statistic.wald = 284; p.value.wald = 0.000; statistic.robust = NA; p.value.robust = NA; $R^2$ = 0.039; r.squared.max = 0.605; c-index = 0.687; c-index SE = 0.016; Log-likelihood = -3,319; AIC = 6,658; BIC = 6,703; No. Obs. = 7,474 | | | |
